# Compact clinical features capturing organ history for dynamic prediction of organ deterioration in sepsis

**DOI:** 10.64898/2026.11.01.26367016

**Authors:** Luomeng Wang, Jialin Song, Marina Vereshchako, Xinran Law, Qingyu Zhou, Rongyang Zhou, Jiajun Zhu, David Luk, Hengbing Zhou, Yiming Sun, Hongmei Zi, Chuhan Zhong, Lu Zheng, Jianhua Zhang, Xihui Gao, Hongyong He, Shuying Chen

## Abstract

Organ scores can conceal recent instability. We studied confirmed six-hour deterioration in six organs with low recorded severity among 19,690 adults with sepsis in MIMIC-IV, eICU, CareVue and SICdb. Using a 175-variable model reduced from 552, we compared 3,072 combinations of 12 historical sources. With hourly predictions, at least four hours between alerts and 0.50 alerts per patient, the model using ten sources captured 319 eICU events versus 245 using current information alone (+30.2%; difference 74, 95% CI 49–106.5). Gains were 34.6% in CareVue (144 versus 107) and 81.5% in SICdb (49 versus 27); using six sources captured 312 eICU events. Training with incomplete records increased capture from 180 to 211 (+31) under 50% withholding. Retrieving one existing laboratory record improved timely correct organ selection from 145 to 159 (+9.7%). Repeated new low-severity measurements changed earlier history’s value. Models were evaluated without retraining in 1,661 adults with pancreatitis-related conditions.

## Introduction

Identical organ scores can conceal different clinical courses.^1,2^ An organ that has recently improved after dysfunction may carry a different risk from one that has remained stable. Current scores omit how long the abnormality lasted, the support required during recovery and the sequence of improvement. Recovery after acute kidney injury illustrates how apparent normalisation can coexist with continuing vulnerability to recurrent injury.^3^ We asked whether these earlier changes help identify which of six organs currently recorded at low severity is most likely to deteriorate over the next six hours.

Models using physiological time series can anticipate circulatory failure and cardiorespiratory instability.^4,5^ A systematic review has examined the predictive value of repeated measurements in critical care.^6^ Studies of organ trajectories also describe sepsis subphenotypes and predict deterioration. ^7,8^ The remaining question is how much earlier history adds once current physiology, treatment and measurement timing are known and the organ is recorded at low severity. Clinical usefulness also depends on which organ is selected for reassessment, when an alert is issued and how repeat alerts are suppressed within the available workload.^9^

Electronic health records contain detailed histories, but reviewing an entire ICU stay at every assessment is impractical. Differences in charting systems also make large sets of derived variables difficult to transfer between hospitals. A useful summary should improve prediction beyond current clinical information while requiring few additional record sources. The number of model variables alone does not describe this requirement: a derived organ state may rely on laboratory tests, ventilator settings and infusion records.^10,11^

We evaluated how much of the benefit of organ history could be retained using fewer record sources (Fig. 1). We compared all 3,072 distinct combinations, including a derived organ state only when the records needed to calculate it were available. We tested history beyond current physiology and measurement patterns and compared compact history from ten sources and brief history from six sources with the full 175-variable model. We then evaluated hourly organ reassessment, incomplete records, retrieval of existing laboratory results and changes in the value of earlier history as new measurements arrived.

**Figure 1.**
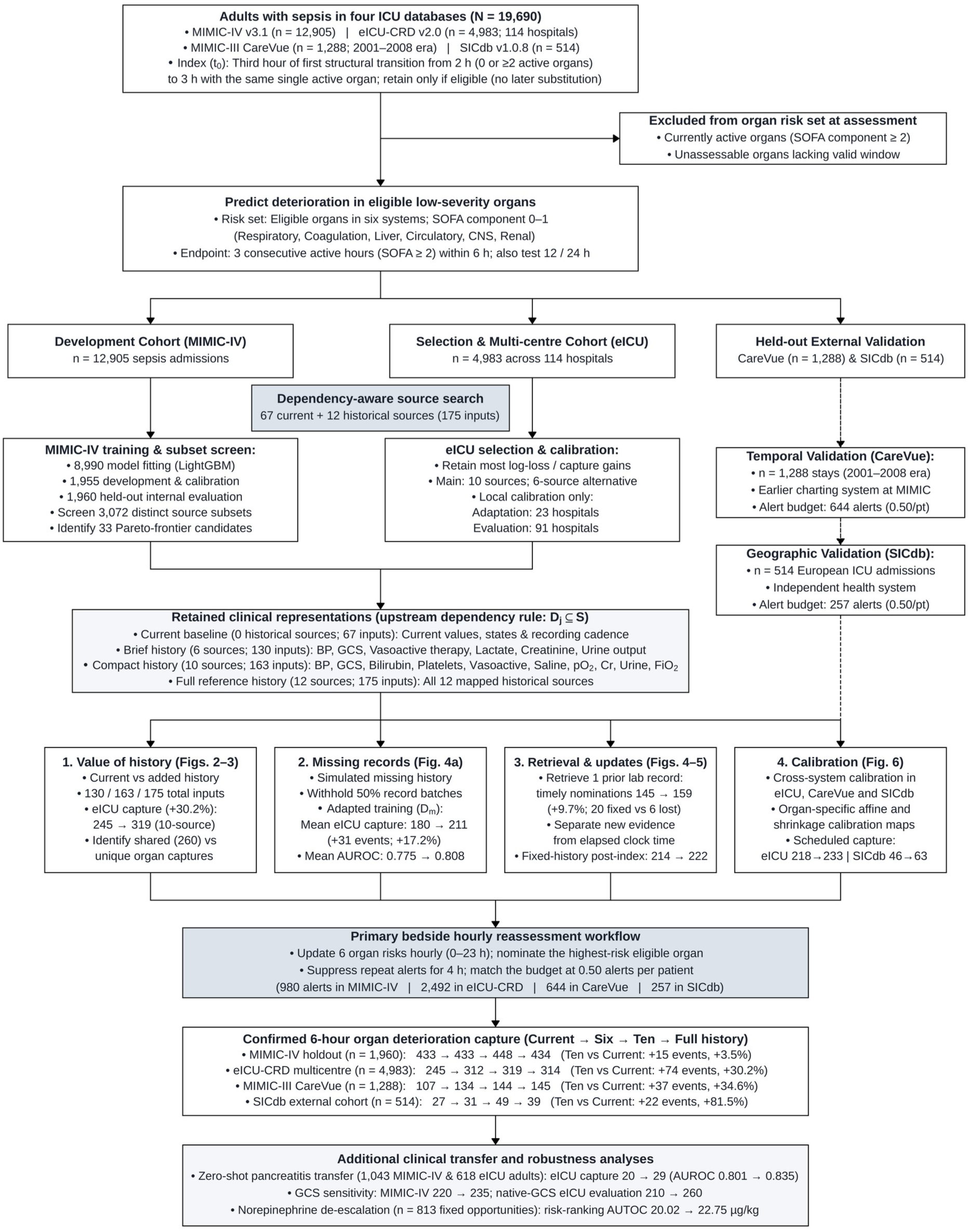
Study design and dynamic organ reassessment. Four databases supplied 19,690 adults with sepsis. MIMIC-IV supported development and held-out internal evaluation; eICU contributed to selecting the historical sources. CareVue provides earlier-era evaluation at the MIMIC institution, whereas SICdb provides geographic external evaluation. For each eligible low-severity organ, deterioration required three consecutive hourly states with SOFA component ≥2 confirmed within six hours. Source counts denote additional historical record sources; total inputs include the shared 67-input current-information baseline. The dependency rule D_ⱼ_ ⊆ S retains an expression only when its required sources D_ⱼ_ are included in the selected source set S. The 260 shared eICU captures are the events captured by both compact and full history. The 214-to-222 comparison concerns early-history down-weighting in the separate fixed-history, post-index experiment. The observed-range GCS analysis used strict component bounds for MIMIC-IV reconstruction and refitting among retained qualifying entries; eICU retained its native total-GCS definition. Local eICU calibration separated 23 adaptation hospitals from 91 evaluation hospitals. Each additional analysis uses its stated assessment schedule and population. That workflow refreshed risk hourly, nominated one eligible organ, and applied a four-hour patient cooldown at 0.50 alerts per patient. Arrows show the study organisation; CareVue and SICdb were separate evaluations after model selection. BP, blood pressure; GCS, Glasgow Coma Scale; CNS, central nervous system; pO2, oxygen partial pressure; Cr, creatinine; FiO2, inspired oxygen fraction; AUROC, area under the receiver operating characteristic curve; AUTOC, area under the targeting operator characteristic curve; Dm, missingness-adapted model.

## Results

### Historical information improves prediction beyond the current assessment

We studied 19,690 adults with sepsis across four databases: 12,905 in MIMIC-IV (8,990 fitting, 1,955 auxiliary development/calibration, and 1,960 held-out evaluation patients), 4,983 across 114 hospitals in eICU-CRD, 1,288 in MIMIC-III CareVue, and 514 in the European SICdb cohort (Fig. 1; Table 1; Supplementary Tables 1–2). At each assessment, we predicted confirmed deterioration (SOFA component >=2 sustained across three consecutive hourly positions) within six hours—with 12- and 24-hour extensions—in each of the six organ systems (respiratory, coagulation, hepatic, circulatory, central nervous, and renal) currently recorded below threshold (Fig. 1). MIMIC-IV supplied model training and internal evaluation, eICU supported selection using both databases, CareVue tested temporal shift across an earlier charting era, and SICdb provided held-out external validation.

**Table 1.**
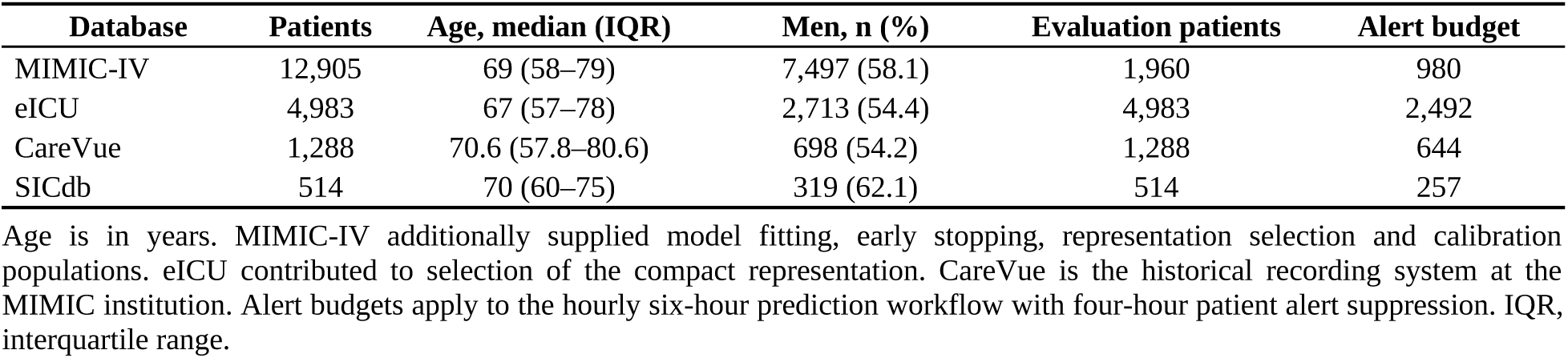
Study populations.

| Database | Patients | Age, median (IQR) | Men, n (%) | Evaluation patients | Alert budget |
|---|---|---|---|---|---|
| MIMIC-IV | 12,905 | 69 (58–79) | 7,497 (58.1) | 1,960 | 980 |
| eICU | 4,983 | 67 (57–78) | 2,713 (54.4) | 4,983 | 2,492 |
| CareVue | 1,288 | 70.6 (57.8–80.6) | 698 (54.2) | 1,288 | 644 |
| SICdb | 514 | 70 (60–75) | 319 (62.1) | 514 | 257 |

Age is in years. MIMIC-IV additionally supplied model fitting, early stopping, representation selection and calibration populations. eICU contributed to selection of the compact representation. CareVue is the historical recording system at the MIMIC institution. Alert budgets apply to the hourly six-hour prediction workflow with four-hour patient alert suppression. IQR, interquartile range.

Low current severity did not erase risk associated with prior organ involvement. Standardising for the active index organ and entry route, six-hour circulatory deterioration risk in MIMIC-IV was 15.69% with prior circulatory involvement versus 1.07% without it (6.33% versus 0.89% in eICU), and respiratory risk was 18.24% versus 7.44% in MIMIC-IV (11.55% versus 3.26% in eICU; Supplementary Fig. 1a; Supplementary Table 4). Excess risk also persisted in coagulation (8.49% versus 1.11% in MIMIC-IV; 6.08% versus 0.57% in eICU), renal (10.05% versus 1.09%; 3.40% versus 0.58%), and neurological systems (5.01% versus 1.55%; 7.44% versus 2.77%). Conditioning on each patient’s eligible organ set and total number of organs that deteriorated within the prediction horizon at each assessment confirmed that prior history localised which organ would deteriorate within the same patient, reducing conditional log loss by 0.1396 in MIMIC-IV (95% confidence interval 0.1231–0.1547) and 0.1093 in eICU (0.0654–0.1554; Supplementary Fig. 1b; Supplementary Table 5).

Across nine scheduled origins with six-hour alert suppression, the full model with 175 variables (67 current plus 108 historical columns across 12 sources) increased six-hour AUROC from 0.8979 to 0.9145 in MIMIC-IV and from 0.8418 to 0.8706 in eICU, reducing log loss from 0.08219 to 0.07867 and from 0.05406 to 0.05203 (Table 2; Supplementary Table 6). The compact history (163 columns) achieved AUROCs of 0.9114 and 0.8695, log losses of 0.07927 and 0.05173, and lifted eICU within-patient organ discrimination from 0.8421 to 0.8770. Scheduled six-hour capture (0.50 alerts/patient) increased from 363 to 382 in MIMIC-IV, 219 to 272 in eICU (282 with full history), 108 to 126 in CareVue, and 26 to 46 in SICdb (Table 2; Supplementary Table 39). Consistent gains held at 12 and 24 hours, with eICU median onset lead times of 2 hours (interquartile range 1–3) at 6 hours, 5 hours (2–7) at 12 hours, and 7 hours (3–14) at 24 hours (Supplementary Fig. 3a,b,d; Supplementary Tables 6 and 37).

**Table 2.** Six-hour prediction performance and hourly reassessment capture across representations and cohorts. Hourly capture denotes distinct confirmed six-hour organ deteriorations captured under hourly reassessment with a four-hour patient cooldown at a matched budget of 0.50 alerts per patient (980 alerts in MIMIC-IV, 2,492 in eICU, 644 in CareVue, and 257 in SICdb). Paired Δ vs Current (95% CI) gives the patient-cluster bootstrap difference relative to Current information (67 columns). Scheduled six-hour AUROC, patient-weighted log loss, within-patient organ AUROC, and scheduled capture are evaluated across the nine prespecified assessment origins with six-hour alert suppression (source calibration shown; values in brackets for CareVue and SICdb denote organ-specific local calibration).

| **Cohort & representation** | **Inputs (hist. sources)** | **Hourly capture** | **Paired Δ vs Current (95% CI)** | **6-h AUROC** | **Patient-weighted log loss** | **Organ AUROC** | **9-origin capture** |
|---|---|---|---|---|---|---|---|
| MIMIC-IV holdout (n = 1,960) |  |  |  |  |  |  |  |
| Current information | 67 (0) | 433 | Reference | 0.8979 | 0.08219 | 0.8877 | 363 |
| Brief history (6-source) | 130 (6) | 433 | 0 (−16 to 23) | 0.9098 | 0.07946 | 0.9079 | 374 |
| Compact history (10-source) | 163 (10) | 448 | +15 (−4 to 38) | 0.9114 | 0.07927 | 0.9103 | 382 |
| Full history (12-source) | 175 (12) | 434 | +1 (−21 to 24.52) | 0.9145 | 0.07867 | 0.9137 | 381 |
| eICU-CRD multicentre (n = 4,983) |  |  |  |  |  |  |  |
| Current information | 67 (0) | 245 | Reference | 0.8418 | 0.05406 | 0.8421 | 219 |
| Brief history (6-source) | 130 (6) | 312 | +67 (34.48 to 96) | 0.8662 | 0.05203 | 0.8732 | 276 |
| Compact history (10-source) | 163 (10) | 319 | +74 (49 to 106.5) | 0.8695 | 0.05173 | 0.8770 | 272 |
| Full history (12-source) | 175 (12) | 314 | +69 (41 to 101.52) | 0.8706 | 0.05203 | 0.8813 | 282 |
| MIMIC-III CareVue (n = 1,288) |  |  |  |  |  |  |  |
| Current information | 67 (0) | 107 | Reference | 0.8380 [0.8500] | 0.10094 [0.09422] | 0.8367 [0.8660] | 108 [105] |
| Brief history (6-source) | 130 (6) | 134 | +27 (3 to 45) | 0.8468 [0.8539] | 0.09931 [0.09310] | 0.8524 [0.8672] | 122 [126] |
| Compact history (10-source) | 163 (10) | 144 | +37 (14 to 54) | 0.8487 [0.8553] | 0.09904 [0.09284] | 0.8553 [0.8692] | 126 [129] |
| Full history (12-source) | 175 (12) | 145 | +38 (15 to 57.52) | 0.8514 [0.8580] | 0.09864 [0.09260] | 0.8586 [0.8677] | 117 [130] |
| SICdb external cohort (n = 514) |  |  |  |  |  |  |  |
| Current information | 67 (0) | 27 | Reference | 0.8591 [0.8788] | 0.09360 [0.08709] | 0.9069 [0.9226] | 26 [51] |
| Brief history (6-source) | 130 (6) | 31 | +4 (−5 to 11) | 0.8685 [0.8814] | 0.09482 [0.08612] | 0.9148 [0.9318] | 35 [58] |
| Compact history (10-source) | 163 (10) | 49 | +22 (8 to 35) | 0.8742 [0.8854] | 0.09507 [0.08556] | 0.9211 [0.9354] | 46 [63] |
| Full history (12-source) | 175 (12) | 39 | +12 (−1 to 20.52) | 0.8698 [0.8854] | 0.09548 [0.08555] | 0.9153 [0.9402] | 40 [61] |

Historical gains persisted in three comparisons addressing current information and measurement frequency. First, expanding the current baseline from 67 to 91 variables yielded 205 eICU captures (log loss 0.05427), whereas adding compact history on top of the 91-variable baseline reduced log loss to 0.05265 and increased capture to 270 (274 with full history; Supplementary Tables 6–7). Second, restricting the index assessment to observations with a target-organ measurement within the preceding three hours preserved historical gains in MIMIC-IV (AUROC 0.8742 to 0.8950) and eICU (0.7936 to 0.8405; Supplementary Table 8). Third, decomposing the model with 175 variables in eICU showed that current values/states captured 238 events (log loss 0.05450), adding recording-process indicators captured 219 (0.05406), adding 90 numerical-history variables captured 240 (0.05378), and adding the 18 derived organ-state history variables lifted capture to 282 (0.05203; Supplementary Fig. 3c; Supplementary Table 31), surpassing simple summary statistics plus state durations (256 captures; Supplementary Fig. 2d; Supplementary Table 11).

Increasing training sample size did not replace patient history. Across nested MIMIC-IV training sets of 2,247 (25%), 4,495 (50%), and 8,990 (100%) patients, models using current information captured 191, 202, and 219 eICU episodes, whereas models with target-organ history captured 233, 243, and 282 (Supplementary Fig. 2c; Supplementary Tables 9–10). A 25%-sample history model (233 captures, AUROC 0.8563) exceeded the 100%-sample model using current information (219 captures, AUROC 0.8418).

### Ten record sources retain substantial historical prediction gain

We began with 5,492 types of structured clinical records, yielding 272,043 derived variables. Selection left 17 record groups with 1,492 variables. We excluded 912 variables without an equivalent eICU mapping from training and removed 28 constant or unused variables from the remaining 580, leaving 552 candidates in 13 groups. We compared 27 model sizes, down to 38 variables, and selected 175 as the full reference (−68.3% derived variables). Compared with 552 variables, this model captured 381 rather than 387 MIMIC-IV events and 282 rather than 273 eICU events; 38 variables captured 361 and 254 events, respectively (Supplementary Fig. 2a,b; Supplementary Table 12). Within the full model, we considered 8,192 combinations of 12 historical sources and the pain-assessment group already included in the current baseline. A derived organ state could be used only when all records needed to calculate it were available. After merging combinations with identical model inputs, 3,072 remained, all sharing the 67 variables for current information (Fig. 2a; Supplementary Table 3). Three-fold cross-validation in MIMIC-IV identified 33 combinations on the Pareto frontier with 0 to 12 sources for evaluation in eICU (Fig. 2b; Supplementary Table 14). Selecting record groups, mapping between databases and pruning derived variables were separate steps.

**Figure 2.**
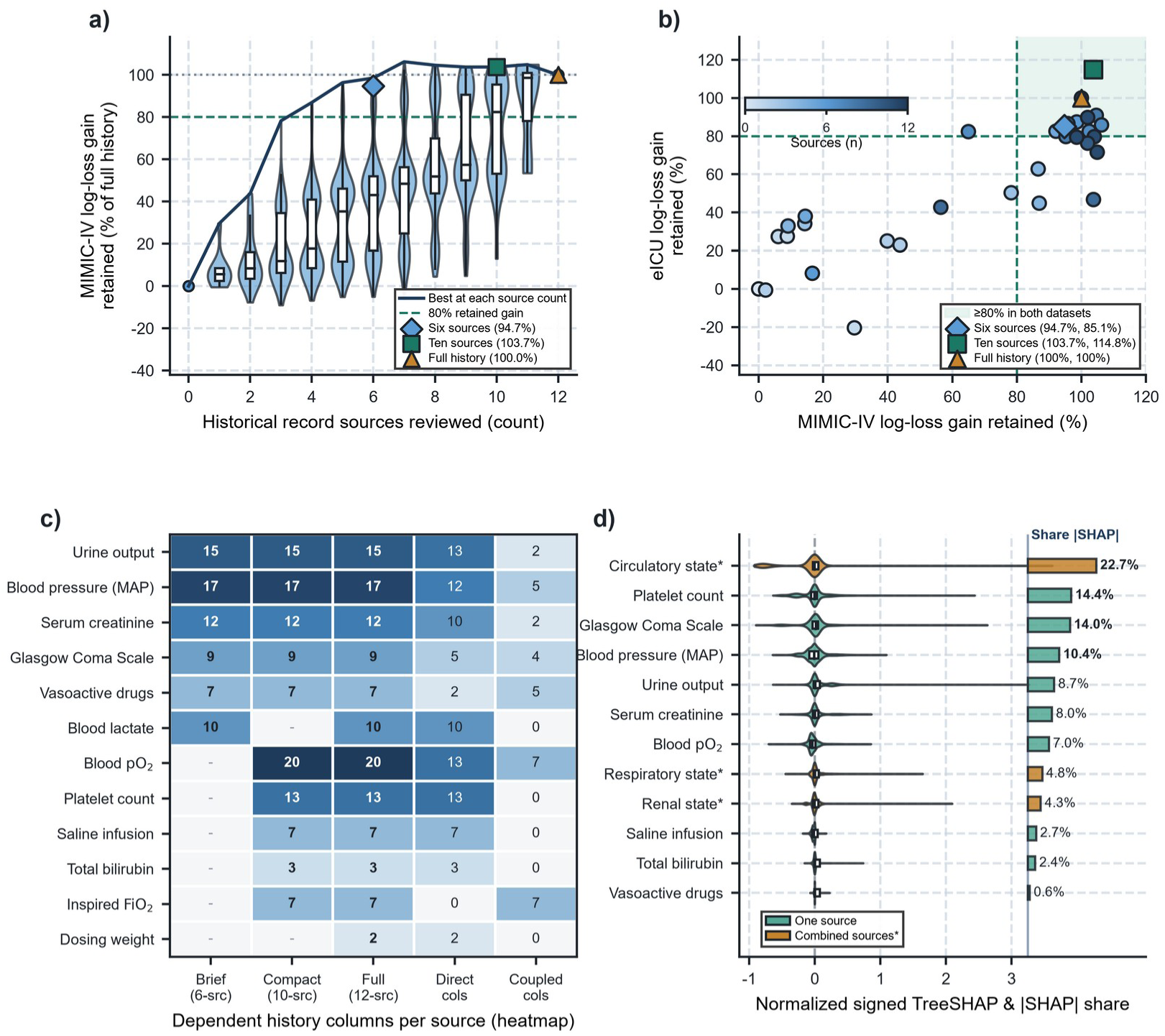
Record sources and retained prediction gains. a), Six-hour log-loss gain retained across 3,072 distinct input sets in MIMIC-IV. Violins show the empirical distribution; white boxes show medians and interquartile ranges, with whiskers at 1.5 times the interquartile range. The curve marks the best gain at each source count. b), Log-loss retention in MIMIC-IV and eICU for 33 source-frontier candidates; dashed lines mark 80%. c), Historical columns supplied by each source, including derived organ-state columns. Six, ten and full histories add 63, 96 and 108 columns to the common baseline with 67 variables. d), Signed TreeSHAP contributions on the log-odds scale for compact history in eICU; percentages give shares of mean absolute contribution. Asterisks identify joint dependencies counted once. GCS, Glasgow Coma Scale; MAP, mean arterial pressure; pO2, oxygen partial pressure; FiO2, inspired oxygen fraction.

The smallest combination meeting the prespecified requirement to retain >=80% of the full model’s log-loss and capture gains in both MIMIC-IV and eICU used ten sources: blood pressure, Glasgow Coma Scale (GCS), bilirubin, platelets, vasoactive therapy, saline infusion, blood pO2, creatinine, urine output and FiO2 (96 historical variables; 163 total inputs; Fig. 2c). The brief model used six sources: blood pressure, GCS, vasoactive therapy, lactate, creatinine and urine output (63 historical variables; 130 inputs). The two combinations differed: brief history included lactate, whereas compact history included five other sources; together they covered 11 sources (Fig. 2c). Brief history retained 85.1% of the eICU log-loss gain and 90.5% of the scheduled capture gain. Its 75.0% capture retention in MIMIC-IV development was below the joint 80% requirement (Supplementary Table 14).

At six hours, compact history retained 82.8% of full-history log-loss gain in the MIMIC-IV holdout cohort (95% confidence interval 65.4–97.6%) and 114.8% in eICU (97.9–150.2%), with scheduled capture retentions of 105.6% and 84.1%, respectively (Supplementary Table 13). Brief history retained 94.7% of source development log-loss gain and 85.1% in eICU (90.5% capture retention, 276 versus 282). At 12 and 24 hours, eICU log-loss retention for the compact model selected at six hours moderated to 53.3% and 61.7%.

In eICU TreeSHAP explanations for the compact model (Fig. 2d), coupled circulatory state history (blood pressure plus vasoactive support) contributed the largest share of mean absolute SHAP magnitude (22.7%), followed by platelets (14.4%), GCS (14.0%), blood pressure (10.4%), urine output (8.7%), creatinine (8.0%), blood pO2 (7.0%), coupled respiratory state (4.8%), and coupled renal state (4.3%). Variables requiring more than one source were counted once under the corresponding group of sources.

### Compact histories increase capture at an unchanged alert budget

We updated risk every hour and allowed at least four hours between alerts for a patient. Each model had the same budget of 0.50 alerts per patient (980 alerts in MIMIC-IV, 2,492 in eICU, 644 in CareVue and 257 in SICdb; Fig. 1).

In eICU, compact history captured 319 distinct organ deteriorations versus 245 with current information (+30.2%; difference +74, patient 95% confidence interval 49 to 106.5; hospital interval 43.3 to 106.9; Fig. 3a,b; Table 2; Supplementary Table 16). Capture rose from 107 to 144 in CareVue (+34.6%; difference +37, 14 to 54), from 27 to 49 in SICdb (+81.5%; difference +22, 8 to 35), and from 433 to 448 in MIMIC-IV (+3.5%; difference +15, −4 to 38). Brief (6-source) and full (175-column) histories captured 312 and 314 events in eICU, 134 and 145 in CareVue, and 31 and 39 in SICdb (Fig. 3a; Table 2). Sequential active query policies requesting 1–2 additional sources on demand from the 6-source baseline captured 297–306 eICU events, compared with 312 (fixed 6-source) and 319 (fixed 10-source; Supplementary Fig. 4c; Supplementary Table 29).

**Figure 3.**
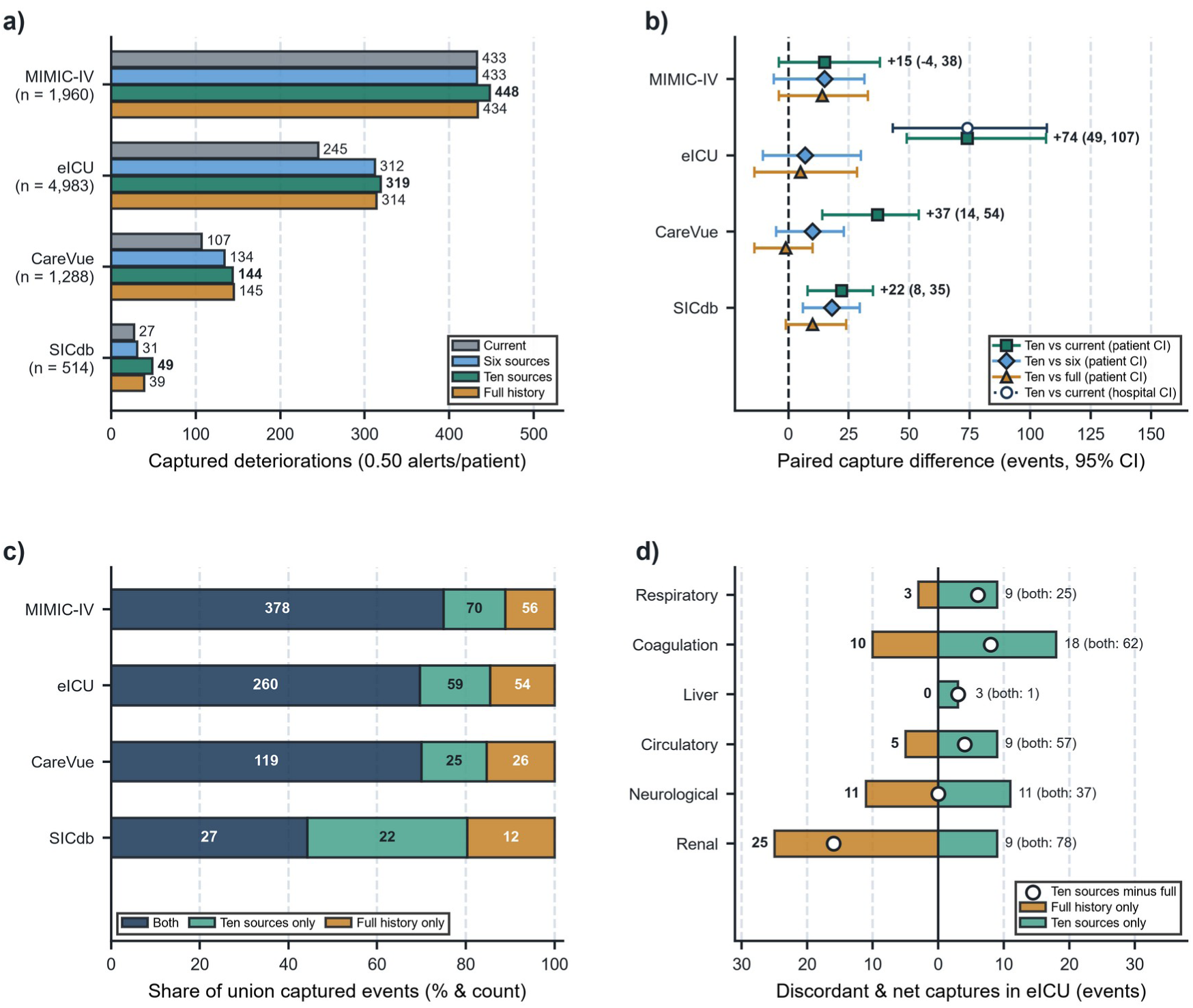
Event capture at equal alert budgets. a), Distinct deteriorations captured with hourly predictions and a four-hour patient cooldown. Budgets were 980, 2,492, 644 and 257 alerts in MIMIC-IV, eICU, CareVue and SICdb, respectively. b), Paired capture differences and 95% intervals from 500 patient-cluster bootstrap samples; the open eICU marker denotes hospital-cluster uncertainty. c), Shared captures, additions and losses for ten-source versus full history, expressed as counts and percentages of their union. d), eICU organ-specific events captured only by full history (left) or compact history (right); open circles give net differences and parentheses give shared captures. All comparisons retain the same eligible population, total alert budget and event-deduplication rule.

Similar total captures did not imply identical event coverage. In eICU, compact (319) and full (314) history shared 260 captures, with 59 unique to compact and 54 unique to full history (Fig. 3c; Supplementary Table 17). Compact history captured more unique events in coagulation (18 versus 10), respiratory (9 versus 3), circulatory (9 versus 5), and hepatic (3 versus 0) systems, matched neurological unique captures (11 versus 11), and captured fewer unique renal events (9 versus 25, alongside 78 shared renal captures; Fig. 3d; Supplementary Table 15). Non-circulatory organs accounted for 253 of the 319 compact-history captures (79.3%) in eICU. In the earlier scheduled own-organ-history comparison, 43 of 122 newly captured episodes had a higher rank for the affected organ within the patient, with a median paired risk increase of 10.29 percentage points across the newly captured group (Supplementary Table 42).

### Adaptation and record retrieval recover different kinds of information loss

When 50% of historical record batches were withheld across three masking assignments in eICU, the standard compact model (D0) captured a mean of 180.0 episodes (175, 183, and 182; mean AUROC 0.7749), whereas training with incomplete records (Dm) increased mean capture to 211.0 (+31.0 events, +17.2%; 95% confidence interval 16.3 to 47.7; mean AUROC 0.8080; Fig. 4a; Supplementary Table 21). Adapted training also recovered full-history capture under 50% withholding (173.7 to 210.3) and 1–3 hour history truncation (Supplementary Table 21). Conditional generated-history comparisons improved log loss in some settings without establishing additional event capture over matched visible-sequence models; effects varied by generator and reader (Supplementary Tables 33–34).

**Figure 4.**
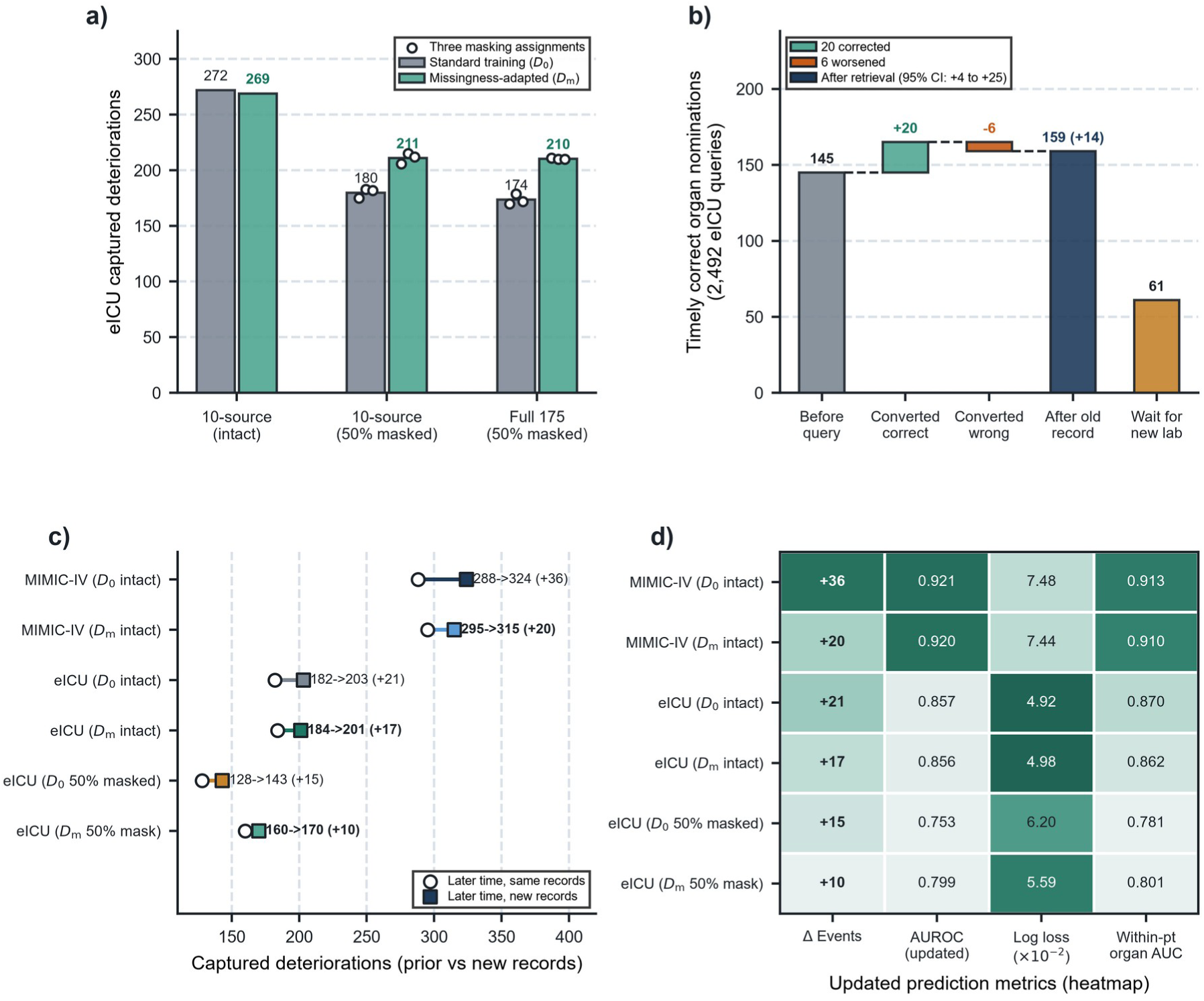
Record availability and additional information. a), eICU capture with standard training (D0) or training with incomplete records (Dm). Points identify three masking assignments and bars their means. With half the record batches withheld, capture with compact history increased from 180 to 211 (difference 31; 95% patient-bootstrap interval 16.3–47.7). b), Timely correct nominations across 2,492 requests: retrieval corrected 20 and worsened six choices, changing 145 to 159 (difference 14; 95% interval 4–25). Waiting for an actual new record produced 61 timely correct nominations among 139 replies. c), Clock advancement alone versus newly arrived measurement contents under intact or withheld history. d), Capture gain, AUROC, log loss and within-patient organ AUROC for the same future target. Waiting and retrieval use distinct information opportunities.

In arrival-time aligned evaluation across 2,492 eICU queries, immediately retrieving one existing historical laboratory group (collected 3–24 hours pre-index) increased timely correct six-hour organ nominations from 145 to 159 (+9.7%; net +14, patient 95% confidence interval 4 to 25; hospital interval 3.9 to 23.8), converting 20 incorrect nominations to correct and 6 in the opposite direction (Fig. 4b; Supplementary Tables 23–24). Waiting up to two hours for a newly collected laboratory test yielded 139 timely replies and 61 timely correct nominations.

### New measurements change the contribution of earlier history

For a fixed future outcome window, incorporating newly arrived measurements improved capture beyond advancing the clock alone: capture rose from 288 to 324 (+36) in MIMIC-IV and from 182 to 203 (+21) in eICU for D0 (295 to 315 and 184 to 201 for Dm; Fig. 4c,d; Supplementary Table 22). Under 50% history withholding in eICU, the adapted model before new records arrived (160 captures, AUROC 0.7831) already exceeded the unadapted model after new records arrived (143 captures, AUROC 0.7531), and reached 170 captures (AUROC 0.7993) once new records were incorporated (Fig. 4c,d).

To separate earlier history from new information, we compared models receiving the same 225 variables describing current observations, recording patterns and post-index measurements; one also received history from before index, kept unchanged. Overall eICU capture after index increased from 177 without earlier history to 214 with compact earlier history (Supplementary Table 25). With zero new measurements indicating low severity, compact earlier history improved eICU AUROC by +0.0258 (capture 185 to 195; paired Δ log loss −3.04 × 10−3); after one new measurement, the AUROC gain was +0.0158 (86 to 93). After >=2 low-severity measurements spanning >=1 hour, however, it overpredicted risk (1.47% compact and 1.74% full versus 1.38% observed; Fig. 5b) and reduced capture from 62 to 54 (ΔAUROC +0.0004; paired Δ log loss +1.30 × 10−3; Fig. 5a,c).

**Figure 5.**
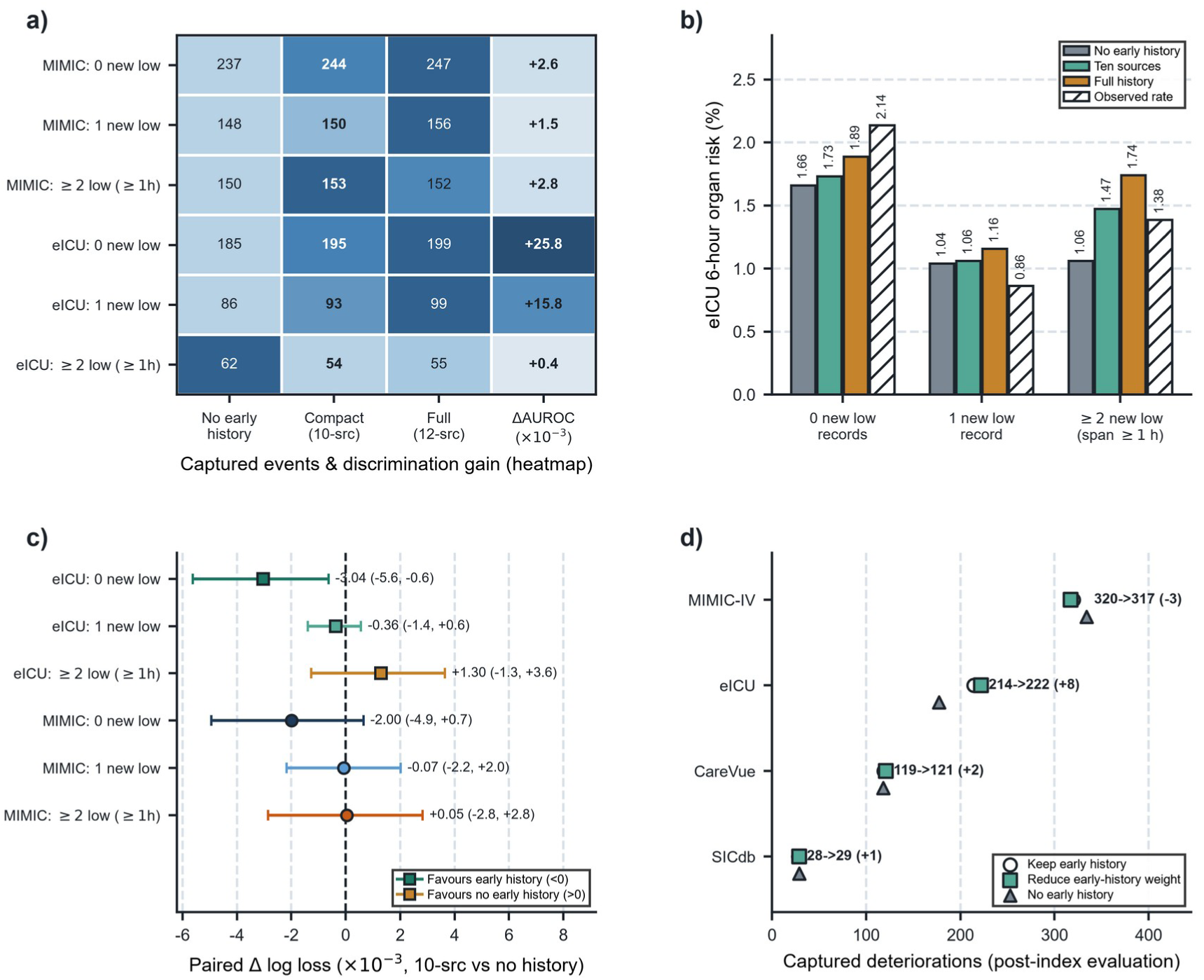
New measurements change the value of earlier history. a), Captures and AUROC differences with no fixed earlier history, compact earlier history or full earlier history, stratified by distinct new low-severity measurements; repeated measurements span at least one hour. b), Predicted risk and observed event frequency in eICU, showing overprediction after repeated low-severity evidence. c), Paired log-loss differences for ten-source versus no fixed earlier history, with 95% patient-bootstrap intervals. d), Capture with fixed earlier history, the rule, or no fixed earlier history across four databases. The rule halves its weight after at least two low measurements spanning one hour and sets it to zero after at least three spanning three hours; a new abnormal measurement resets the sequence. These experiments share current and post-index information and use the nine-origin protocol.

We progressively reduced the weight of earlier history as new measurements accumulated: full weight at 0–1 low measurements, half at >=2 spanning >=1 hour, and zero at >=3 spanning >=3 hours. In eICU, capture after index increased from 214 to 222 (log loss 0.05196 to 0.05148). Capture changed from 119 to 121 in CareVue and from 28 to 29 in SICdb, and from 320 to 317 in MIMIC-IV (Fig. 5d; Supplementary Tables 25–26).

### Local calibration improves the comparability of organ risks

Under matched local calibration opportunities (Fig. 6a,b; Supplementary Tables 18–19), organ-specific affine calibration across 91 eICU evaluation hospitals (adapted on 23 disjoint hospitals) reduced compact-model log loss from 0.05159 to 0.05079 and lifted capture from 218 to 233. Organ-specific shrinkage reduced compact-model log loss from 0.09904 to 0.09284 in CareVue (captures 126 to 129) and from 0.09507 to 0.08556 in SICdb, lifting SICdb AUROC from 0.8742 to 0.8854 and capture from 46 to 63 (Fig. 6a). AUROC varied by organ in MIMIC-IV and eICU (Fig. 6b; Supplementary Table 15). Action-matched decision curves nominating at most one organ per patient quantified net benefit across 0.5%–20% thresholds in MIMIC-IV and eICU (eICU net benefit at 5% threshold: 0.01224 versus 0.00913 for current information; Fig. 6c,d; Supplementary Table 20). In the hourly workflow, local calibration changed SICdb capture from 49 to 72 (difference 23, 95% confidence interval 8 to 38), compared with 263 to 269 in the 91 eICU evaluation hospitals, 144 to 140 in CareVue, and 448 to 448 in MIMIC-IV (Supplementary Table 44).

**Figure 6.**
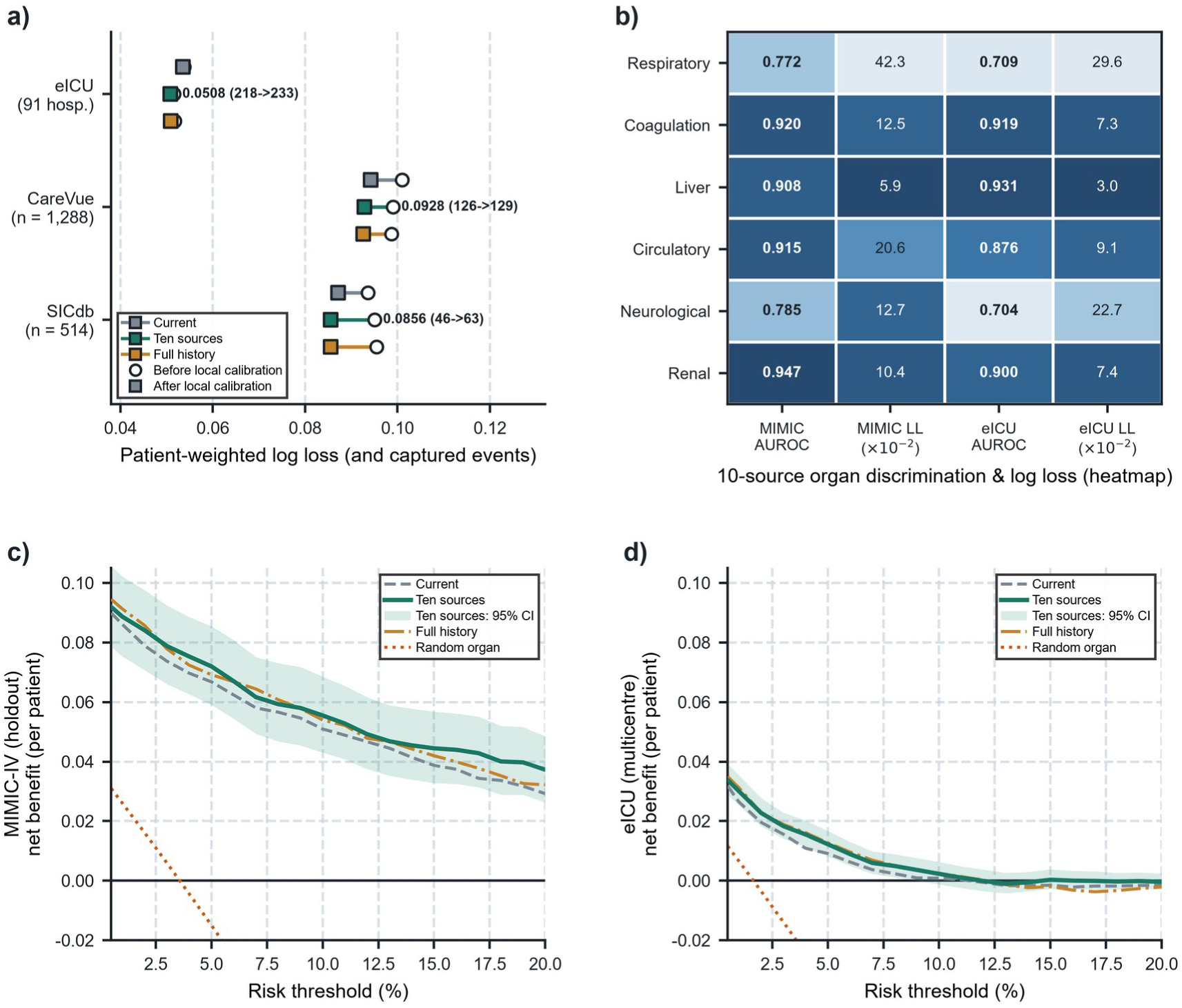
Local calibration and decision utility. a), Six-hour patient-weighted log loss before and after local calibration, with compact-history captures in parentheses. eICU evaluation used 91 hospitals separate from 23 adaptation hospitals (4,196 patients); CareVue and SICdb used patient-disjoint cross-fitting. b), Organ-specific AUROC and log loss for compact history in MIMIC-IV and eICU. c),d), Action-matched decision curves in MIMIC-IV and eICU at index, nominating at most one eligible organ per patient over risk thresholds of 0.5–20%. Shading gives pointwise 95% intervals from 1,200 patient-bootstrap samples for compact history; dotted lines show random eligible-organ nomination. AUROC, area under the receiver operating characteristic curve. Full history uses 175 inputs; compact history uses 163, including the same 67-column current baseline.

Recalculating GCS from observed components and refitting the models in MIMIC-IV preserved the benefit of compact history (MIMIC-IV capture 220 to 235; eICU evaluation with its native total-GCS definition, 210 to 260; Supplementary Fig. 4b; Supplementary Table 27). We also applied the models without retraining to pancreatitis-related cohorts of 1,043 MIMIC-IV and 618 eICU patients. In eICU, six-hour AUROC increased from 0.8012 to 0.8347 and capture from 20 to 29 (Supplementary Fig. 4a; Supplementary Tables 28 and 40). In the separate MIMIC-IV norepinephrine analysis, AUTOC for risk-based prioritisation increased from 20.02 to 22.75 µg/kg with compact history and to 24.62 µg/kg with full history; using current vasoactive treatment information to rank estimated treatment benefit gave an AUTOC of 23.33 µg/kg (Supplementary Fig. 4d; Supplementary Tables 35 and 38). In MIMIC-IV pancreatitis-related patients, 24-hour capture increased from 245 to 256 (Supplementary Table 28). The norepinephrine comparison used the same 813 first opportunities to reduce treatment. Adding history did not establish an advantage over the shared treatment-context comparator for ranking treatment benefit (Supplementary Tables 35 and 38).

## Discussion

Earlier organ history helped prioritise reassessment when current scores indicated low severity. At the same hourly alert budget, compact history increased six-hour event capture in eICU (+30.2%), CareVue (+34.6%) and SICdb (+81.5%). Counting events brought to attention within a fixed workload connects prediction to a specific clinical task.^12^ Patients with the same currently active organ may have reached that state after recovery from multiorgan involvement or through direct onset. Their histories help identify which other, currently low-severity organ deserves reassessment. This extends the question from a patient’s overall recovery pathway to the organ that may deteriorate next.

Selecting record sources addresses a practical problem that counting model variables alone misses. Established methods balance prediction, simplicity and interpretability, including the cost shared by variables obtained from the same source.^11,13,14^ Work in omics has similarly sought signatures that transfer between measurement systems.^15^ Here, each derived organ state was included only when its required records were available, and combinations were selected for the gains they retained in both development databases. Respiratory state, for example, requires arterial blood gases and FiO2; renal state requires creatinine and timed urine output. A single derived variable can therefore require several sources. ^10,11^ Ten sources retained substantial predictive benefit, while six sources offered an alternative with fewer records. The latter included lactate instead of the five additional sources used by the compact model. Reducing redundant variables and reducing the records needed to calculate them are different steps. Cross-database mapping is another constraint: training was restricted to variables that could be represented in both databases. The full 175-variable model consequently reflects both predictive selection and measurement compatibility, with unmapped variables excluded before the final comparison of source combinations.

Similar capture totals can cover different patients and organs. In eICU, compact and full history captured 319 and 314 events, respectively, but full history uniquely captured more renal events, while compact history uniquely captured more coagulation, respiratory and circulatory events. These differences matter when choosing which records to retrieve. The separate organ-ranking analysis also showed that history could change the organ selected for a patient’s reassessment (Supplementary Table 42). The choice between six and ten sources was sensitive to the size of the development gain: the brief model retained 9 of the 12 additional events captured by full history, so one more capture would have exceeded the 80% requirement. These results describe the trade-off between information requirements and event coverage across the available combinations.

The usefulness of history depended on what was available and what arrived next. Training with incomplete records recovered 31 eICU events when half the historical batches were withheld. Retrieving one existing laboratory result corrected 20 organ choices without waiting for a new sample. After two or more new measurements consistent with low severity over at least one hour, however, earlier history overpredicted risk and its discrimination gain diminished. This prompted the rule that reduced its weight as new evidence accumulated. Choosing which record to retrieve also relates to work on explanation-guided information acquisition.^16^ Some methods for generating missing history improved probability accuracy but did not clearly increase capture over models using the available records (Supplementary Tables 33–34). Training on incomplete records helps the model use what is visible, retrieval supplies an actual observation, and generation estimates missing information. Their value depends on whether the missing information can be recovered from the chart or requires a new measurement.

New measurements and elapsed time provided different information. Reusing a carried-forward normal value adds no new observation, whereas repeated measurements consistent with low severity can change estimated deterioration risk. When six organ risks compete for one alert, differences in organ event rates between hospitals can also change which organ is selected, even if ranking within each organ is preserved. Local calibration improved this allocation and substantially increased capture in SICdb.^17^ No tested threshold met the adjusted criteria for lower-priority reassessment at 1%, 2% or 5% failure targets (Supplementary Table 41). The model therefore supports prioritising review rather than deciding when monitoring can stop.

Routine ICU records reflect both illness and clinical decisions about when to measure it.^18,19^ Historical gains persisted when the 67-variable baseline included measurement timing and frequency and when the current-information comparison was expanded to 91 variables. In 26 catheter-removal episodes from 25 patients, urine collections decreased by 1.61 per six observed ICU hours after removal (95% confidence interval −2.25 to −0.96; Supplementary Table 43). This change in recording frequency explains why the absence of a new record should not itself count as evidence of recovery. Blood counts also vary with age and sex in community-dwelling older adults.^20^

Outcomes were defined from routine records using three consecutive hourly states and were not prospectively adjudicated. eICU informed model selection; CareVue and SICdb assessed performance after selection, with CareVue representing an earlier recording system at the MIMIC institution. Source counts describe data requirements; clinician time and financial costs were not measured. The rule for reducing the weight of earlier history had small external capture differences with intervals spanning zero. Estimates of treatment prioritisation rely on confounder adjustment and treatment overlap. Prospective evaluation should first assess alerts without affecting care, then test whether they prompt appropriate reassessment and improve management and outcomes. This should include the time required to obtain each record source.

Much of the predictive value of organ history could be retained with fewer record sources when selection accounted for the records required by derived variables. Compact history and the brief alternative support six-hour organ reassessment, while training with incomplete records, retrieval of existing results and local calibration address common differences in what information is available. New measurements can then change how much earlier history remains useful.

## Methods

### Cohorts and time-aligned organ states

Sepsis cohorts comprised MIMIC-IV v3.1 (12,905 adults), eICU-CRD v2.0 (4,983; 114 hospitals), MIMIC-III v1.4 CareVue (1,288) and SICdb v1.0.8 (514); pancreatitis was reserved for transfer testing.^21–29^ Sepsis ascertainment followed Sepsis-3 criteria in MIMIC-IV (suspected infection paired with rolling SOFA >=2), antimicrobial-treated infection with qualifying SOFA >=2 in eICU, antimicrobial-supported coded sepsis in CareVue, and recorded sepsis diagnosis in SICdb. Within each candidate observation period, we identified the first structural transition from two consecutive hours with either zero or >=2 active organs to three consecutive hours with the same single active organ. Index (t0) was the third single-organ hour. The selected transition was then checked for eligibility; a later transition was not substituted if the first failed. In MIMIC-IV and eICU, all five defining hours had to satisfy the sepsis information-time eligibility criterion. Across cohorts, t0 had to precede ICU discharge or death, with at least one other organ observed below the activity threshold.

Activity meant SOFA component >=2 in the six organ systems. Features and labels respected measurement times and, where recorded, availability times. Respiratory scoring paired arterial blood gas pO2 with FiO2 recorded within the preceding 2 h; vasoactive exposure followed documented infusion intervals; and neurological scoring used total GCS. In the MIMIC-IV main branch, recorded GCS components were carried forward for up to 6 h. If any component was present, missing companion components received normal values; an endotracheal or tracheostomy verbal code of zero mapped to total GCS 15 under the public concept convention. Fully unobserved hours remained missing. An observed-component sensitivity analysis propagated uncertainty rather than applying these normal defaults. Other lookback validity windows were 6 h for vital signs and FiO2, 12 h for blood gases and lactate, and 24 h for platelets, bilirubin, and creatinine; urine output required 22–30 h of covered intervals (none >6 h) normalised to 24 h.

### Prediction task and matched information comparisons

Eligible organs had observed SOFA components 0–1. Deterioration required three consecutive active hourly states (SOFA >=2), with onset 1–4 h after prediction and confirmation within 6 h before death or ICU exit. Valid carried measurements could support adjacent states; unknown did not establish recovery. Each distinct patient-organ-onset episode was credited at most once; 12- and 24-h extensions used identical confirmation rules.

Six LightGBM predictors used depth 5, 31 leaves, learning rate 0.06, minimum leaf 20, L2 penalty 5, feature fraction 0.9, and up to 400 trees with 30-round early stopping; eligible windows had equal total patient weight.^30^ MIMIC-IV was partitioned into patient-disjoint fitting (n = 8,990), early-stopping (n = 390), temporary calibration (n = 293), selection (n = 294), final calibration (n = 978), and held-out evaluation (n = 1,960) splits. The baseline with 67 variables comprised current values, states and recording-process timing/coverage. Controls used 91 current columns, recent measurements (<=3 h), 25%/50%/100% fitting samples, and current-value (50), recording-process (+17), numerical-history (+90) and state-history (+18) components.

### Clinical feature compression

Upstream selection grouped inputs by source-record type, using patient-weighted TreeSHAP contributions normalised within organ and averaged across the six outcomes. Groups were assigned equal costs; candidate budgets of 45, 32, 23, 17, 12 and 9 record groups were evaluated, and refitted group budgets were compared against at least 98% reference capture and no more than 1% increases in log loss and Brier score. The retained 17-group representation contained 1,492 columns. To align source training with eICU availability, 912 unmapped columns were masked; 28 constant or unused variables were then removed, leaving 552. This step limited training to variables that could be represented in both databases. After group selection and cross-database mapping, pruning individual variables recomputed TreeSHAP after each refit and removed approximately 10% of optional inputs while retaining all six current organ states. The 27 evaluated representations ranged from 552 to 38 inputs; 175 was retained as the reference model (Supplementary Table 12). The source-level search then used its 67 shared current and 108 historical columns from 12 additional sources. A thirteenth registered switch, pain assessment, was already in the baseline. Historical column j was admitted only when its upstream dependencies Dj were available in source subset S:

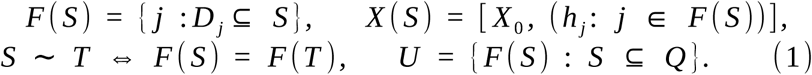

The 8,192 candidate combinations yielded 3,072 distinct inputs. Three-fold patient-disjoint MIMIC-IV evaluation identified 33 frontier candidates for eICU. Retention used patient-weighted log loss L and captured episodes C; subscripts 0, S and F denote current information, subset S and full history:

Q denotes registered switches; U removes equivalent inputs. N counts patients, I their eligible organ-windows and n their number. Ratios require positive reference gains. Ties favour greater minimum retention, fewer historical columns, then the registered mask.

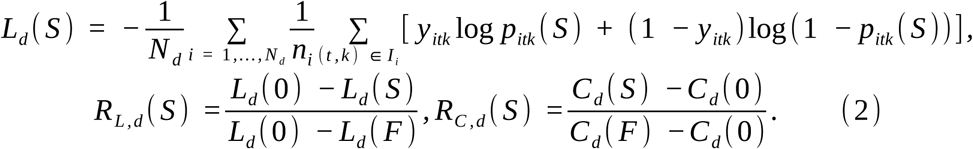

Within the 33 source-frontier candidates P evaluated in eICU, selection minimised historical-source count subject to retaining at least 80% of both gains in each development database:

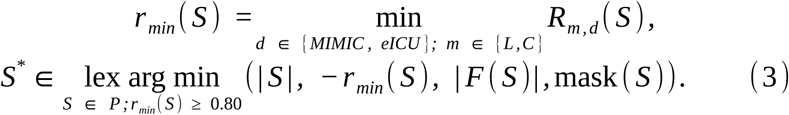

We selected compact history from ten sources (96 historical variables, 163 in total) and retained brief history from six (63 historical, 130 in total) as an alternative requiring fewer records. Signed TreeSHAP values on the log-odds scale were grouped by the records needed to calculate each variable, counting each coupled organ state once.^31^ Brief history preserved much of the eICU gain but did not meet the joint 80% criterion (Supplementary Table 14).

### Reassessment allocation and probability evaluation

The main workflow refreshed risk hourly through hour 23, nominated the highest-risk eligible organ, and imposed a 4-h patient cooldown at 0.50 alerts per patient. Mechanistic experiments retained nine origins and 6-h cooldown. Probability accuracy was assessed via patient-weighted log loss, Brier score, AUROC, and within-patient organ AUROC.^32^ Action-matched decision curves evaluated at most one nominated organ per patient at index across thresholds p in [0.5%, 20%], weighting false-positive nominations by p/(1-p).^33^ Local calibration compared identity, affine, shrinkage of organ intercepts with a shared slope, beta^34^, and isotonic maps selected by patient-weighted log loss (using a 23-to-91 hospital split in eICU and patient-disjoint cross-validation in CareVue and SICdb).

Hourly nomination, patient cooldown and episode-level capture were defined by:

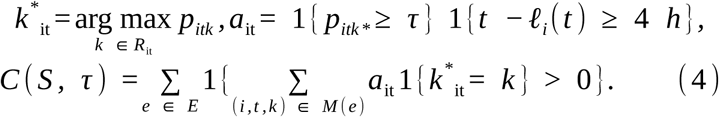

R denotes eligible organs, ℓ the last accepted alert time, τ the workload threshold, and M(e) nominations matching episode e. Labels enter episode scoring, not nomination.

### Withheld records, retrieval and new evidence

In three assignments, we withheld 50% of historical record batches throughout follow-up. We compared standard training (D0) with training on paired complete and incomplete histories (Dm), including indicators of record availability. For retrieval, we hid groups of laboratory records collected 3–24 h before index and evaluated 2,492 eICU requests. Each request could reveal at most one existing group, and timely six-hour organ choices were counted only after results were available; the comparison allowed waiting <=2 h for a new test. Update experiments compared elapsed time and new measurements for the same future outcome, using complete histories and 50% withholding. To study earlier history, two models received the same 225 variables describing current and post-index information; one also received history recorded strictly before index and kept unchanged. We stratified by 0, 1 or >=2 new measurements supporting low severity over >=1 h. The rule gave earlier history weight 1 initially, 0.5 after >=2 measurements spanning >=1 h, and 0 after >=3 spanning >=3 h.

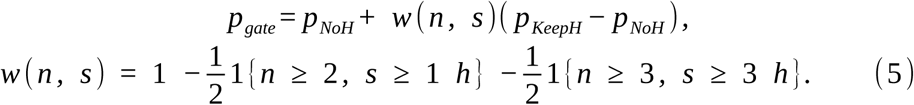

Both predictions share current and new information; n counts distinct new low-severity measurements since the last new abnormality, and s is their time span. Carried values do not increment n.

### Supporting comparisons

Within-patient localisation conditioned on the eligible organ set and the observed number of organs deteriorating within the prediction horizon. The latter was an offline scoring condition, not a predictor available at deployment. Sequential access began with six sources and allowed two additional sources or stopping, using source out-of-fold correction targets and equal request budgets. Scheduling held predictions fixed; a future-label dynamic-programming bound measured headroom only. Conditional-history comparisons used two patient-disjoint donor banks, 32 Extra-Trees per bank (depth 10, minimum leaf 32), opposite-bank training queries, 16-dimensional SVD or patient-weighted PCA targets, and matched 18-summary readers.^35^ Additional readers included conditional tree diffusion and Schrödinger-bridge imputation.^36,37^

In MIMIC-IV, observed-range GCS assigned missing eye, motor and verbal components ranges of 1–4, 1 –6 and 1–5. Lower bounds ≥13 established low severity, upper bounds ≤12 activity, and overlaps remained unknown; inputs and labels were rebuilt and models refitted among retained qualifying entries. eICU evaluation retained its native total-GCS definition. Pancreatitis-related cohorts used diagnoses or marked pancreatic enzymes, excluded sepsis-cohort overlaps, and reconstructed the same organ task without an infection gate; features, weights and calibration transferred unchanged.

NE comparisons fixed 813 first opportunities, indices, exposure definitions and strategies. Shared treatment-context inputs numbered 151; history increased totals to 214, 247 or 259. Patient-separated augmented effects trained benefit rankings; prognostic rankings prioritised lower six-hour hypotension-or-death risk.^38^ AUTOC^39^ and 20% initiation capacity (162 patients) used fixed fitted models with 500 paired patient draws. Lower-priority rules tested 19 thresholds per representation against 1%, 2% and 5% risk targets using Bonferroni-adjusted binomial tests over 76 combinations, plus a hospital-level bounded-mean test; death and unresolved follow-up counted as failures.

Nominated-organ calibration used patient cross-fitting in each complete cohort. Matched missingness controls fitted separate maps within the same family on fully observed adaptation data before transfer to masked views. Cross-routing retained the same patients and assessment times while changing only organ nomination.

Clinical interpretation grouped raw TreeSHAP contributions at first alerts into current severity, target/other-organ history and recording timing, and examined organ-ranking margins. Urine collections were compared in paired six-hour windows around catheter removal, excluding irrigation and unusable times, standardising to observed ICU time and using 2,000 patient resamples.

### Statistics and reproducibility

Reporting was checked against TRIPOD+AI.^40^ Uncertainty was quantified via paired patient-cluster bootstrap resampling (plus hospital-cluster resampling in eICU), rebuilding chronological alert allocation, cooldown suppression, and episode deduplication within each draw (500 draws for hourly replay; 1,200 draws for retention, missingness, retrieval, and decision-curve analyses). All databases provided deidentified records under approved institutional governance (Beth Israel Deaconess Medical Center/MIT for MIMIC; Salzburg EK1115/2021 with waiver of consent for SICdb).

Generative AI assisted language editing and figure-layout code preparation; empirical results came from the study analyses.

## Data Availability

The clinical data used in this study are available from PhysioNet through MIMIC-IV, the MIMIC-III Clinical Database (CareVue subset), the eICU Collaborative Research Database and the Salzburg Intensive Care database (SICdb), subject to the respective access requirements and data-use agreements. Access requires the applicable credentialing, training and permissions specified by the data providers. Restricted patient-level records and patient-level predictions are not redistributed by the authors. Aggregate results are reported in the manuscript and supplementary materials.

https://physionet.org/content/mimiciv/

https://physionet.org/content/mimiciii/1.4/

https://physionet.org/content/eicu-crd/2.0/

https://physionet.org/content/sicdb/

## Data availability

MIMIC-IV, eICU and the MIMIC-III CareVue subset are available through PhysioNet under their respective credentialing and data-use requirements. SICdb is available under its source access conditions. Patient-level records and predictions are not redistributed. Aggregate results are reported in the manuscript and Supplementary Information.

## Code availability

The analysis code is not publicly available and is not included with this preprint.

## Acknowledgements

This work was sponsored by the National Natural Science Fund of China (82373417) and Shanghai Pujiang Talents Program (24PJD013, 24PJA012). The funders had no role in study design, data analysis, interpretation or manuscript preparation.

## Author contributions

L.W.: Conceptualization, Methodology, Software, Data curation, Formal analysis, Validation, Visualization, Writing – original draft. J.S., M.V., X.L., Q.Z.: Investigation, Writing – review and editing. R.Z., J.J.Z., D.L., H.Z., H.M.Z., C.Z., L.Z., J.H.Z.: Investigation, Writing – review and editing. X.G., H.H., Y.S., S.C.: Supervision, Writing – review and editing.

