## Supplementary Information for "Compact clinical features capturing organ history for dynamic prediction of organ deterioration in sepsis"

### Supplementary Results

#### History adds information within patients and beyond simpler summaries

Supplementary Table 7 compares history of the target organ and derived organ states using the ten selected sources. The training-sample comparisons in Supplementary Tables 9–10 use target-organ history from the full model. Each experiment compares models using the same training procedure and current information. A separate analysis required a target-organ measurement within the preceding three hours at index (Supplementary Table 8); the measurement could be normal or abnormal.

Conditioning on each patient's eligible organs and event count, target-organ history reduced log loss by 0.1396 in MIMIC-IV and 0.1093 in eICU. Requiring new measurements supporting the subsequent event reduced the respective gains to 0.0134 and 0.0433, while their confidence intervals remained above zero (Supplementary Table 5). These comparisons localise historical information within the patient rather than relying only on between-patient risk differences.

Simple memory was useful but did not reproduce the full historical representation. In eICU, adding state duration increased capture from 219 to 247 episodes; 24 historical summaries captured 235, and adding duration to those summaries captured 256. Full history captured 282. Adding duration to full history instead yielded 262, illustrating that more expressions were not uniformly beneficial (Supplementary Table 11). Across fitting samples of 2,247, 4,495 and 8,990 patients, the corresponding captures using current information were 191, 202 and 219, versus 233, 243 and 282 with organ history (Supplementary Tables 9–10).

#### Compression changes organ coverage as well as total performance

Supplementary Tables 3 and 12 describe the record groups and reduction of model variables. Supplementary Tables 13–14 report the gains retained and the 33 candidate combinations evaluated in eICU. Candidate scores in MIMIC-IV were obtained by cross-validation during development; performance in the separate evaluation group is reported independently. Supplementary Table 16 gives paired capture differences under hourly reassessment.

Compact history increased six-hour AUROC over current information from 0.8979 to 0.9114 in MIMIC-IV and from 0.8418 to 0.8695 in eICU. The paired differences, probability scores and expanded-current comparisons are given in Supplementary Tables 6 and 39. Its gains were heterogeneous across organs (Supplementary Table 15). Under hourly reassessment, compact history shared 260 eICU captures with full history, added 59 and lost 54. Coagulation contributed 18 additions and 10 losses; renal events contributed nine additions and 25 losses. Full event intersections for all four databases are retained in Supplementary Table 17.

#### Access and timing determine the usefulness of new information

Across three assignments withholding half the historical records, training with incomplete data increased mean eICU capture from 180 to 211. Retrieving an existing record corrected 20 organ choices and worsened six, increasing timely correct choices from 145 to 159. Waiting for a new recorded result gave 61 timely correct choices, compared with 60 after the same delay without the result. Retrieval and waiting were evaluated as separate ways of obtaining information (Supplementary Tables 21–24).

Adding new measurements increased eICU capture from 182 to 203 when the future outcome window was held constant. With records persistently withheld, the model trained for incomplete data captured 160 events before new results and 170 after receiving them, compared with 128 for the original model without new results. Both training and access to new measurements contributed to the improvement.

#### Later evidence changes the contribution of fixed earlier history

In eICU, compact fixed history increased capture from 177 to 214 overall. After repeated new low-severity measurements spanning at least one hour, however, it captured 54 rather than 62 events and increased log loss from 0.06683 to 0.06813. In MIMIC-IV, overall capture was 320 with compact fixed history and 334 without it. Results differed across populations and groups of new measurements (Supplementary Table 25).

Reducing the weight of earlier history increased capture from 214 to 222 in eICU, from 119 to 121 in CareVue and from 28 to 29 in SICdb. The CareVue and SICdb intervals included zero. Omitting earlier history captured 118 in CareVue and 29 in SICdb and gave lower log loss than the weighting rule (Supplementary Table 26). The preferred use of earlier history therefore varied between datasets.

#### Calibration and recording context affect cross-system allocation

In SICdb, local organ calibration changed compact-history capture from 46 to 63 and full-history capture from 40 to 61 across the scheduled assessments. The calibrated current model captured 51, and the local organ prior captured 61. Matched probability estimates and calibration results are reported in Supplementary Tables 18–19. Decision curves retain the action of nominating one organ, with threshold-specific probability and workload trade-offs rather than treating any event in a nominated patient as a correct organ choice (Supplementary Table 20).

The total SOFA score, its components and the recorded-state comparator are evaluated on the same scheduled assessments in Supplementary Table 32. The total-score model uses a source organ prior when the total is unavailable, so that comparison retains the contribution of the fallback rather than treating a missing score as a measured normal result.

In eICU, adding the 18 organ-state expressions to the current and numerical-history inputs reduced log loss by 0.001748 (95% confidence interval 0.001095–0.002610) and increased capture from 240 to 282, a difference of 42 (17–68). Adding recording-process expressions to current and numerical history instead increased log loss by 0.000384 (0.000053–0.000779). The components therefore provided different conditional information across settings (Supplementary Table 31).

#### Sequential access and alternative history methods

The two-step access policy captured 300 eICU events, compared with 312 using brief history and 319 using compact history. Risk and uncertainty priorities captured 297 and 306. The selected opportunity-cost scheduling rule changed compact-history capture by +3 in MIMIC-IV, +1 in eICU, −2 in CareVue and −1 in SICdb; every paired capture interval included zero (Supplementary Tables 29–30). We therefore used the simpler risk ordering for the main analysis.

Several methods for estimating missing history improved log loss compared with models using available sequences, without a clear increase in capture. Averaging predicted probabilities reduced log loss by 0.000959; logistic combination gave higher log loss than this average. Schrödinger-bridge imputation and the recurrent model using generated history also gave less favourable results (Supplementary Tables 33–34). Generation improved prediction of coarse historical types but increased four-block log loss. The difference between intact and permuted history was larger for the generator alone than for the final mixture (Supplementary Table 36).

#### Risk thresholds for lower-priority reassessment

None of the 36 evaluations across models, risk targets and testing settings selected a threshold meeting the prespecified criteria (Supplementary Table 41). We therefore used prioritisation at an equal alert budget rather than defining a group in which monitoring could be reduced.

#### Neurological ascertainment

With the observed-component GCS definition in retained MIMIC-IV entries, compact history increased capture from 220 to 235. Evaluation using the unchanged native eICU total-GCS definition yielded 260 versus 210 captures (Supplementary Table 27).

#### Transfer to pancreatitis-related cohorts

In the pancreatitis-related eICU cohort, applying the compact model without retraining increased six-hour AUROC from 0.8012 to 0.8347 and reduced log loss from 0.04810 to 0.04629. Capture increased from 20 to 29. In MIMIC-IV, models using current information and compact history both captured 189 events; full history captured 190. Supplementary Tables 28 and 40 report longer prediction windows and paired intervals.

#### Organ history in norepinephrine prioritisation

In the separate norepinephrine task, matched treatment-context information alone achieved a benefit-ranking AUTOC of 23.33, compared with 17.61, 18.40 and 14.84 after adding brief, compact and full organ history. At 20% capacity, their estimated recorded-exposure reductions were 18.44, 14.02, 16.86 and 11.27 micrograms/kg, respectively. Risk rankings gave a different pattern, with AUTOC values of 20.02, 22.75 and 24.62 for current, compact and full information. The paired contrasts did not establish an additional prioritisation benefit from the organ-history representations (Supplementary Tables 35 and 38).

#### Lead time

Lead times are summarised across captured deterioration episodes. Under the scheduled-assessment protocol, compact-history median onset lead times in eICU were two, five and seven hours for the six-, twelve- and twenty-four-hour prediction windows. Confirmation followed onset by two hours under the three-position rule. These scheduled-assessment medians are reported separately from the hourly workflow (Supplementary Table 37).

#### Clinical interpretation and recording context

In the scheduled comparison using each organ's own history, history added 122 events in 117 patients and lost 59. Among the additions, the affected organ ranked higher in 43. After grouping SHAP values by information type, target-organ history had the largest absolute contribution in 109 additions and contributed most to the change in ranking margin in 37 improved choices. This analysis distinguishes changes in the risk score from changes in which organ is prioritised (Supplementary Table 42).

Recording frequency changed after support removal. Across 26 urinary-catheter removal episodes in 25 patients, mean urine collection counts fell from 3.23 to 1.58 in paired six-hour windows. After standardising to observed ICU time, the difference was −1.61 collections per six hours (95% CI −2.25 to −0.96; Supplementary Table 43). Fewer recorded collections after catheter removal do not by themselves indicate physiological recovery.

#### Calibration and selection diagnostics

In the hourly workflow, local calibration changed capture with compact history from 49 to 72 in SICdb, 144 to 140 in CareVue and 263 to 269 in the hospital-held-out eICU evaluation (Supplementary Table 44). Under a matched calibration family, training with incomplete records retained an average advantage of 14.67 captures (patient 95% CI 1.83 to 26.84; hospital CI 3.20 to 23.95; Supplementary Table 45).

Among eICU organs selected at index above 5% risk, predictions before local calibration averaged 13.04%, compared with 9.32% observed. After organ-specific calibration, the corresponding values were 10.38% and 9.61% among the nominations selected by that model (Supplementary Table 46). At scheduled assessments, current information captured 219 events; changing only the organ choice captured 223, changing patient and assessment-time selection captured 237, and using history for both captured 272 (Supplementary Table 47).

#### Supplementary Figure 1 | Baseline risk stratification by prior organ involvement, within-patient localisation, and organ-specific discrimination.

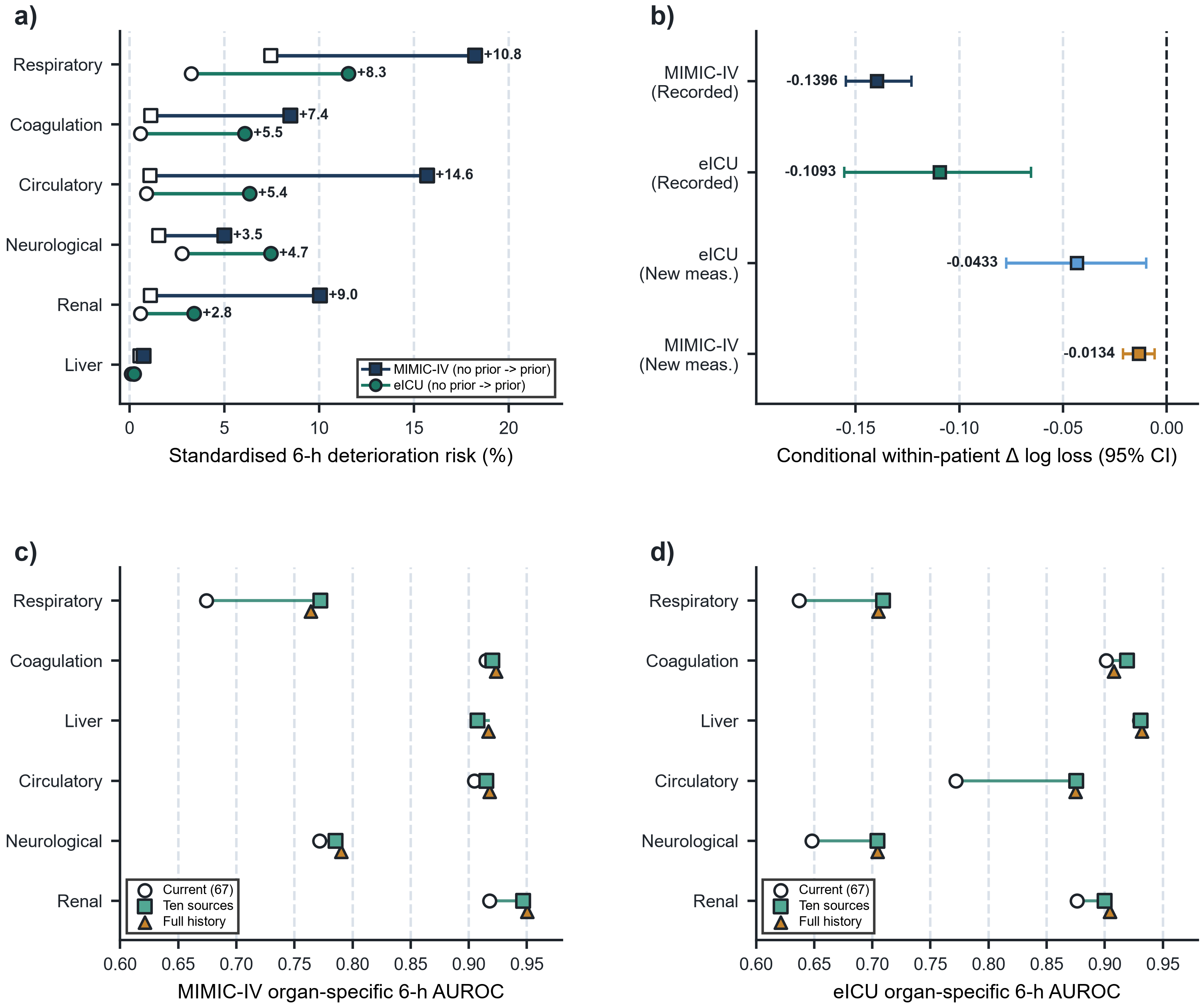

a), Standardised six-hour risk with versus without prior involvement of the same organ, adjusted for the active index organ and entry route. b), Conditional within-patient log-loss differences after adding target-organ history, with 95% bootstrap intervals; patients have both event and non-event organs. c),d), Organ-specific AUROC in MIMIC-IV and eICU for current information, compact history and full history (67, 163 and 175 inputs). Filled and open markers in a) indicate prior involvement and no prior involvement, respectively. AUROC, area under the receiver operating characteristic curve. Supplementary Tables 4–5 and 15.

#### Supplementary Figure 2 | Scalar feature-pruning trajectory, training sample-size scaling, and comparison with simple summary memory.

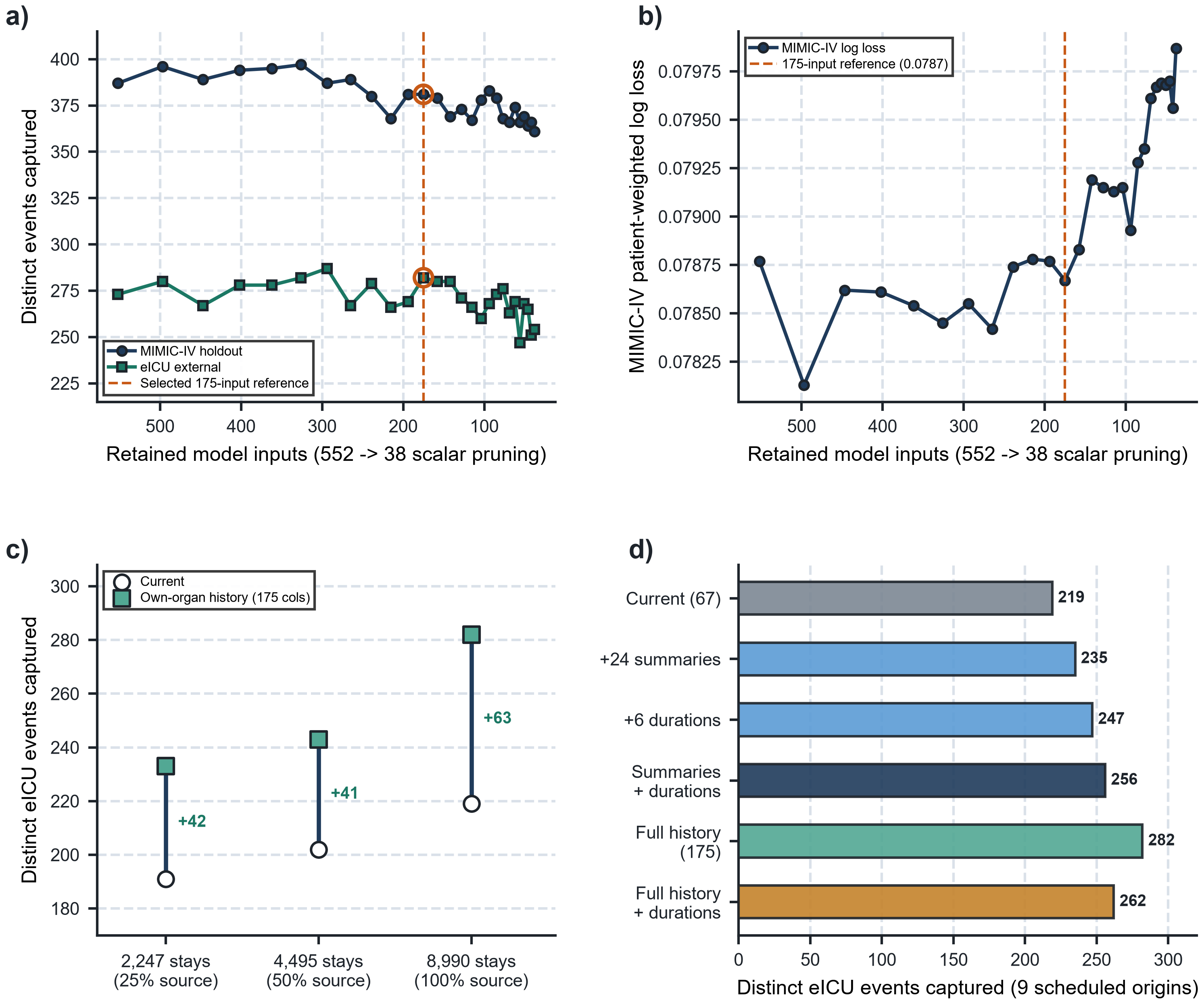

a),b), Six-hour captures in MIMIC-IV and eICU and source log loss along the 552-to-38-column pruning path; the highlighted 175-variable reference model reduces derived inputs by 68.3%. c), eICU capture with nested training samples of 2,247, 4,495 and 8,990 patients, comparing current information with target-organ history. d), eICU capture with current information, 24 historical summaries, six state-duration variables, their combination, full history, and full history plus duration. These comparisons use nine scheduled assessment origins and six-hour patient suppression. Supplementary Tables 9–12.

#### Supplementary Figure 3 | Multi-horizon prediction performance (6, 12, and 24 hours), feature-group decomposition, and warning lead times.

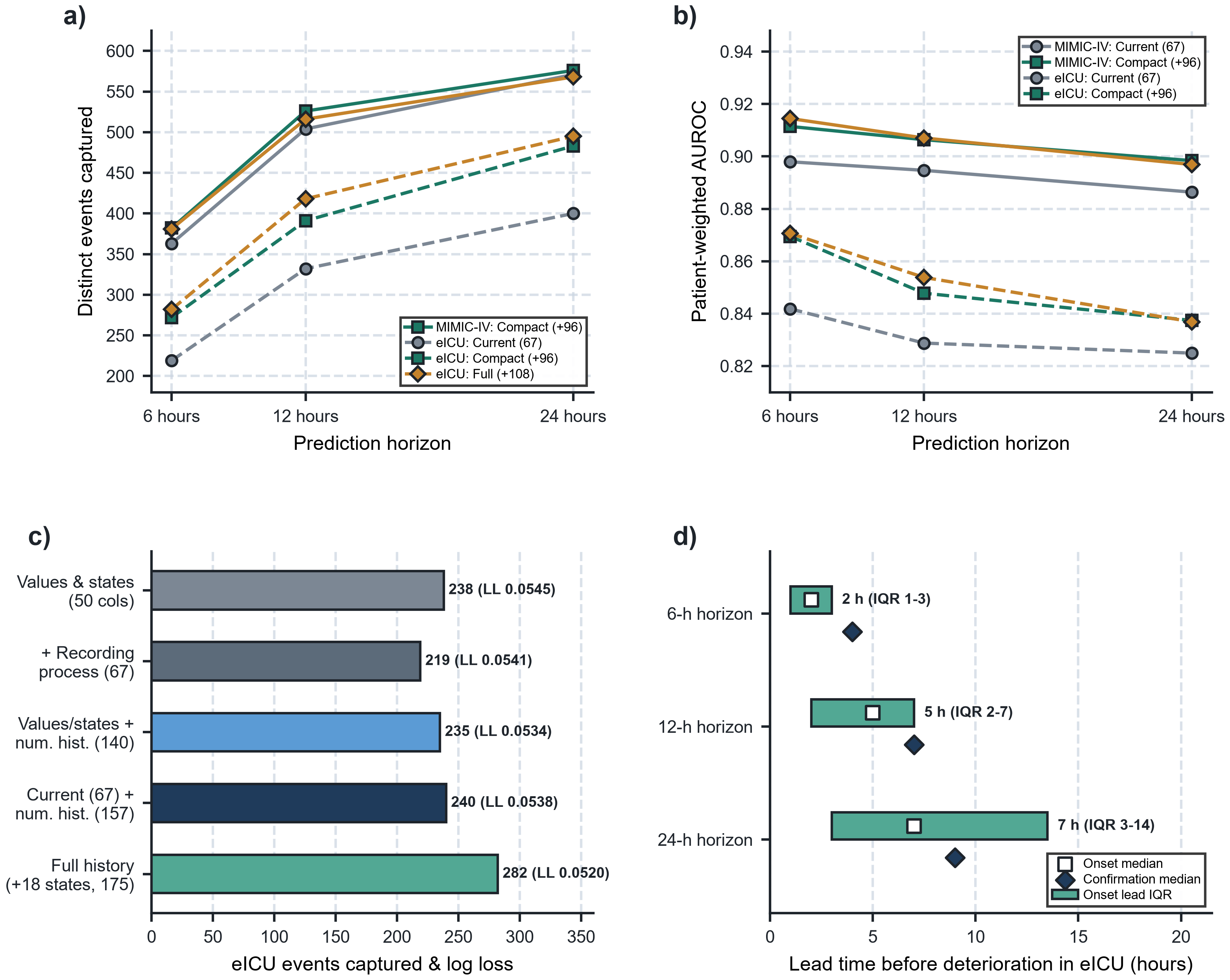

a),b), Captures and AUROC at 6, 12 and 24 hours in MIMIC-IV and eICU for current information, compact history and full history. c), eICU capture and log loss with current values/states (50 columns), recording-process inputs (67), numerical history (140 or 157), and derived state history (175). d), eICU onset lead-time medians and interquartile ranges (squares and bars) and confirmation medians (diamonds) for compact history. Results use the scheduled-assessment protocol, not the primary hourly workflow. AUROC, area under the receiver operating characteristic curve. Supplementary Tables 6, 31 and 37.

#### Supplementary Figure 4 | Cross-disease transfer to acute pancreatitis, strict neurological scoring sensitivity, active source querying, and norepinephrine prioritisation.

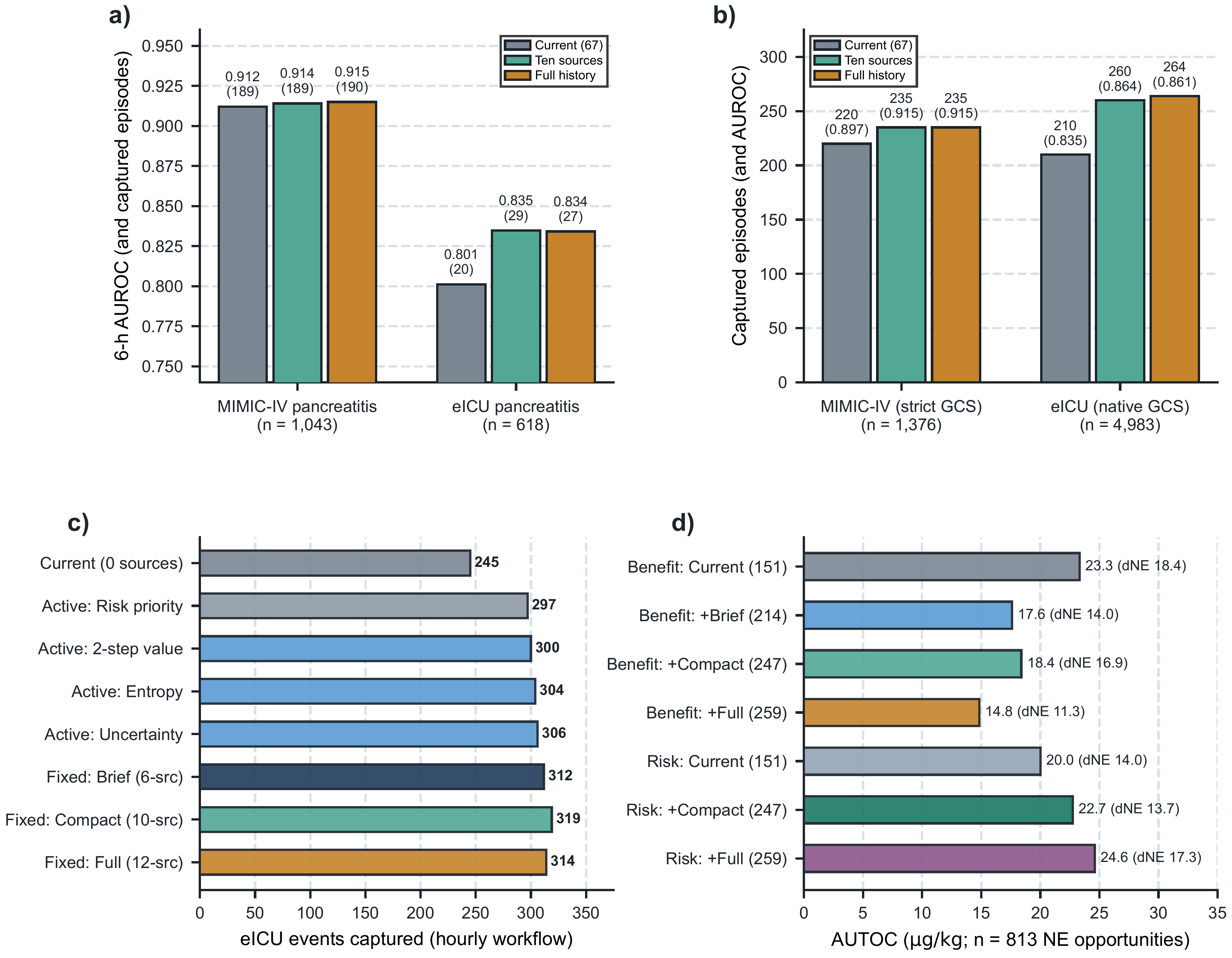

a), Sepsis models applied without retraining to pancreatitis-related MIMIC-IV (1,043 patients) and eICU (618), showing AUROC with captures in parentheses. b), Observed-component Glasgow Coma Scale sensitivity in retained MIMIC-IV entries (1,376), with evaluation using unchanged native total-GCS labels in eICU (4,983); captures are shown with AUROC in parentheses. c), eICU capture for sequential requests of up to two additional sources from the six-source baseline versus fixed representations. d), AUTOC and recorded NE-exposure reductions for matched representations at 813 MIMIC-IV norepinephrine decision opportunities. AUROC, area under the receiver operating characteristic curve; AUTOC, area under the targeting operator characteristic curve; NE, norepinephrine. Supplementary Tables 27–29, 35, 38 and 40.

#### Supplementary Table 1 Patient characteristics and follow-up

| **Database** | **n** | **Age, median (IQR)** | **Men, n (%)** | **Index to ICU exit, median h** | **Deaths by 24 / 48 h** |
| --- | --- | --- | --- | --- | --- |
| MIMIC-IV | 12,905 | 69 (58–79) | 7,497 (58.1) | 65.1 | 373 / 635 |
| eICU | 4,983 | 67 (57–78) | 2,713 (54.4) | 58.2 | 118 / 195 |
| CareVue | 1,288 | 70.6 (57.8–80.6) | 698 (54.2) | 84.96 | 44 / 85 |
| SICdb | 514 | 70 (60–75) | 319 (62.1) | 99.5 | 10 / 15 |

The four sepsis cohorts are shown, covering the full eligible sepsis populations. In CareVue, index-to-exit IQR was 38.83–241.80 h; deaths by 24 and 48 h were 3.42% and 6.60%. At index, 77, 434, 553, 145 and 79 CareVue patients had six, five, four, three and two observed organ states, respectively. IQR, interquartile range.

#### Supplementary Table 2 MIMIC-IV development and evaluation roles

| **Role** | **Patients** | **Prediction times** | **Eligible organ windows** | **Positive windows** |
| --- | --- | --- | --- | --- |
| Source fitting | 8,990 | 72,973 | 259,748 | 6,689 |
| Source auxiliary development | 977 | 7,961 | 28,527 | 682 |
| Final source calibration | 978 | 7,977 | 28,386 | 705 |
| Source evaluation | 1,960 | 15,901 | 56,941 | 1,555 |
| eICU development and selection | 4,983 | 39,619 | 124,484 | 1,455 |

All roles are patient-disjoint. Auxiliary development comprises 390 stopping, 293 temporary-calibration and 294 selection patients. Positive windows count organ assessments rather than distinct episodes.

#### Supplementary Table 3 Clinical record groups in the full reference

| **Record group** | **Source table** | **Recorded identifier** | **Columns** |
| --- | --- | --- | --- |
| Non-invasive systolic blood pressure | chartevents | 220179 | 16 |
| Categorical chart context | chartevents | 223791 | 3 |
| Glasgow Coma Scale verbal response | chartevents | 223900 | 11 |
| Bilirubin chart record | chartevents | 225690 | 7 |
| Platelet chart record | chartevents | 227457 | 10 |
| Norepinephrine record and recorded patient weight | inputevents | 221906 | 9 |
| Saline administration record | inputevents | 225158 | 12 |
| Lactate laboratory record | labevents | 50813 | 15 |
| Blood-gas oxygen laboratory record | labevents | 50821 | 18 |
| Creatinine laboratory record | labevents | 50912 | 14 |
| Platelet laboratory record | labevents | 51265 | 11 |
| Urinary catheter output record | outputevents | 226559 | 18 |
| Derived organ states and history | Derived state engine | Not applicable | 31 |

Counts denote derived model columns in the 175-variable reference model representation. Organ-state construction uses additional upstream measurements; record-source selection accounts for these shared dependencies.

#### Supplementary Table 4 Earlier organ involvement and subsequent six-hour risk

| **Database** | **Organ** | **Common-support patients** | **Prior / no prior activity risk (%)** | **Risk difference (95% CI), points** |
| --- | --- | --- | --- | --- |
| MIMIC-IV | Respiratory | 1,504 | 18.24 / 7.44 | 10.80 (7.03 to 14.56) |
| MIMIC-IV | Coagulation | 9,839 | 8.49 / 1.11 | 7.38 (4.29 to 10.46) |
| MIMIC-IV | Liver | 4,027 | 0.73 / 0.56 | 0.17 (−1.11 to 1.44) |
| MIMIC-IV | Cardiovascular | 11,359 | 15.69 / 1.07 | 14.62 (13.15 to 16.09) |
| MIMIC-IV | Neurological | 9,385 | 5.01 / 1.55 | 3.47 (2.10 to 4.83) |
| MIMIC-IV | Renal | 9,544 | 10.05 / 1.09 | 8.96 (4.93 to 12.98) |
| eICU | Respiratory | 360 | 11.55 / 3.26 | 8.28 (3.81 to 12.75) |
| eICU | Coagulation | 3,744 | 6.08 / 0.57 | 5.51 (3.88 to 7.14) |
| eICU | Liver | 2,448 | 0.24 / 0.09 | 0.15 (−0.33 to 0.64) |
| eICU | Cardiovascular | 3,412 | 6.33 / 0.89 | 5.44 (2.04 to 8.84) |
| eICU | Neurological | 2,352 | 7.44 / 2.77 | 4.67 (2.21 to 7.14) |
| eICU | Renal | 3,360 | 3.40 / 0.58 | 2.81 (1.04 to 4.59) |

Risks are standardised for the active organ and entry route. Intervals use patient clusters in MIMIC-IV and hospital clusters in eICU. Descriptive cohorts include the eligible patients, not only the prediction holdout.

#### Supplementary Table 5 Conditional localisation of future affected organs

| **Database** | **Outcome confirmation** | **Patients** | **Conditional log loss difference (95% CI)** |
| --- | --- | --- | --- |
| eICU | Recorded states | 787 | −0.1093 (−0.1554 to −0.0654) |
| MIMIC-IV | Recorded states | 3760 | −0.1396 (−0.1547 to −0.1231) |
| eICU | New supporting measurements | 682 | −0.0433 (−0.0774 to −0.0098) |
| MIMIC-IV | New supporting measurements | 1706 | −0.0134 (−0.0210 to −0.0058) |

Initial-assessment comparisons condition on the eligible organ set and total future event-organ count within a patient. Only patients with both event and non-event organs contribute. The comparison requiring new supporting measurements is reported separately.

#### Supplementary Table 6 Matched information comparisons across prediction horizons

| **Database** | **Current inputs** | **History inputs** | **Horizon (h)** | **AUROC** | **Log loss** | **Brier score** | **Events captured** |
| --- | --- | --- | --- | --- | --- | --- | --- |
| MIMIC-IV | 67 | 0 | 24 | 0.8864 | 0.18831 | 0.05444 | 571 |
| eICU | 67 | 0 | 24 | 0.8249 | 0.15603 | 0.04084 | 400 |
| MIMIC-IV | 67 | 0 | 12 | 0.8947 | 0.13372 | 0.03644 | 504 |
| eICU | 67 | 0 | 12 | 0.8288 | 0.10164 | 0.02418 | 332 |
| MIMIC-IV | 67 | 0 | 6 | 0.8979 | 0.08219 | 0.02056 | 363 |
| eICU | 67 | 0 | 6 | 0.8418 | 0.05406 | 0.01128 | 219 |
| MIMIC-IV | 91 | 0 | 6 | 0.8974 | 0.08220 | 0.02057 | 360 |
| eICU | 91 | 0 | 6 | 0.8420 | 0.05427 | 0.01138 | 205 |
| MIMIC-IV | 67 | 96 | 6 | 0.9114 | 0.07927 | 0.02020 | 382 |
| eICU | 67 | 96 | 6 | 0.8695 | 0.05173 | 0.01114 | 272 |
| MIMIC-IV | 67 | 108 | 6 | 0.9145 | 0.07867 | 0.02014 | 381 |
| eICU | 67 | 108 | 6 | 0.8706 | 0.05203 | 0.01130 | 282 |
| MIMIC-IV | 67 | 108 | 12 | 0.9070 | 0.12923 | 0.03570 | 516 |
| eICU | 67 | 108 | 12 | 0.8538 | 0.09790 | 0.02389 | 418 |
| MIMIC-IV | 67 | 108 | 24 | 0.8969 | 0.18361 | 0.05361 | 568 |
| eICU | 67 | 108 | 24 | 0.8369 | 0.15326 | 0.04067 | 495 |
| MIMIC-IV | 91 | 108 | 6 | 0.9132 | 0.07903 | 0.02020 | 373 |
| eICU | 91 | 108 | 6 | 0.8682 | 0.05241 | 0.01132 | 274 |
| MIMIC-IV | 67 | 96 | 12 | 0.9064 | 0.12925 | 0.03567 | 526 |
| eICU | 67 | 96 | 12 | 0.8478 | 0.09965 | 0.02429 | 391 |
| MIMIC-IV | 67 | 96 | 24 | 0.8984 | 0.18274 | 0.05357 | 576 |
| eICU | 67 | 96 | 24 | 0.8374 | 0.15432 | 0.04131 | 483 |
| MIMIC-IV | 91 | 96 | 6 | 0.9133 | 0.07888 | 0.02017 | 395 |
| eICU | 91 | 96 | 6 | 0.8662 | 0.05265 | 0.01137 | 270 |

Nine scheduled assessments with six-hour patient suppression; six-hour outcome unless otherwise indicated. The current model has zero historical variables; compact and full history add 96 and 108. The expanded current assessment has 91 variables.

#### Supplementary Table 7 Target-organ history and stronger current assessments

| **Database** | **Information** | **AUROC** | **Log loss** | **Brier score** | **Within-patient organ C** | **Events captured** |
| --- | --- | --- | --- | --- | --- | --- |
| MIMIC-IV | Target-organ history | 0.9130 | 0.07916 | 0.02022 | 0.9094 | 386 |
| MIMIC-IV | Organ-state history | 0.9081 | 0.07993 | 0.02028 | 0.9039 | 369 |
| MIMIC-IV | Target-organ history (25% fitting sample) | 0.9040 | 0.08202 | 0.02072 | 0.8931 | 349 |
| MIMIC-IV | Current information (25% fitting sample) | 0.8830 | 0.08503 | 0.02083 | 0.8696 | 349 |
| MIMIC-IV | Target-organ history (50% fitting sample) | 0.9106 | 0.07999 | 0.02035 | 0.9023 | 367 |
| MIMIC-IV | Current information (50% fitting sample) | 0.8920 | 0.08356 | 0.02074 | 0.8866 | 361 |
| MIMIC-IV | Expanded current + target-organ history | 0.9131 | 0.07899 | 0.02025 | 0.9063 | 379 |
| eICU | Target-organ history | 0.8707 | 0.05182 | 0.01118 | 0.8726 | 284 |
| eICU | Organ-state history | 0.8679 | 0.05257 | 0.01135 | 0.8722 | 253 |
| eICU | Target-organ history (25% fitting sample) | 0.8579 | 0.05403 | 0.01152 | 0.8628 | 231 |
| eICU | Current information (25% fitting sample) | 0.8267 | 0.05673 | 0.01187 | 0.8330 | 191 |
| eICU | Target-organ history (50% fitting sample) | 0.8628 | 0.05281 | 0.01128 | 0.8761 | 248 |
| eICU | Current information (50% fitting sample) | 0.8353 | 0.05480 | 0.01140 | 0.8412 | 202 |
| eICU | Expanded current + target-organ history | 0.8699 | 0.05249 | 0.01142 | 0.8740 | 277 |
| CareVue | Target-organ history | 0.8536 | 0.09734 | 0.02259 | 0.8642 | 117 |
| CareVue | Organ-state history | 0.8504 | 0.09829 | 0.02275 | 0.8562 | 115 |
| CareVue | Target-organ history (25% fitting sample) | 0.8372 | 0.10026 | 0.02287 | 0.8543 | 116 |
| CareVue | Current information (25% fitting sample) | 0.8337 | 0.10287 | 0.02313 | 0.8405 | 100 |
| CareVue | Target-organ history (50% fitting sample) | 0.8462 | 0.09826 | 0.02277 | 0.8616 | 119 |
| CareVue | Current information (50% fitting sample) | 0.8402 | 0.09955 | 0.02304 | 0.8471 | 103 |
| SICdb | Target-organ history | 0.8679 | 0.09395 | 0.02334 | 0.9160 | 30 |
| SICdb | Organ-state history | 0.8740 | 0.09407 | 0.02375 | 0.9172 | 41 |
| SICdb | Target-organ history (25% fitting sample) | 0.8549 | 0.09355 | 0.02264 | 0.9068 | 51 |
| SICdb | Current information (25% fitting sample) | 0.8492 | 0.09549 | 0.02281 | 0.9060 | 33 |
| SICdb | Target-organ history (50% fitting sample) | 0.8611 | 0.09381 | 0.02322 | 0.9108 | 38 |
| SICdb | Current information (50% fitting sample) | 0.8597 | 0.09365 | 0.02326 | 0.9104 | 28 |

Nine scheduled assessments with six-hour patient suppression; six-hour outcome unless otherwise indicated. Historical inputs are drawn from the ten selected sources and assigned to the target organ; the state-memory model retains the selected organ-state expressions. Current and complete-history references at 67 and 91 common inputs appear in the preceding table. Sample-size labels refer to fractions of the 8,990 fitting patients; current and target-organ history are matched at each size. This compact-source experiment differs from the full-reference training-sample comparisons in Supplementary Tables 9–10.

#### Supplementary Table 8 Historical information with a recent measurement at index

| **Database** | **Information** | **Patients** | **AUROC** | **Log loss** | **Brier score** | **Events captured** |
| --- | --- | --- | --- | --- | --- | --- |
| MIMIC-IV | Current information | 1,847 | 0.8742 | 0.13435 | 0.03548 | 137 |
| MIMIC-IV | Compact history | 1,847 | 0.8950 | 0.12671 | 0.03423 | 145 |
| MIMIC-IV | Full history | 1,847 | 0.8964 | 0.12712 | 0.03459 | 145 |
| MIMIC-IV | Own-organ history | 1,847 | 0.8918 | 0.12800 | 0.03457 | 143 |
| eICU | Current information | 3,838 | 0.7936 | 0.10107 | 0.02211 | 107 |
| eICU | Compact history | 3,838 | 0.8405 | 0.09327 | 0.02171 | 117 |
| eICU | Full history | 3,838 | 0.8390 | 0.09398 | 0.02185 | 119 |
| eICU | Own-organ history | 3,838 | 0.8407 | 0.09310 | 0.02168 | 114 |
| CareVue | Current information | 1,146 | 0.8380 | 0.14255 | 0.03448 | 68 |
| CareVue | Compact history | 1,146 | 0.8251 | 0.14036 | 0.03393 | 67 |
| CareVue | Full history | 1,146 | 0.8191 | 0.14385 | 0.03476 | 69 |
| CareVue | Own-organ history | 1,146 | 0.8276 | 0.14076 | 0.03434 | 69 |
| SICdb | Current information | 414 | 0.7825 | 0.32011 | 0.09205 | 43 |
| SICdb | Compact history | 414 | 0.7805 | 0.29085 | 0.08662 | 42 |
| SICdb | Full history | 414 | 0.7957 | 0.28284 | 0.08382 | 43 |
| SICdb | Own-organ history | 414 | 0.7634 | 0.29166 | 0.08642 | 43 |

Initial assessment with at least one numerical target-organ measurement no older than three hours. Recent does not require a normal value. The original common request budget is retained.

#### Supplementary Table 9 Historical value across source fitting-sample sizes

| **Information and fitting patients** | **Log loss** | **Brier** | **AUROC** | **Captures** |
| --- | --- | --- | --- | --- |
| Current and observation patterns, 2,247 fitting patients | 0.056631 | 0.011774 | 0.8267 | 191 |
| Own organ history, 2,247 fitting patients | 0.054475 | 0.011574 | 0.8563 | 233 |
| Current and observation patterns, 4,495 fitting patients | 0.054907 | 0.011406 | 0.8353 | 202 |
| Own organ history, 4,495 fitting patients | 0.053234 | 0.011346 | 0.8624 | 243 |
| Current and observation patterns, 8,990 fitting patients | 0.054162 | 0.011287 | 0.8418 | 219 |
| Own organ history, 8,990 fitting patients | 0.051991 | 0.011212 | 0.8706 | 282 |

Nested fitting samples retain target-organ history from the full 175-input reference, the same auxiliary roles and eICU evaluation patients. These are not the compact models in Supplementary Table 7. The fixed budget is 2,492 alerts in 4,983 patients. Feature selection is not repeated at each sample size. Intervals use paired patient samples; hospital contrasts are retained in the study source results. Positive capture differences and negative probability-score differences favour organ history.

#### Supplementary Table 10 Historical value across source fitting-sample sizes (continued)

| **Fitting patients** | **Capture difference (95% CI)** | **Log loss difference (95% CI)** | **Brier difference (95% CI)** |
| --- | --- | --- | --- |
| 2,247 | 42.00 (16.48 to 70.00) | −0.002156 (−0.002947 to −0.001401) | −0.000200 (−0.000323 to −0.000083) |
| 4,495 | 41.00 (18.00 to 68.00) | −0.001672 (−0.002447 to −0.000969) | −0.000061 (−0.000164 to 0.000032) |
| 8,990 | 63.00 (38.00 to 93.52) | −0.002171 (−0.003121 to −0.001347) | −0.000075 (−0.000207 to 0.000027) |

Nested fitting samples retain target-organ history from the full 175-input reference, the same auxiliary roles and eICU evaluation patients. These are not the compact models in Supplementary Table 7. The fixed budget is 2,492 alerts in 4,983 patients. Feature selection is not repeated at each sample size. Intervals use paired patient samples; hospital contrasts are retained in the study source results. Positive capture differences and negative probability-score differences favour organ history.

#### Supplementary Table 11 Simple historical summaries and time in the current state

| **Database** | **Comparison** | **Log loss difference (95% CI)** | **Brier difference (95% CI)** | **Captured candidate / reference** | **Capture difference (95% CI)** |
| --- | --- | --- | --- | --- | --- |
| MIMIC-IV | Add state duration to current information | −0.002003 (−0.002813 to −0.001225) | −0.000219 (−0.000342 to −0.000093) | 369 / 363 | 6.00 (−14.53 to 19.52) |
| MIMIC-IV | Add history summaries to current information | −0.000610 (−0.001242 to 0.000045) | −0.000040 (−0.000154 to 0.000065) | 367 / 363 | 4.00 (−12.53 to 16.00) |
| MIMIC-IV | Add state duration to history summaries | −0.001560 (−0.002227 to −0.000856) | −0.000231 (−0.000349 to −0.000112) | 368 / 367 | 1.00 (−11.53 to 18.52) |
| MIMIC-IV | All history versus summaries and duration | −0.001417 (−0.002339 to −0.000596) | −0.000158 (−0.000312 to −0.000018) | 381 / 368 | 13.00 (−11.00 to 31.00) |
| MIMIC-IV | Add state duration to all history | −0.000040 (−0.000495 to 0.000442) | −0.000081 (−0.000178 to 0.000035) | 385 / 381 | 4.00 (−11.00 to 17.00) |
| eICU | Add state duration to current information | −0.001783 (−0.002460 to −0.001132) | −0.000128 (−0.000258 to −0.000032) | 247 / 219 | 28.00 (4.00 to 50.00) |
| eICU | Add history summaries to current information | −0.001110 (−0.001750 to −0.000474) | −0.000026 (−0.000083 to 0.000033) | 235 / 219 | 16.00 (−5.00 to 40.52) |
| eICU | Add state duration to history summaries | −0.000781 (−0.001241 to −0.000357) | −0.000109 (−0.000193 to −0.000039) | 256 / 235 | 21.00 (−3.00 to 40.52) |
| eICU | All history versus summaries and duration | −0.000172 (−0.000777 to 0.000439) | 0.000137 (0.000031 to 0.000240) | 282 / 256 | 26.00 (3.00 to 49.52) |
| eICU | Add state duration to all history | 0.000201 (−0.000117 to 0.000567) | −0.000018 (−0.000075 to 0.000040) | 262 / 282 | −20.00 (−38.52 to −1.00) |
| SICdb | Add state duration to current information | −0.000552 (−0.003616 to 0.002500) | −0.000217 (−0.000772 to 0.000330) | 35 / 26 | 9.00 (−4.00 to 19.00) |
| SICdb | Add history summaries to current information | 0.002928 (0.000257 to 0.005463) | 0.000457 (0.000031 to 0.000861) | 31 / 26 | 5.00 (−4.00 to 15.00) |
| SICdb | Add state duration to history summaries | −0.002384 (−0.004970 to 0.000291) | −0.000209 (−0.000789 to 0.000318) | 36 / 31 | 5.00 (−9.00 to 16.00) |
| SICdb | All history versus summaries and duration | 0.001335 (−0.000926 to 0.003548) | 0.000828 (0.000198 to 0.001508) | 40 / 36 | 4.00 (−9.53 to 16.00) |
| SICdb | Add state duration to all history | 0.000441 (−0.001085 to 0.001796) | −0.000213 (−0.000701 to 0.000151) | 49 / 40 | 9.00 (0.00 to 19.00) |

Nine scheduled assessments with six-hour patient suppression; six-hour outcome unless otherwise indicated. These are the supporting full-reference comparisons, distinct from the new record-source search. Candidate minus reference differences use matched inputs and review capacity within each database. MIMIC-IV and SICdb use their corresponding matched populations.

#### Supplementary Table 12 Reduction from the mapped candidate features to the full reference

| **Selection step** | **Record groups** | **Inputs** | **MIMIC-IV AUROC** | **MIMIC-IV log loss** | **MIMIC-IV captures** | **eICU AUROC** | **eICU captures** |
| --- | --- | --- | --- | --- | --- | --- | --- |
| 1 | 13 | 552 | 0.9126 | 0.07877 | 387 | 0.8680 | 273 |
| 2 | 13 | 497 | 0.9150 | 0.07813 | 396 | 0.8689 | 280 |
| 3 | 13 | 447 | 0.9134 | 0.07862 | 389 | 0.8703 | 267 |
| 4 | 13 | 402 | 0.9129 | 0.07861 | 394 | 0.8674 | 278 |
| 5 | 13 | 362 | 0.9133 | 0.07854 | 395 | 0.8703 | 278 |
| 6 | 13 | 326 | 0.9134 | 0.07845 | 397 | 0.8678 | 282 |
| 7 | 13 | 294 | 0.9135 | 0.07855 | 387 | 0.8697 | 287 |
| 8 | 13 | 265 | 0.9142 | 0.07842 | 389 | 0.8676 | 267 |
| 9 | 13 | 239 | 0.9142 | 0.07874 | 380 | 0.8699 | 279 |
| 10 | 13 | 215 | 0.9140 | 0.07878 | 368 | 0.8714 | 266 |
| 11 | 13 | 194 | 0.9136 | 0.07877 | 381 | 0.8706 | 269 |
| 12 (selected) | 13 | 175 | 0.9145 | 0.07867 | 381 | 0.8706 | 282 |
| 13 | 13 | 158 | 0.9140 | 0.07883 | 379 | 0.8703 | 280 |
| 14 | 13 | 142 | 0.9120 | 0.07919 | 369 | 0.8677 | 280 |
| 15 | 13 | 128 | 0.9119 | 0.07915 | 373 | 0.8660 | 271 |
| 16 | 13 | 115 | 0.9124 | 0.07913 | 367 | 0.8677 | 266 |
| 17 | 13 | 104 | 0.9115 | 0.07915 | 378 | 0.8646 | 260 |
| 18 | 13 | 94 | 0.9119 | 0.07893 | 383 | 0.8680 | 268 |
| 19 | 13 | 85 | 0.9120 | 0.07928 | 379 | 0.8676 | 273 |
| 20 | 13 | 77 | 0.9107 | 0.07935 | 368 | 0.8682 | 276 |
| 21 | 13 | 69 | 0.9097 | 0.07961 | 366 | 0.8668 | 263 |
| 22 | 13 | 62 | 0.9102 | 0.07967 | 374 | 0.8666 | 269 |
| 23 | 13 | 56 | 0.9095 | 0.07969 | 366 | 0.8647 | 247 |
| 24 | 13 | 51 | 0.9103 | 0.07968 | 369 | 0.8655 | 268 |
| 25 | 13 | 46 | 0.9097 | 0.07970 | 364 | 0.8650 | 265 |
| 26 | 12 | 42 | 0.9112 | 0.07956 | 366 | 0.8606 | 251 |
| 27 | 12 | 38 | 0.9107 | 0.07987 | 361 | 0.8595 | 254 |

Nine scheduled assessments with six-hour patient suppression; six-hour outcome unless otherwise indicated. This scalar reduction follows upstream source selection and cross-database mapping. It is not a comparison with all EHR information. The initial 552 inputs already belong to the 13 record groups retained by group selection. Group counts use the acquisition-group dictionary. Scalar reduction produced 27 evaluated representations through 38 inputs; the selected representation contains 175 inputs. MIMIC-IV evaluation includes 1,960 patients and 980 alerts; eICU includes 4,983 patients and 2,492 alerts. Source selection used 294 MIMIC-IV patients; eICU supplied the multicentre representation comparisons described in the manuscript.

#### Supplementary Table 13 Retention of the full historical gain

| **Database** | **Current inputs** | **Horizon (h)** | **Sampling unit** | **Measure** | **Gain retained (95% CI)** | **No-gain draws** |
| --- | --- | --- | --- | --- | --- | --- |
| MIMIC-IV | 67 | 6 | Patient | Log loss | 0.82845 (0.65416 to 0.97597) | 0 |
| MIMIC-IV | 67 | 6 | Patient | Events captured | 1.06 (CI not estimable) | 85 |
| MIMIC-IV | 91 | 6 | Patient | Log loss | 1.05 (0.92084 to 1.22) | 0 |
| MIMIC-IV | 91 | 6 | Patient | Events captured | 2.69 (CI not estimable) | 183 |
| MIMIC-IV | 67 | 12 | Patient | Log loss | 0.99425 (0.84108 to 1.18) | 0 |
| MIMIC-IV | 67 | 12 | Patient | Events captured | 1.83 (CI not estimable) | 160 |
| MIMIC-IV | 67 | 24 | Patient | Log loss | 1.19 (0.91448 to 1.68) | 0 |
| MIMIC-IV | 67 | 24 | Patient | Events captured | -1.67 (CI not estimable) | 714 |
| eICU | 67 | 6 | Patient | Log loss | 1.15 (0.97899 to 1.50) | 0 |
| eICU | 67 | 6 | Patient | Events captured | 0.84127 (0.53219 to 1.22) | 0 |
| eICU | 67 | 6 | Hospital | Log loss | 1.15 (0.99003 to 1.56) | 1 |
| eICU | 67 | 6 | Hospital | Events captured | 0.84127 (0.51852 to 1.16) | 0 |
| eICU | 91 | 6 | Patient | Log loss | 0.86951 (0.67110 to 1.05) | 0 |
| eICU | 91 | 6 | Patient | Events captured | 0.94203 (0.64286 to 1.19) | 0 |
| eICU | 91 | 6 | Hospital | Log loss | 0.86951 (0.52027 to 1.06) | 0 |
| eICU | 91 | 6 | Hospital | Events captured | 0.94203 (0.59990 to 1.18) | 0 |
| eICU | 67 | 12 | Patient | Log loss | 0.53309 (0.25478 to 0.69146) | 0 |
| eICU | 67 | 12 | Patient | Events captured | 0.68605 (0.43742 to 0.91094) | 0 |
| eICU | 67 | 12 | Hospital | Log loss | 0.53309 (0.20783 to 0.71501) | 0 |
| eICU | 67 | 12 | Hospital | Events captured | 0.68605 (0.46149 to 0.95920) | 0 |
| eICU | 67 | 24 | Patient | Log loss | 0.61716 (-0.36739 to 0.95943) | 4 |
| eICU | 67 | 24 | Patient | Events captured | 0.87368 (0.62500 to 1.10) | 0 |
| eICU | 67 | 24 | Hospital | Log loss | 0.61716 (-0.92854 to 1.07) | 12 |
| eICU | 67 | 24 | Hospital | Events captured | 0.87368 (0.64810 to 1.11) | 0 |

Nine scheduled assessments with six-hour patient suppression; six-hour outcome unless otherwise indicated. Retention is the ratio of improvements over the same current model. Ratios are fractions, not percentages. A confidence interval is not reported when the full-reference denominator is unstable; the number of non-positive denominators among 1,200 draws is shown.

#### Supplementary Table 14 Source-selected candidates evaluated in eICU

| **Candidate ID** | **Historical sources** | **History inputs** | **MIMIC-IV log loss** | **MIMIC-IV captures** | **eICU log loss** | **eICU captures** |
| --- | --- | --- | --- | --- | --- | --- |
| 0 | 0 | 0 | 0.07911 | 1,580 | 0.05406 | 219 |
| 49 | 2 | 19 | 0.07812 | 1,585 | 0.05355 | 250 |
| 256 | 1 | 10 | 0.07896 | 1,603 | 0.05350 | 216 |
| 512 | 1 | 13 | 0.07889 | 1,597 | 0.05350 | 222 |
| 1028 | 2 | 13 | 0.07905 | 1,618 | 0.05407 | 223 |
| 1034 | 3 | 32 | 0.07875 | 1,628 | 0.05336 | 236 |
| 1037 | 4 | 38 | 0.07888 | 1,631 | 0.05339 | 235 |
| 1434 | 6 | 51 | 0.07869 | 1,635 | 0.05389 | 242 |
| 2048 | 1 | 13 | 0.07837 | 1,594 | 0.05447 | 218 |
| 2050 | 2 | 22 | 0.07802 | 1,578 | 0.05359 | 239 |
| 2097 | 3 | 32 | 0.07716 | 1,574 | 0.05304 | 240 |
| 2107 | 5 | 54 | 0.07682 | 1,598 | 0.05238 | 254 |
| 2353 | 4 | 42 | 0.07695 | 1,585 | 0.05315 | 241 |
| 2611 | 5 | 54 | 0.07671 | 1,577 | 0.05242 | 268 |
| 2943 | 9 | 82 | 0.07653 | 1,603 | 0.05311 | 250 |
| 7217 | 4 | 44 | 0.07696 | 1,591 | 0.05278 | 257 |
| 7475 | 6 | 63 | 0.07675 | 1,589 | 0.05233 | 276 |
| 7731 | 6 | 66 | 0.07666 | 1,590 | 0.05229 | 264 |
| 7739 | 7 | 79 | 0.07656 | 1,599 | 0.05238 | 255 |
| 7995 | 8 | 89 | 0.07651 | 1,601 | 0.05222 | 267 |
| 8191 | 11 | 101 | 0.07658 | 1,619 | 0.05224 | 268 |
| 9787 | 7 | 71 | 0.07749 | 1,642 | 0.05239 | 271 |
| 9800 | 5 | 45 | 0.07875 | 1,632 | 0.05329 | 237 |
| 10233 | 9 | 81 | 0.07771 | 1,634 | 0.05319 | 259 |
| 11059 | 7 | 71 | 0.07647 | 1,593 | 0.05232 | 264 |
| 11263 | 11 | 96 | 0.07650 | 1,615 | 0.05261 | 275 |
| 16063 | 10 | 96 | 0.07653 | 1,596 | 0.05173 | 272 |
| 16185 | 8 | 87 | 0.07674 | 1,638 | 0.05244 | 254 |
| 16255 | 11 | 101 | 0.07652 | 1,603 | 0.05244 | 277 |
| 16307 | 9 | 90 | 0.07673 | 1,614 | 0.05230 | 269 |
| 16311 | 10 | 93 | 0.07658 | 1,636 | 0.05251 | 266 |
| 16319 | 11 | 106 | 0.07666 | 1,619 | 0.05245 | 285 |
| 16383 | 12 | 108 | 0.07662 | 1,592 | 0.05203 | 282 |

The input signature uniquely indexes each deduplicated feature matrix. MIMIC-IV values are development out-of-fold results; eICU contributed to selection. Source and eICU alert budgets are 4,495 and 2,492. The complete source-combination distribution is summarised in Figure 2.

#### Supplementary Table 15 Six-hour performance by organ

| **Database** | **Information** | **Organ** | **Patients** | **Positive windows** | **AUROC** | **Log loss** | **Brier score** | **Events captured** |
| --- | --- | --- | --- | --- | --- | --- | --- | --- |
| MIMIC-IV | Current | Respiratory | 391 | 165 | 0.6741 | 0.47359 | 0.14938 | 5 |
| MIMIC-IV | Current | Coagulation | 1,614 | 202 | 0.9148 | 0.12657 | 0.03344 | 31 |
| MIMIC-IV | Current | Liver | 859 | 41 | 0.9076 | 0.05504 | 0.01199 | 1 |
| MIMIC-IV | Current | Circulatory | 1,935 | 702 | 0.9048 | 0.21577 | 0.06571 | 298 |
| MIMIC-IV | Current | Neurological | 1,857 | 257 | 0.7718 | 0.13028 | 0.02984 | 0 |
| MIMIC-IV | Current | Renal | 1,620 | 188 | 0.9179 | 0.12009 | 0.03098 | 28 |
| MIMIC-IV | Full | Respiratory | 391 | 165 | 0.7642 | 0.43222 | 0.13590 | 30 |
| MIMIC-IV | Full | Coagulation | 1,614 | 202 | 0.9234 | 0.12073 | 0.03262 | 28 |
| MIMIC-IV | Full | Liver | 859 | 41 | 0.9167 | 0.05715 | 0.01222 | 0 |
| MIMIC-IV | Full | Circulatory | 1,935 | 702 | 0.9179 | 0.20422 | 0.06362 | 291 |
| MIMIC-IV | Full | Neurological | 1,857 | 257 | 0.7902 | 0.12584 | 0.02943 | 1 |
| MIMIC-IV | Full | Renal | 1,620 | 188 | 0.9502 | 0.10705 | 0.02921 | 31 |
| MIMIC-IV | Compact | Respiratory | 391 | 165 | 0.7724 | 0.42258 | 0.13303 | 31 |
| MIMIC-IV | Compact | Coagulation | 1,614 | 202 | 0.9203 | 0.12533 | 0.03282 | 27 |
| MIMIC-IV | Compact | Liver | 859 | 41 | 0.9076 | 0.05930 | 0.01226 | 0 |
| MIMIC-IV | Compact | Circulatory | 1,935 | 702 | 0.9153 | 0.20600 | 0.06405 | 291 |
| MIMIC-IV | Compact | Neurological | 1,857 | 257 | 0.7854 | 0.12670 | 0.02950 | 1 |
| MIMIC-IV | Compact | Renal | 1,620 | 188 | 0.9469 | 0.10427 | 0.02848 | 32 |
| eICU | Current | Respiratory | 660 | 123 | 0.6371 | 0.31389 | 0.08704 | 42 |
| eICU | Current | Coagulation | 4,062 | 287 | 0.9014 | 0.07570 | 0.01783 | 72 |
| eICU | Current | Liver | 3,179 | 65 | 0.9299 | 0.02857 | 0.00569 | 1 |
| eICU | Current | Circulatory | 3,715 | 218 | 0.7717 | 0.11385 | 0.02337 | 14 |
| eICU | Current | Neurological | 3,064 | 528 | 0.6481 | 0.23972 | 0.05553 | 37 |
| eICU | Current | Renal | 3,776 | 234 | 0.8762 | 0.07879 | 0.01825 | 53 |
| eICU | Full | Respiratory | 660 | 123 | 0.7052 | 0.29890 | 0.08316 | 30 |
| eICU | Full | Coagulation | 4,062 | 287 | 0.9079 | 0.07391 | 0.01758 | 69 |
| eICU | Full | Liver | 3,179 | 65 | 0.9317 | 0.03076 | 0.00575 | 1 |
| eICU | Full | Circulatory | 3,715 | 218 | 0.8749 | 0.09126 | 0.02158 | 56 |
| eICU | Full | Neurological | 3,064 | 528 | 0.7043 | 0.22689 | 0.05444 | 55 |
| eICU | Full | Renal | 3,776 | 234 | 0.9042 | 0.07521 | 0.01861 | 71 |
| eICU | Compact | Respiratory | 660 | 123 | 0.7094 | 0.29604 | 0.08264 | 32 |
| eICU | Compact | Coagulation | 4,062 | 287 | 0.9192 | 0.07339 | 0.01749 | 68 |
| eICU | Compact | Liver | 3,179 | 65 | 0.9308 | 0.03046 | 0.00572 | 2 |
| eICU | Compact | Circulatory | 3,715 | 218 | 0.8755 | 0.09139 | 0.02173 | 59 |
| eICU | Compact | Neurological | 3,064 | 528 | 0.7043 | 0.22689 | 0.05457 | 49 |
| eICU | Compact | Renal | 3,776 | 234 | 0.8999 | 0.07403 | 0.01782 | 62 |

Nine scheduled assessments with six-hour patient suppression; six-hour outcome unless otherwise indicated. Signatures 0, 16063 and 16383 denote current, compact and full history. Event windows count repeated assessments, while captures count distinct episodes.

#### Supplementary Table 16 Paired event capture under hourly reassessment

| **Database** | **Information** | **Comparator** | **Sampling unit** | **Difference (95% CI)** |
| --- | --- | --- | --- | --- |
| MIMIC-IV | Compact history | Current information | Patient | 15 (-4 to 38) |
| MIMIC-IV | Compact history | Full history | Patient | 14 (-4 to 33) |
| MIMIC-IV | Full history | Current information | Patient | 1 (-21 to 24.52) |
| MIMIC-IV | Brief history | Current information | Patient | 0 (-16 to 23) |
| MIMIC-IV | Compact history | Brief history | Patient | 15 (-6 to 31.52) |
| eICU | Compact history | Current information | Patient | 74 (49 to 106.52) |
| eICU | Compact history | Full history | Patient | 5 (-14 to 28.52) |
| eICU | Full history | Current information | Patient | 69 (41 to 101.52) |
| eICU | Brief history | Current information | Patient | 67 (34.48 to 96) |
| eICU | Compact history | Brief history | Patient | 7 (-10.53 to 30) |
| eICU | Compact history | Current information | Hospital | 74 (43.28 to 106.86) |
| eICU | Compact history | Full history | Hospital | 5 (-11.75 to 24.47) |
| eICU | Full history | Current information | Hospital | 69 (33.19 to 102.24) |
| eICU | Brief history | Current information | Hospital | 67 (32.74 to 93.57) |
| eICU | Compact history | Brief history | Hospital | 7 (-11.86 to 35.82) |
| CareVue | Compact history | Current information | Patient | 37 (14 to 54) |
| CareVue | Compact history | Full history | Patient | -1 (-14 to 10) |
| CareVue | Full history | Current information | Patient | 38 (15 to 57.52) |
| CareVue | Brief history | Current information | Patient | 27 (3 to 45) |
| CareVue | Compact history | Brief history | Patient | 10 (-5 to 23) |
| SICdb | Compact history | Current information | Patient | 22 (8 to 35) |
| SICdb | Compact history | Full history | Patient | 10 (-1 to 24) |
| SICdb | Full history | Current information | Patient | 12 (-1 to 20.52) |
| SICdb | Brief history | Current information | Patient | 4 (-5 to 11) |
| SICdb | Compact history | Brief history | Patient | 18 (6 to 29.52) |

Hourly reassessment with four-hour patient suppression and a six-hour outcome. All methods receive the same alert budget. Intervals use 500 paired draws with allocation reconstructed in each draw.

#### Supplementary Table 17 Shared, lost and added events after compression

| **Database** | **Information** | **Organ** | **Shared captures** | **Full only** | **Added captures** |
| --- | --- | --- | --- | --- | --- |
| MIMIC-IV | Brief history | All | 374 | 60 | 59 |
| MIMIC-IV | Brief history | Respiratory | 1 | 15 | 0 |
| MIMIC-IV | Brief history | Coagulation | 17 | 6 | 11 |
| MIMIC-IV | Brief history | Liver | 0 | 0 | 0 |
| MIMIC-IV | Brief history | Circulatory | 324 | 29 | 44 |
| MIMIC-IV | Brief history | Neurological | 0 | 1 | 0 |
| MIMIC-IV | Brief history | Renal | 32 | 9 | 4 |
| MIMIC-IV | Compact history | All | 378 | 56 | 70 |
| MIMIC-IV | Compact history | Respiratory | 15 | 1 | 10 |
| MIMIC-IV | Compact history | Coagulation | 16 | 7 | 9 |
| MIMIC-IV | Compact history | Liver | 0 | 0 | 0 |
| MIMIC-IV | Compact history | Circulatory | 317 | 36 | 47 |
| MIMIC-IV | Compact history | Neurological | 0 | 1 | 0 |
| MIMIC-IV | Compact history | Renal | 30 | 11 | 4 |
| eICU | Brief history | All | 256 | 58 | 56 |
| eICU | Brief history | Respiratory | 17 | 11 | 9 |
| eICU | Brief history | Coagulation | 58 | 14 | 20 |
| eICU | Brief history | Liver | 0 | 1 | 1 |
| eICU | Brief history | Circulatory | 54 | 8 | 7 |
| eICU | Brief history | Neurological | 37 | 11 | 11 |
| eICU | Brief history | Renal | 90 | 13 | 8 |
| eICU | Compact history | All | 260 | 54 | 59 |
| eICU | Compact history | Respiratory | 25 | 3 | 9 |
| eICU | Compact history | Coagulation | 62 | 10 | 18 |
| eICU | Compact history | Liver | 1 | 0 | 3 |
| eICU | Compact history | Circulatory | 57 | 5 | 9 |
| eICU | Compact history | Neurological | 37 | 11 | 11 |
| eICU | Compact history | Renal | 78 | 25 | 9 |
| CareVue | Brief history | All | 101 | 44 | 33 |
| CareVue | Brief history | Respiratory | 10 | 30 | 8 |
| CareVue | Brief history | Coagulation | 11 | 0 | 7 |
| CareVue | Brief history | Liver | 0 | 1 | 0 |
| CareVue | Brief history | Circulatory | 31 | 2 | 9 |
| CareVue | Brief history | Neurological | 5 | 6 | 1 |
| CareVue | Brief history | Renal | 44 | 5 | 8 |
| CareVue | Compact history | All | 119 | 26 | 25 |
| CareVue | Compact history | Respiratory | 32 | 8 | 6 |
| CareVue | Compact history | Coagulation | 10 | 1 | 7 |
| CareVue | Compact history | Liver | 0 | 1 | 0 |
| CareVue | Compact history | Circulatory | 32 | 1 | 7 |
| CareVue | Compact history | Neurological | 5 | 6 | 0 |
| CareVue | Compact history | Renal | 40 | 9 | 5 |
| SICdb | Brief history | All | 26 | 13 | 5 |
| SICdb | Brief history | Respiratory | 0 | 5 | 0 |
| SICdb | Brief history | Coagulation | 6 | 0 | 2 |
| SICdb | Brief history | Liver | 0 | 0 | 0 |
| SICdb | Brief history | Circulatory | 7 | 7 | 2 |
| SICdb | Brief history | Neurological | 0 | 0 | 0 |
| SICdb | Brief history | Renal | 13 | 1 | 1 |
| SICdb | Compact history | All | 27 | 12 | 22 |
| SICdb | Compact history | Respiratory | 5 | 0 | 11 |
| SICdb | Compact history | Coagulation | 6 | 0 | 3 |
| SICdb | Compact history | Liver | 0 | 0 | 0 |
| SICdb | Compact history | Circulatory | 8 | 6 | 8 |
| SICdb | Compact history | Neurological | 0 | 0 | 0 |
| SICdb | Compact history | Renal | 8 | 6 | 0 |

Hourly reassessment with four-hour patient suppression and a six-hour outcome. All methods receive the same alert budget. Shared events are captured by both compared models; full-only and compact-only events are mutually exclusive.

#### Supplementary Table 18 Matched opportunities for local probability calibration

| **Database** | **Information** | **AUROC** | **Log loss** | **Brier score** | **Mean risk** | **Observed risk** | **Events captured** |
| --- | --- | --- | --- | --- | --- | --- | --- |
| MIMIC-IV | Current information | 0.8979 | 0.08219 | 0.02056 | 0.02209 | 0.02445 | 363 |
| MIMIC-IV | Compact history | 0.9114 | 0.07927 | 0.02020 | 0.02185 | 0.02445 | 382 |
| MIMIC-IV | Full history | 0.9145 | 0.07867 | 0.02014 | 0.02155 | 0.02445 | 381 |
| MIMIC-IV | Current SOFA total | 0.6506 | 0.11011 | 0.02353 | 0.02255 | 0.02445 | 109 |
| MIMIC-IV | Current SOFA components | 0.7771 | 0.10245 | 0.02317 | 0.02268 | 0.02445 | 130 |
| MIMIC-IV | Current organ states | 0.7079 | 0.10726 | 0.02334 | 0.02258 | 0.02445 | 126 |
| MIMIC-IV | Own-organ history | 0.9130 | 0.07916 | 0.02022 | 0.02202 | 0.02445 | 386 |
| MIMIC-IV | Current information, global calibration | 0.8976 | 0.08201 | 0.02056 | 0.02446 | 0.02445 | 367 |
| MIMIC-IV | Current information, organ calibration | 0.8975 | 0.08200 | 0.02054 | 0.02448 | 0.02445 | 369 |
| MIMIC-IV | Local organ prior | 0.6334 | 0.11287 | 0.02375 | 0.02447 | 0.02445 | 70 |
| MIMIC-IV | Compact history, global calibration | 0.9113 | 0.07908 | 0.02019 | 0.02445 | 0.02445 | 385 |
| MIMIC-IV | Compact history, organ calibration | 0.9115 | 0.07907 | 0.02018 | 0.02447 | 0.02445 | 389 |
| MIMIC-IV | Full history, global calibration | 0.9144 | 0.07844 | 0.02012 | 0.02445 | 0.02445 | 379 |
| MIMIC-IV | Full history, organ calibration | 0.9143 | 0.07845 | 0.02012 | 0.02447 | 0.02445 | 384 |
| MIMIC-IV | Own-organ history, global calibration | 0.9129 | 0.07898 | 0.02021 | 0.02445 | 0.02445 | 386 |
| MIMIC-IV | Own-organ history, organ calibration | 0.9130 | 0.07897 | 0.02021 | 0.02447 | 0.02445 | 384 |
| eICU | Current information | 0.8418 | 0.05406 | 0.01128 | 0.01232 | 0.01177 | 219 |
| eICU | Compact history | 0.8695 | 0.05173 | 0.01114 | 0.01338 | 0.01177 | 272 |
| eICU | Full history | 0.8706 | 0.05203 | 0.01130 | 0.01363 | 0.01177 | 282 |
| eICU | Current SOFA total | 0.4747 | 0.06830 | 0.01190 | 0.02036 | 0.01177 | 84 |
| eICU | Current SOFA components | 0.7476 | 0.06224 | 0.01189 | 0.02086 | 0.01177 | 94 |
| eICU | Current organ states | 0.5657 | 0.06769 | 0.01197 | 0.01992 | 0.01177 | 85 |
| eICU | Own-organ history | 0.8707 | 0.05182 | 0.01118 | 0.01319 | 0.01177 | 284 |
| eICU | Current information, global calibration | 0.8414 | 0.05376 | 0.01119 | 0.01176 | 0.01177 | 220 |
| eICU | Current information, organ calibration | 0.8447 | 0.05334 | 0.01113 | 0.01176 | 0.01177 | 223 |
| eICU | Local organ prior | 0.6542 | 0.06174 | 0.01156 | 0.01177 | 0.01177 | 101 |
| eICU | Compact history, global calibration | 0.8692 | 0.05137 | 0.01104 | 0.01176 | 0.01177 | 271 |
| eICU | Compact history, organ calibration | 0.8760 | 0.05050 | 0.01094 | 0.01175 | 0.01177 | 269 |
| eICU | Full history, global calibration | 0.8704 | 0.05146 | 0.01109 | 0.01176 | 0.01177 | 281 |
| eICU | Full history, organ calibration | 0.8748 | 0.05072 | 0.01098 | 0.01175 | 0.01177 | 282 |
| eICU | Own-organ history, global calibration | 0.8704 | 0.05150 | 0.01106 | 0.01176 | 0.01177 | 284 |
| eICU | Own-organ history, organ calibration | 0.8748 | 0.05079 | 0.01099 | 0.01175 | 0.01177 | 286 |
| CareVue | Current information | 0.8380 | 0.10094 | 0.02298 | 0.01362 | 0.02435 | 108 |
| CareVue | Compact history | 0.8487 | 0.09904 | 0.02269 | 0.01568 | 0.02435 | 126 |
| CareVue | Full history | 0.8514 | 0.09864 | 0.02265 | 0.01490 | 0.02435 | 117 |
| CareVue | Current SOFA total | 0.5314 | 0.11433 | 0.02359 | 0.02342 | 0.02435 | 100 |
| CareVue | Current SOFA components | 0.7490 | 0.10522 | 0.02321 | 0.01898 | 0.02435 | 86 |
| CareVue | Current organ states | 0.6387 | 0.11216 | 0.02346 | 0.01934 | 0.02435 | 85 |
| CareVue | Own-organ history | 0.8536 | 0.09734 | 0.02259 | 0.01549 | 0.02435 | 117 |
| CareVue | Current information, global calibration | 0.8373 | 0.09584 | 0.02270 | 0.02434 | 0.02435 | 109 |
| CareVue | Current information, organ calibration | 0.8500 | 0.09422 | 0.02258 | 0.02441 | 0.02435 | 105 |
| CareVue | Compact history, global calibration | 0.8479 | 0.09468 | 0.02252 | 0.02439 | 0.02435 | 129 |
| CareVue | Compact history, organ calibration | 0.8553 | 0.09284 | 0.02235 | 0.02445 | 0.02435 | 129 |
| CareVue | Full history, global calibration | 0.8507 | 0.09412 | 0.02245 | 0.02439 | 0.02435 | 117 |
| CareVue | Full history, organ calibration | 0.8580 | 0.09260 | 0.02233 | 0.02444 | 0.02435 | 130 |
| CareVue | Own-organ history, global calibration | 0.8530 | 0.09364 | 0.02244 | 0.02438 | 0.02435 | 121 |
| CareVue | Own-organ history, organ calibration | 0.8609 | 0.09196 | 0.02230 | 0.02446 | 0.02435 | 131 |
| CareVue | Local organ prior | 0.6758 | 0.10747 | 0.02323 | 0.02436 | 0.02435 | 104 |
| SICdb | Current information | 0.8591 | 0.09360 | 0.02317 | 0.02815 | 0.02389 | 26 |
| SICdb | Compact history | 0.8742 | 0.09507 | 0.02397 | 0.04184 | 0.02389 | 46 |
| SICdb | Full history | 0.8698 | 0.09548 | 0.02437 | 0.03821 | 0.02389 | 40 |
| SICdb | Current SOFA total | 0.7923 | 0.09850 | 0.02234 | 0.02247 | 0.02389 | 61 |
| SICdb | Current SOFA components | 0.8260 | 0.09558 | 0.02257 | 0.02441 | 0.02389 | 33 |
| SICdb | Current organ states | 0.7886 | 0.09990 | 0.02246 | 0.02844 | 0.02389 | 49 |
| SICdb | Own-organ history | 0.8679 | 0.09395 | 0.02334 | 0.03614 | 0.02389 | 30 |
| SICdb | Current information, global calibration | 0.8579 | 0.09292 | 0.02258 | 0.02394 | 0.02389 | 25 |
| SICdb | Current information, organ calibration | 0.8788 | 0.08709 | 0.02174 | 0.02389 | 0.02389 | 51 |
| SICdb | Compact history, global calibration | 0.8728 | 0.08865 | 0.02184 | 0.02394 | 0.02389 | 47 |
| SICdb | Compact history, organ calibration | 0.8854 | 0.08556 | 0.02126 | 0.02387 | 0.02389 | 63 |
| SICdb | Full history, global calibration | 0.8681 | 0.09011 | 0.02212 | 0.02394 | 0.02389 | 38 |
| SICdb | Full history, organ calibration | 0.8854 | 0.08555 | 0.02127 | 0.02386 | 0.02389 | 61 |
| SICdb | Own-organ history, global calibration | 0.8662 | 0.09016 | 0.02197 | 0.02393 | 0.02389 | 33 |
| SICdb | Own-organ history, organ calibration | 0.8812 | 0.08653 | 0.02138 | 0.02386 | 0.02389 | 56 |
| SICdb | Local organ prior | 0.7672 | 0.09890 | 0.02228 | 0.02391 | 0.02389 | 61 |

Nine scheduled assessments with six-hour patient suppression; six-hour outcome unless otherwise indicated.

#### Supplementary Table 19 Selected probability maps at six, twelve and twenty-four hours

**MIMIC-IV · Source calibration**

| **Horizon (h)** | **Information** | **Method** | **Patients** | **AUROC** | **Log loss** | **Brier score** | **Events captured** |
| --- | --- | --- | --- | --- | --- | --- | --- |
| 6 | Current information | Unchanged | 1,960 | 0.8979 | 0.08219 | 0.02056 | 363 |
| 6 | Compact history | Unchanged | 1,960 | 0.9114 | 0.07927 | 0.02020 | 382 |
| 6 | Full history | Unchanged | 1,960 | 0.9145 | 0.07867 | 0.02014 | 381 |
| 6 | Own-organ history | Unchanged | 1,960 | 0.9130 | 0.07916 | 0.02022 | 386 |
| 12 | Current information | Unchanged | 1,960 | 0.8947 | 0.13372 | 0.03644 | 504 |
| 12 | Compact history | Unchanged | 1,960 | 0.9064 | 0.12925 | 0.03567 | 526 |
| 12 | Full history | Unchanged | 1,960 | 0.9070 | 0.12923 | 0.03570 | 516 |
| 24 | Current information | Unchanged | 1,960 | 0.8864 | 0.18831 | 0.05444 | 571 |
| 24 | Compact history | Unchanged | 1,960 | 0.8984 | 0.18274 | 0.05357 | 576 |
| 24 | Full history | Unchanged | 1,960 | 0.8969 | 0.18361 | 0.05361 | 568 |

**eICU · Source calibration**

| **Horizon (h)** | **Information** | **Method** | **Patients** | **AUROC** | **Log loss** | **Brier score** | **Events captured** |
| --- | --- | --- | --- | --- | --- | --- | --- |
| 6 | Current information | Unchanged | 4,983 | 0.8418 | 0.05406 | 0.01128 | 219 |
| 6 | Compact history | Unchanged | 4,983 | 0.8695 | 0.05173 | 0.01114 | 272 |
| 6 | Full history | Unchanged | 4,983 | 0.8706 | 0.05203 | 0.01130 | 282 |
| 6 | Own-organ history | Unchanged | 4,983 | 0.8707 | 0.05182 | 0.01118 | 284 |
| 12 | Current information | Unchanged | 4,983 | 0.8288 | 0.10164 | 0.02418 | 332 |
| 12 | Compact history | Unchanged | 4,983 | 0.8478 | 0.09965 | 0.02429 | 391 |
| 12 | Full history | Unchanged | 4,983 | 0.8538 | 0.09790 | 0.02389 | 418 |
| 24 | Current information | Unchanged | 4,983 | 0.8249 | 0.15603 | 0.04084 | 400 |
| 24 | Compact history | Unchanged | 4,983 | 0.8374 | 0.15432 | 0.04131 | 483 |
| 24 | Full history | Unchanged | 4,983 | 0.8369 | 0.15326 | 0.04067 | 495 |

**CareVue · Source calibration**

| **Horizon (h)** | **Information** | **Method** | **Patients** | **AUROC** | **Log loss** | **Brier score** | **Events captured** |
| --- | --- | --- | --- | --- | --- | --- | --- |
| 6 | Current information | Unchanged | 1,288 | 0.8380 | 0.10094 | 0.02298 | 108 |
| 6 | Compact history | Unchanged | 1,288 | 0.8487 | 0.09904 | 0.02269 | 126 |
| 6 | Full history | Unchanged | 1,288 | 0.8514 | 0.09864 | 0.02265 | 117 |
| 6 | Own-organ history | Unchanged | 1,288 | 0.8536 | 0.09734 | 0.02259 | 117 |
| 12 | Current information | Unchanged | 1,288 | 0.8243 | 0.17368 | 0.04414 | 170 |
| 12 | Compact history | Unchanged | 1,288 | 0.8335 | 0.17063 | 0.04429 | 193 |
| 12 | Full history | Unchanged | 1,288 | 0.8308 | 0.17180 | 0.04431 | 197 |
| 24 | Current information | Unchanged | 1,288 | 0.8152 | 0.25344 | 0.07028 | 226 |
| 24 | Compact history | Unchanged | 1,288 | 0.8242 | 0.25008 | 0.07048 | 243 |
| 24 | Full history | Unchanged | 1,288 | 0.8219 | 0.24986 | 0.07023 | 238 |

**SICdb · Source calibration**

| **Horizon (h)** | **Information** | **Method** | **Patients** | **AUROC** | **Log loss** | **Brier score** | **Events captured** |
| --- | --- | --- | --- | --- | --- | --- | --- |
| 6 | Current information | Unchanged | 514 | 0.8591 | 0.09360 | 0.02317 | 26 |
| 6 | Compact history | Unchanged | 514 | 0.8742 | 0.09507 | 0.02397 | 46 |
| 6 | Full history | Unchanged | 514 | 0.8698 | 0.09548 | 0.02437 | 40 |
| 6 | Own-organ history | Unchanged | 514 | 0.8679 | 0.09395 | 0.02334 | 30 |
| 12 | Current information | Unchanged | 514 | 0.8376 | 0.15863 | 0.04315 | 57 |
| 12 | Compact history | Unchanged | 514 | 0.8616 | 0.16259 | 0.04718 | 68 |
| 12 | Full history | Unchanged | 514 | 0.8630 | 0.15777 | 0.04467 | 71 |
| 24 | Current information | Unchanged | 514 | 0.8332 | 0.22192 | 0.06610 | 48 |
| 24 | Compact history | Unchanged | 514 | 0.8539 | 0.22585 | 0.06958 | 88 |
| 24 | Full history | Unchanged | 514 | 0.8545 | 0.21905 | 0.06650 | 93 |

**eICU: 91 evaluation hospitals · 23-hospital adaptation**

| **Horizon (h)** | **Information** | **Method** | **Patients** | **AUROC** | **Log loss** | **Brier score** | **Events captured** |
| --- | --- | --- | --- | --- | --- | --- | --- |
| 6 | Current information | Organ logistic (λ = 1) | 4,196 | 0.8429 | 0.05350 | 0.01115 | 198 |
| 6 | Compact history | Organ logistic (λ = 1) | 4,196 | 0.8740 | 0.05079 | 0.01098 | 233 |
| 6 | Full history | Organ logistic (λ = 1) | 4,196 | 0.8749 | 0.05088 | 0.01101 | 236 |
| 6 | Own-organ history | Organ logistic (λ = 1) | 4,196 | 0.8755 | 0.05100 | 0.01103 | 236 |
| 12 | Current information | Organ beta (λ = 1) | 4,196 | 0.8343 | 0.09973 | 0.02377 | 305 |
| 12 | Compact history | Organ beta (λ = 1) | 4,196 | 0.8572 | 0.09651 | 0.02349 | 365 |
| 12 | Full history | Organ beta (λ = 1) | 4,196 | 0.8597 | 0.09600 | 0.02341 | 363 |
| 24 | Current information | Organ beta (λ = 5) | 4,196 | 0.8312 | 0.15212 | 0.03983 | 363 |
| 24 | Compact history | Organ beta (λ = 5) | 4,196 | 0.8435 | 0.14950 | 0.03965 | 428 |
| 24 | Full history | Organ beta (λ = 5) | 4,196 | 0.8416 | 0.14994 | 0.03958 | 408 |

Nine scheduled assessments with six-hour patient suppression; six-hour outcome unless otherwise indicated. Source selection and the eICU 23-hospital adaptation / 91-hospital evaluation are distinct. Only selected maps and matched current comparators are shown; all candidates are retained in the source results. Organ logistic calibration shares a slope and shrinks organ intercepts. Organ beta calibration shares the coefficients of log(p) and −log(1−p) and shrinks organ intercepts; λ is the intercept penalty.

#### Supplementary Table 20 Action-matched net benefit at example risk thresholds

| **Database** | **Information** | **Risk threshold** | **Net benefit (95% CI)** | **Actions per 100 patients** |
| --- | --- | --- | --- | --- |
| MIMIC-IV | Current information | 0.05 | 0.06678 (0.05454 to 0.07997) | 39.44 |
| MIMIC-IV | Current information | 0.10 | 0.05096 (0.03917 to 0.06361) | 28.62 |
| MIMIC-IV | Compact history | 0.05 | 0.07197 (0.05913 to 0.08545) | 41.84 |
| MIMIC-IV | Compact history | 0.10 | 0.05550 (0.04308 to 0.06854) | 30.66 |
| MIMIC-IV | Full history | 0.05 | 0.06917 (0.05703 to 0.08308) | 42.04 |
| MIMIC-IV | Full history | 0.10 | 0.05397 (0.04172 to 0.06622) | 30 |
| MIMIC-IV | Compact history, adapted to missing records | 0.05 | 0.06896 (0.05665 to 0.08282) | 42.45 |
| MIMIC-IV | Compact history, adapted to missing records | 0.10 | 0.05368 (0.04133 to 0.06723) | 31.28 |
| eICU | Current information | 0.05 | 0.00913 (0.00559 to 0.01308) | 21.59 |
| eICU | Current information | 0.10 | 0.00085 (-0.00152 to 0.00346) | 7.26 |
| eICU | Compact history | 0.05 | 0.01224 (0.00819 to 0.01670) | 26.91 |
| eICU | Compact history | 0.10 | 0.00227 (-0.00125 to 0.00629) | 16.01 |
| eICU | Full history | 0.05 | 0.01270 (0.00856 to 0.01697) | 26.45 |
| eICU | Full history | 0.10 | 0.00248 (-0.00134 to 0.00642) | 15.83 |
| eICU | Compact history, adapted to missing records | 0.05 | 0.01349 (0.00921 to 0.01809) | 26.55 |
| eICU | Compact history, adapted to missing records | 0.10 | 0.00381 (0.00007 to 0.00774) | 15.83 |

At index, each action nominates at most one eligible organ. Net benefit credits an event only in that organ; the denominator is all eligible patients. Pointwise patient intervals use 1,200 draws. The example thresholds are selected from the full action-matched curves.

#### Supplementary Table 21 Adaptation to persistent missing records

| **Database** | **Condition** | **Method** | **Values** | **Total inputs** | **Log loss** | **Brier score** | **Events captured** |
| --- | --- | --- | --- | --- | --- | --- | --- |
| eICU | Available records | Dm | 163 | 326 | 0.05262 | 0.01139 | 269 |
| eICU | Available records | D0 | 163 | 163 | 0.05146 | 0.01107 | 272 |
| eICU | 50% batch withholding, assignment 1 | Dm | 163 | 326 | 0.05807 | 0.01188 | 206 |
| eICU | 50% batch withholding, assignment 1 | D0 | 163 | 163 | 0.06349 | 0.01259 | 175 |
| eICU | 50% batch withholding, assignment 2 | Dm | 163 | 326 | 0.05841 | 0.01204 | 215 |
| eICU | 50% batch withholding, assignment 2 | D0 | 163 | 163 | 0.06280 | 0.01252 | 183 |
| eICU | 50% batch withholding, assignment 3 | Dm | 163 | 326 | 0.05828 | 0.01202 | 212 |
| eICU | 50% batch withholding, assignment 3 | D0 | 163 | 163 | 0.06264 | 0.01251 | 182 |
| eICU | 25% batch withholding | Dm | 163 | 326 | 0.07183 | 0.01628 | 155 |
| eICU | 25% batch withholding | D0 | 163 | 163 | 0.07130 | 0.01618 | 150 |
| eICU | 75% batch withholding | Dm | 163 | 326 | 0.07618 | 0.01628 | 131 |
| eICU | 75% batch withholding | D0 | 163 | 163 | 0.08070 | 0.01679 | 118 |
| eICU | 1-hour record window | Dm | 163 | 326 | 0.08222 | 0.01649 | 93 |
| eICU | 1-hour record window | D0 | 163 | 163 | 0.09104 | 0.01665 | 52 |
| eICU | 3-hour record window | Dm | 163 | 326 | 0.08005 | 0.01627 | 99 |
| eICU | 3-hour record window | D0 | 163 | 163 | 0.08733 | 0.01655 | 63 |
| eICU | 6-hour record window | Dm | 163 | 326 | 0.07786 | 0.01636 | 126 |
| eICU | 6-hour record window | D0 | 163 | 163 | 0.08055 | 0.01624 | 108 |
| eICU | 12-hour record window | Dm | 163 | 326 | 0.07522 | 0.01642 | 139 |
| eICU | 12-hour record window | D0 | 163 | 163 | 0.07624 | 0.01598 | 141 |
| eICU | 24-hour record window | Dm | 163 | 326 | 0.07092 | 0.01619 | 168 |
| eICU | 24-hour record window | D0 | 163 | 163 | 0.06851 | 0.01560 | 163 |
| eICU | Three-hour initial history | Dm | 163 | 326 | 0.05808 | 0.01155 | 188 |
| eICU | Three-hour initial history | D0 | 163 | 163 | 0.06449 | 0.01161 | 152 |
| MIMIC-IV | Available records | Dm | 163 | 326 | 0.07950 | 0.02020 | 377 |
| MIMIC-IV | Available records | D0 | 163 | 163 | 0.07953 | 0.02027 | 382 |
| MIMIC-IV | 50% batch withholding, assignment 1 | Dm | 163 | 326 | 0.08522 | 0.02103 | 347 |
| MIMIC-IV | 50% batch withholding, assignment 1 | D0 | 163 | 163 | 0.09351 | 0.02237 | 275 |
| CareVue | Naturally missing records | Dm | 163 | 326 | 0.09737 | 0.02268 | 123 |
| CareVue | Naturally missing records | D0 | 163 | 163 | 0.09829 | 0.02265 | 126 |
| SICdb | Naturally missing records | Dm | 163 | 326 | 0.09654 | 0.02490 | 45 |
| SICdb | Naturally missing records | D0 | 163 | 163 | 0.09227 | 0.02296 | 46 |

Six-hour outcomes are used. Complete records, three 50% withholding assignments and the three-hour initial-history condition are evaluated across nine scheduled assessments with six-hour patient suppression; 25% and 75% withholding and rolling-window restrictions use index assessments only. Initial history keeps records from three hours before index onward, allowing later records to accumulate. D0 is the original predictor; Dm is trained with missing-record views and includes availability indicators. Mean capture across the three 50% assignments is the main contrast.

#### Supplementary Table 22 New information and adaptation for the same future outcome

| **Database** | **Condition** | **Information** | **Patients** | **Log loss** | **Brier score** | **Events captured** |
| --- | --- | --- | --- | --- | --- | --- |
| MIMIC-IV | Available records | Original model, clock update only | 1,932 | 0.07808 | 0.01989 | 288 |
| MIMIC-IV | Available records | Original model, new contents | 1,932 | 0.07478 | 0.01925 | 324 |
| MIMIC-IV | Available records | Adapted model, clock update only | 1,932 | 0.07743 | 0.01969 | 295 |
| MIMIC-IV | Available records | Adapted model, new contents | 1,932 | 0.07444 | 0.01907 | 315 |
| eICU | Available records | Original model, clock update only | 4,832 | 0.05035 | 0.01039 | 182 |
| eICU | Available records | Original model, new contents | 4,832 | 0.04922 | 0.01029 | 203 |
| eICU | Available records | Adapted model, clock update only | 4,832 | 0.05088 | 0.01056 | 184 |
| eICU | Available records | Adapted model, new contents | 4,832 | 0.04975 | 0.01043 | 201 |
| eICU | 50% batch withholding | Original model, clock update only | 4,835 | 0.06381 | 0.01229 | 128 |
| eICU | 50% batch withholding | Original model, new contents | 4,835 | 0.06196 | 0.01199 | 143 |
| eICU | 50% batch withholding | Adapted model, clock update only | 4,835 | 0.05717 | 0.01136 | 160 |
| eICU | 50% batch withholding | Adapted model, new contents | 4,835 | 0.05585 | 0.01121 | 170 |

Nine scheduled assessments with six-hour patient suppression; six-hour outcome unless otherwise indicated. Old and new predictions share the future target. D0 is the original predictor, Dm the adapted predictor; full and batch50 denote complete and persistently withheld records.

#### Supplementary Table 23 Timely organ selection after retrieval or recorded arrival

| **Database** | **Policy** | **Requests** | **Timely correct choices** | **Corrected** | **Worsened** | **Changed choices** |
| --- | --- | --- | --- | --- | --- | --- |
| eICU | Immediate assessment | 2,492 | 145 | 0 | 0 | 0 |
| eICU | Retrieve one existing fact | 2,492 | 159 | 20 | 6 | 507 |
| eICU | Wait without new contents | 2,492 | 60 | 0 | 85 | 10 |
| eICU | Wait for recorded arrival | 2,492 | 61 | 0 | 84 | 23 |

The same 2,492 requests and original six-hour target are used in all four conditions. Immediate retrieval reveals an existing old fact; waiting is capped at two hours and uses observed arrival times, not hypothetical test orders. Counts are timely organ choices, not sequential event capture.

#### Supplementary Table 24 Paired effects of retrieval and waiting

| **Policy** | **Comparator** | **Sampling unit** | **Difference (95% CI)** |
| --- | --- | --- | --- |
| Retrieve one existing fact | Immediate assessment | Patient | 14 (4 to 25) |
| Wait for recorded arrival | Immediate assessment | Patient | -84 (-102 to -67) |
| Wait for recorded arrival | Retrieve one existing fact | Patient | -98 (-118.00 to -79) |
| Wait for recorded arrival | Wait without new contents | Patient | 1 (0 to 3) |
| Retrieve one existing fact | Immediate assessment | Hospital | 14 (3.91 to 23.82) |
| Wait for recorded arrival | Immediate assessment | Hospital | -84 (-111.07 to -57.51) |
| Wait for recorded arrival | Retrieve one existing fact | Hospital | -98 (-125.25 to -69.31) |
| Wait for recorded arrival | Wait without new contents | Hospital | 1 (0 to 3.35) |

Differences in timely correct organ choices use 1,200 paired patient or hospital resamples of the same fixed requests.

#### Supplementary Table 25 Fixed earlier history and accumulated new evidence

| **Database** | **Condition** | **Information** | **Log loss** | **Brier score** | **Events captured** |
| --- | --- | --- | --- | --- | --- |
| MIMIC-IV | All post-index assessments | No fixed early history | 0.07686 | 0.01921 | 334 |
| MIMIC-IV | No new low value | No fixed early history | 0.11313 | 0.02880 | 237 |
| MIMIC-IV | One new low value | No fixed early history | 0.07787 | 0.01985 | 148 |
| MIMIC-IV | Repeated low values ≥1 h apart | No fixed early history | 0.09357 | 0.02603 | 150 |
| MIMIC-IV | Repeated low values <1 h apart | No fixed early history | 0.09838 | 0.02772 | 7 |
| eICU | All post-index assessments | No fixed early history | 0.05204 | 0.01074 | 177 |
| eICU | No new low value | No fixed early history | 0.08870 | 0.01983 | 185 |
| eICU | One new low value | No fixed early history | 0.04327 | 0.00853 | 86 |
| eICU | Repeated low values ≥1 h apart | No fixed early history | 0.06683 | 0.01352 | 62 |
| eICU | Repeated low values <1 h apart | No fixed early history | 0.07378 | 0.01471 | 15 |
| MIMIC-IV | All post-index assessments | Full fixed early history | 0.07655 | 0.01928 | 321 |
| MIMIC-IV | No new low value | Full fixed early history | 0.11036 | 0.02861 | 247 |
| MIMIC-IV | One new low value | Full fixed early history | 0.07933 | 0.02012 | 156 |
| MIMIC-IV | Repeated low values ≥1 h apart | Full fixed early history | 0.09391 | 0.02617 | 152 |
| MIMIC-IV | Repeated low values <1 h apart | Full fixed early history | 0.10246 | 0.02851 | 7 |
| eICU | All post-index assessments | Full fixed early history | 0.05234 | 0.01104 | 194 |
| eICU | No new low value | Full fixed early history | 0.08640 | 0.01997 | 199 |
| eICU | One new low value | Full fixed early history | 0.04228 | 0.00859 | 99 |
| eICU | Repeated low values ≥1 h apart | Full fixed early history | 0.06848 | 0.01406 | 55 |
| eICU | Repeated low values <1 h apart | Full fixed early history | 0.07990 | 0.01609 | 14 |
| MIMIC-IV | All post-index assessments | Retain fixed early history | 0.07655 | 0.01931 | 320 |
| MIMIC-IV | All post-index assessments | Halve early history after 12 hours | 0.07591 | 0.01921 | 320 |
| MIMIC-IV | All post-index assessments | Halve early history after new low measurements | 0.07630 | 0.01928 | 318 |
| MIMIC-IV | All post-index assessments | Step down early history after new low measurements | 0.07643 | 0.01929 | 317 |
| MIMIC-IV | No new low value | Retain fixed early history | 0.11113 | 0.02890 | 244 |
| MIMIC-IV | One new low value | Retain fixed early history | 0.07780 | 0.01995 | 150 |
| MIMIC-IV | Repeated low values ≥1 h apart | Retain fixed early history | 0.09362 | 0.02603 | 153 |
| MIMIC-IV | Repeated low values <1 h apart | Retain fixed early history | 0.09911 | 0.02775 | 7 |
| eICU | All post-index assessments | Retain fixed early history | 0.05196 | 0.01086 | 214 |
| eICU | All post-index assessments | Halve early history after 12 hours | 0.05140 | 0.01076 | 212 |
| eICU | All post-index assessments | Halve early history after new low measurements | 0.05153 | 0.01078 | 220 |
| eICU | All post-index assessments | Step down early history after new low measurements | 0.05148 | 0.01077 | 222 |
| eICU | No new low value | Retain fixed early history | 0.08566 | 0.01966 | 195 |
| eICU | One new low value | Retain fixed early history | 0.04291 | 0.00859 | 93 |
| eICU | Repeated low values ≥1 h apart | Retain fixed early history | 0.06813 | 0.01383 | 54 |
| eICU | Repeated low values <1 h apart | Retain fixed early history | 0.07839 | 0.01557 | 14 |

Both predictors share 225 current, observation and subsequent-information inputs. Added history is fixed strictly before index. Each evidence stratum has its own risk set and alert budget; captures are not additive across strata. This analysis uses scheduled post-index assessments, not the hourly main workflow.

#### Supplementary Table 26 Transfer of the earlier-history weighting rule

| **Database** | **History representation** | **Information** | **Patients** | **Log loss** | **Brier score** | **Events captured** |
| --- | --- | --- | --- | --- | --- | --- |
| CareVue | Original target-organ history | Retain early history | 1,269 | 0.09291 | 0.02117 | 104 |
| CareVue | Original target-organ history | No fixed early history | 1,269 | 0.09084 | 0.02100 | 118 |
| CareVue | Original target-organ history | Step down early history | 1,269 | 0.09192 | 0.02104 | 110 |
| CareVue | Compact fixed history | Retain early history | 1,269 | 0.09304 | 0.02120 | 119 |
| CareVue | Compact fixed history | No fixed early history | 1,269 | 0.09085 | 0.02104 | 118 |
| CareVue | Compact fixed history | Step down early history | 1,269 | 0.09243 | 0.02112 | 121 |
| SICdb | Original target-organ history | Retain early history | 500 | 0.09606 | 0.02387 | 37 |
| SICdb | Original target-organ history | No fixed early history | 500 | 0.08821 | 0.02158 | 29 |
| SICdb | Original target-organ history | Step down early history | 500 | 0.09052 | 0.02274 | 36 |
| SICdb | Compact fixed history | Retain early history | 500 | 0.10162 | 0.02597 | 28 |
| SICdb | Compact fixed history | No fixed early history | 500 | 0.08901 | 0.02197 | 29 |
| SICdb | Compact fixed history | Step down early history | 500 | 0.09514 | 0.02446 | 29 |

The rule was selected using MIMIC-IV and eICU, then applied unchanged. The compact version adds 96 fixed historical variables. The step rule progressively reduces their prediction weight. Scheduled post-index assessments and six-hour suppression are used.

#### Supplementary Table 27 Neurological ascertainment using observed score ranges

| **Database** | **Information** | **Patients** | **AUROC** | **Log loss** | **Brier score** | **Events captured** |
| --- | --- | --- | --- | --- | --- | --- |
| MIMIC-IV | Current information | 1,376 | 0.8968 | 0.07472 | 0.01821 | 220 |
| eICU | Current information | 4,983 | 0.8349 | 0.05484 | 0.01133 | 210 |
| MIMIC-IV | Compact history | 1,376 | 0.9153 | 0.07193 | 0.01806 | 235 |
| eICU | Compact history | 4,983 | 0.8640 | 0.05269 | 0.01131 | 260 |
| MIMIC-IV | Full history | 1,376 | 0.9146 | 0.07179 | 0.01803 | 235 |
| eICU | Full history | 4,983 | 0.8613 | 0.05308 | 0.01133 | 264 |

Nine scheduled assessments with six-hour patient suppression; six-hour outcome unless otherwise indicated. Models were fitted on the alternative ascertainment definition among patients retaining the original qualifying entry. This is not a new global search for first entry. The eICU cohort is unchanged.

#### Supplementary Table 28 Application to pancreatitis cohorts without retraining

| **Database** | **Horizon (h)** | **Information** | **Patients** | **AUROC** | **Log loss** | **Brier score** | **Events captured** |
| --- | --- | --- | --- | --- | --- | --- | --- |
| MIMIC-IV | 6 | Current information | 1,043 | 0.9120 | 0.07663 | 0.01934 | 189 |
| MIMIC-IV | 6 | Compact history | 1,043 | 0.9140 | 0.07622 | 0.01917 | 189 |
| MIMIC-IV | 6 | Full history | 1,043 | 0.9150 | 0.07612 | 0.01915 | 190 |
| MIMIC-IV | 12 | Current information | 1,043 | 0.9000 | 0.13517 | 0.03712 | 221 |
| MIMIC-IV | 12 | Compact history | 1,043 | 0.9040 | 0.13490 | 0.03691 | 226 |
| MIMIC-IV | 12 | Full history | 1,043 | 0.9027 | 0.13597 | 0.03707 | 229 |
| MIMIC-IV | 24 | Current information | 1,043 | 0.8734 | 0.20448 | 0.05800 | 245 |
| MIMIC-IV | 24 | Compact history | 1,043 | 0.8793 | 0.20236 | 0.05713 | 256 |
| MIMIC-IV | 24 | Full history | 1,043 | 0.8767 | 0.20407 | 0.05745 | 246 |
| eICU | 6 | Current information | 618 | 0.8012 | 0.04810 | 0.00930 | 20 |
| eICU | 6 | Compact history | 618 | 0.8347 | 0.04629 | 0.00917 | 29 |
| eICU | 6 | Full history | 618 | 0.8342 | 0.04632 | 0.00927 | 27 |
| eICU | 12 | Current information | 618 | 0.7950 | 0.09386 | 0.02099 | 29 |
| eICU | 12 | Compact history | 618 | 0.8172 | 0.09153 | 0.02069 | 32 |
| eICU | 12 | Full history | 618 | 0.8164 | 0.09095 | 0.02051 | 33 |
| eICU | 24 | Current information | 618 | 0.8060 | 0.14963 | 0.03745 | 38 |
| eICU | 24 | Compact history | 618 | 0.8133 | 0.15042 | 0.03814 | 39 |
| eICU | 24 | Full history | 618 | 0.8099 | 0.15084 | 0.03798 | 38 |

The sepsis models, representations and calibration are unchanged. Hourly reassessment and four-hour suppression are applied in separately identified pancreatitis cohorts, excluding patients shared with sepsis development or evaluation. Diagnosis/enzyme eligibility is specified in the main Methods under Supporting comparisons.

#### Supplementary Table 29 Sequential requests for additional record sources

| **Database** | **Policy** | **Additional accesses** | **Alerts** | **Events captured** | **Log loss** | **Brier score** |
| --- | --- | --- | --- | --- | --- | --- |
| MIMIC-IV | Two-step correction value | 1,960 | 980 | 437 | 0.08021 | 0.02039 |
| MIMIC-IV | One-step correction value | 1,947 | 980 | 444 | 0.08036 | 0.02040 |
| MIMIC-IV | Expected entropy reduction | 1,960 | 980 | 440 | 0.08029 | 0.02040 |
| MIMIC-IV | Risk priority | 1,960 | 980 | 439 | 0.08074 | 0.02050 |
| MIMIC-IV | Uncertainty priority | 1,960 | 980 | 437 | 0.08044 | 0.02048 |
| MIMIC-IV | Risk priority, matched request budget | 1,960 | 980 | 439 | 0.08074 | 0.02050 |
| MIMIC-IV | Uncertainty priority, matched request budget | 1,960 | 980 | 437 | 0.08044 | 0.02048 |
| MIMIC-IV | Current information | 0 | 980 | 433 | 0.08307 | 0.02071 |
| MIMIC-IV | Brief history | 0 | 980 | 433 | 0.08050 | 0.02049 |
| MIMIC-IV | Compact history | 9,800 | 980 | 448 | 0.07989 | 0.02035 |
| MIMIC-IV | Full history | 11,760 | 980 | 434 | 0.07934 | 0.02027 |
| MIMIC-IV | Union of brief and compact sources | 9,800 | 980 | 438 | 0.07972 | 0.02034 |
| eICU | Two-step correction value | 4,983 | 2,492 | 300 | 0.05331 | 0.01152 |
| eICU | One-step correction value | 4,930 | 2,492 | 304 | 0.05330 | 0.01153 |
| eICU | Expected entropy reduction | 4,983 | 2,492 | 304 | 0.05303 | 0.01144 |
| eICU | Risk priority | 4,983 | 2,492 | 297 | 0.05328 | 0.01152 |
| eICU | Uncertainty priority | 4,983 | 2,492 | 306 | 0.05330 | 0.01153 |
| eICU | Risk priority, matched request budget | 4,983 | 2,492 | 297 | 0.05328 | 0.01152 |
| eICU | Uncertainty priority, matched request budget | 4,983 | 2,492 | 306 | 0.05330 | 0.01153 |
| eICU | Current information | 0 | 2,492 | 245 | 0.05483 | 0.01145 |
| eICU | Brief history | 0 | 2,492 | 312 | 0.05318 | 0.01152 |
| eICU | Compact history | 24,915 | 2,492 | 319 | 0.05252 | 0.01130 |
| eICU | Full history | 29,898 | 2,492 | 314 | 0.05288 | 0.01148 |
| eICU | Union of brief and compact sources | 24,915 | 2,492 | 313 | 0.05316 | 0.01153 |
| CareVue | Two-step correction value | 1,288 | 644 | 137 | 0.09850 | 0.02291 |
| CareVue | One-step correction value | 1,288 | 644 | 133 | 0.09868 | 0.02297 |
| CareVue | Expected entropy reduction | 1,288 | 644 | 138 | 0.09915 | 0.02304 |
| CareVue | Risk priority | 1,288 | 644 | 132 | 0.09897 | 0.02304 |
| CareVue | Uncertainty priority | 1,288 | 644 | 137 | 0.09871 | 0.02299 |
| CareVue | Risk priority, matched request budget | 1,288 | 644 | 132 | 0.09897 | 0.02304 |
| CareVue | Uncertainty priority, matched request budget | 1,288 | 644 | 137 | 0.09871 | 0.02299 |
| CareVue | Current information | 0 | 644 | 107 | 0.10255 | 0.02335 |
| CareVue | Brief history | 0 | 644 | 134 | 0.09846 | 0.02298 |
| CareVue | Compact history | 6,440 | 644 | 144 | 0.09965 | 0.02297 |
| CareVue | Full history | 7,728 | 644 | 145 | 0.09985 | 0.02299 |
| CareVue | Union of brief and compact sources | 6,440 | 644 | 131 | 0.09949 | 0.02300 |
| SICdb | Two-step correction value | 514 | 257 | 32 | 0.09569 | 0.02437 |
| SICdb | One-step correction value | 514 | 257 | 32 | 0.09638 | 0.02459 |
| SICdb | Expected entropy reduction | 514 | 257 | 35 | 0.09427 | 0.02374 |
| SICdb | Risk priority | 514 | 257 | 27 | 0.09574 | 0.02429 |
| SICdb | Uncertainty priority | 514 | 257 | 31 | 0.09774 | 0.02516 |
| SICdb | Risk priority, matched request budget | 514 | 257 | 27 | 0.09574 | 0.02429 |
| SICdb | Uncertainty priority, matched request budget | 514 | 257 | 31 | 0.09774 | 0.02516 |
| SICdb | Current information | 0 | 257 | 27 | 0.09118 | 0.02234 |
| SICdb | Brief history | 0 | 257 | 31 | 0.09799 | 0.02512 |
| SICdb | Compact history | 2,570 | 257 | 49 | 0.09263 | 0.02310 |
| SICdb | Full history | 3,084 | 257 | 39 | 0.09311 | 0.02361 |
| SICdb | Union of brief and compact sources | 2,570 | 257 | 34 | 0.09702 | 0.02505 |

Hourly reassessment with four-hour patient suppression and a six-hour outcome. All methods receive the same alert budget. Each additional access opens a specified historical source for subsequent updates. Active policies begin with brief history and the common current inputs. Fixed-representation comparators are shown separately. Additional accesses count sources beyond brief history: compact history requires five additions because it does not contain lactate history, whereas full history requires six. This access unit differs from revealing one existing fact.

#### Supplementary Table 30 Event scheduling with full sequential constraints

| **Database** | **Alerts** | **Risk ordering** | **Opportunity-cost ordering** | **Difference (95% CI)** | **Hindsight bound** |
| --- | --- | --- | --- | --- | --- |
| MIMIC-IV | 980 | 448 | 451 | 3 (-11 to 13) | 980 |
| eICU | 2,492 | 319 | 320 | 1 (-7 to 7) | 999 |
| CareVue | 644 | 144 | 142 | -2 (-6 to 4) | 552 |
| SICdb | 257 | 49 | 48 | -1 (-7.52 to 8) | 225 |

Hourly reassessment with four-hour patient suppression and a six-hour outcome. All methods receive the same alert budget. The selected opportunity-cost rule is compared with risk ordering using compact history. The hindsight bound uses future labels and is solely a diagnostic upper bound, not a usable prediction policy.

#### Supplementary Table 31 Numerical history and the recording process

| **Database** | **Information** | **AUROC** | **Log loss** | **Brier score** | **Events captured** |
| --- | --- | --- | --- | --- | --- |
| MIMIC-IV | Current values and states | 0.8952 | 0.08266 | 0.02060 | 362 |
| MIMIC-IV | Current values, states and recording process | 0.8979 | 0.08219 | 0.02056 | 363 |
| MIMIC-IV | Current values and states + numerical history | 0.9041 | 0.08097 | 0.02045 | 365 |
| MIMIC-IV | Current assessment + numerical history | 0.9035 | 0.08115 | 0.02052 | 368 |
| MIMIC-IV | Full history | 0.9145 | 0.07867 | 0.02014 | 381 |
| eICU | Current values and states | 0.8432 | 0.05450 | 0.01139 | 238 |
| eICU | Current values, states and recording process | 0.8418 | 0.05406 | 0.01128 | 219 |
| eICU | Current values and states + numerical history | 0.8535 | 0.05340 | 0.01133 | 235 |
| eICU | Current assessment + numerical history | 0.8496 | 0.05378 | 0.01141 | 240 |
| eICU | Full history | 0.8706 | 0.05203 | 0.01130 | 282 |
| CareVue | Current values and states | 0.8261 | 0.10141 | 0.02292 | 98 |
| CareVue | Current values, states and recording process | 0.8380 | 0.10094 | 0.02298 | 108 |
| CareVue | Current values and states + numerical history | 0.8496 | 0.09889 | 0.02276 | 114 |
| CareVue | Current assessment + numerical history | 0.8524 | 0.09886 | 0.02275 | 116 |
| CareVue | Full history | 0.8514 | 0.09864 | 0.02265 | 117 |
| SICdb | Current values and states | 0.8543 | 0.09252 | 0.02242 | 29 |
| SICdb | Current values, states and recording process | 0.8591 | 0.09360 | 0.02317 | 26 |
| SICdb | Current values and states + numerical history | 0.8736 | 0.09251 | 0.02371 | 36 |
| SICdb | Current assessment + numerical history | 0.8734 | 0.09459 | 0.02468 | 36 |
| SICdb | Full history | 0.8698 | 0.09548 | 0.02437 | 40 |

Nine scheduled assessments with six-hour patient suppression; six-hour outcome unless otherwise indicated. C50 contains current information; C67 adds 17 recording-process expressions; V90 adds 90 numerical-history expressions; the full reference also includes 18 remaining organ-state expressions. Missing values can themselves carry recording-process information.

#### Supplementary Table 32 Clinical scores and recorded-state comparators

| **Database** | **Information** | **AUROC** | **Log loss** | **Brier score** | **Events captured** |
| --- | --- | --- | --- | --- | --- |
| MIMIC-IV | Full history | 0.9145 | 0.07867 | 0.02014 | 381 |
| MIMIC-IV | Current SOFA total | 0.6506 | 0.11011 | 0.02353 | 109 |
| MIMIC-IV | Current SOFA components | 0.7771 | 0.10245 | 0.02317 | 130 |
| MIMIC-IV | Current organ states | 0.7079 | 0.10726 | 0.02334 | 126 |
| eICU | Full history | 0.8706 | 0.05203 | 0.01130 | 282 |
| eICU | Current SOFA total | 0.4747 | 0.06830 | 0.01190 | 84 |
| eICU | Current SOFA components | 0.7476 | 0.06224 | 0.01189 | 94 |
| eICU | Current organ states | 0.5657 | 0.06769 | 0.01197 | 85 |
| CareVue | Full history | 0.8514 | 0.09864 | 0.02265 | 117 |
| CareVue | Current SOFA total | 0.5314 | 0.11433 | 0.02359 | 100 |
| CareVue | Current SOFA components | 0.7490 | 0.10522 | 0.02321 | 86 |
| CareVue | Current organ states | 0.6387 | 0.11216 | 0.02346 | 85 |
| SICdb | Full history | 0.8698 | 0.09548 | 0.02437 | 40 |
| SICdb | Current SOFA total | 0.7923 | 0.09850 | 0.02234 | 61 |
| SICdb | Current SOFA components | 0.8260 | 0.09558 | 0.02257 | 33 |
| SICdb | Current organ states | 0.7886 | 0.09990 | 0.02246 | 49 |

Nine scheduled assessments with six-hour patient suppression; six-hour outcome unless otherwise indicated. These are the clinical-score predictions calibrated using MIMIC-IV and used in the matched comparison in Supplementary Table 18. Local organ calibration is shown there separately. The total-score comparator falls back to a source organ prior when the score is unavailable.

#### Supplementary Table 33 Visible and generated histories with matched summaries

| **Database / records** | **History** | **Log loss** | **Brier** | **AUROC** | **Captures** |
| --- | --- | --- | --- | --- | --- |
| MIMIC-IV / Complete records | Visible history summaries | 0.078748 | 0.020136 | 0.9135 | 378.0 |
| MIMIC-IV / Complete records | Generated history summaries | 0.079008 | 0.020134 | 0.9120 | 382.0 |
| MIMIC-IV / Half of batches withheld | Visible history summaries | 0.084910 | 0.020998 | 0.8853 | 348.0 |
| MIMIC-IV / Half of batches withheld | Generated history summaries | 0.085056 | 0.020984 | 0.8842 | 346.0 |
| eICU / Complete records | Visible history summaries | 0.052540 | 0.011236 | 0.8657 | 269.0 |
| eICU / Complete records | Generated history summaries | 0.052380 | 0.011199 | 0.8647 | 271.0 |
| eICU / Half of batches withheld | Visible history summaries | 0.057885 | 0.011894 | 0.8138 | 215.0 |
| eICU / Half of batches withheld | Generated history summaries | 0.057570 | 0.011765 | 0.8113 | 217.7 |

Nine scheduled assessments with six-hour patient suppression; six-hour outcome unless otherwise indicated. The visible and generated alternatives use the same 18 summary expressions. These supporting comparisons are separate from the record-source compression search. Both readers used 639 factual inputs and the same 18 summary definitions, with equal training exposure. eICU withheld-record values average three assignments; the MIMIC-IV comparison used one assignment.

#### Supplementary Table 34 Alternative uses of historical trajectories

| **Comparison** | **Outcome** | **Log loss difference (95% CI)** | **Brier difference (95% CI)** | **Capture difference (95% CI)** |
| --- | --- | --- | --- | --- |
| Conditional unmasking with trees versus Visible sequence model | Organ event | −0.000228 (−0.000381 to −0.000109) | −0.000005 (−0.000017 to 0.000008) | 0.67 (−4.00 to 5.67) |
| Treeffuser versus Visible sequence model | Organ event | −0.000207 (−0.000369 to −0.000084) | −0.000004 (−0.000016 to 0.000009) | 0.33 (−4.00 to 5.33) |
| Whole history donor sampling versus Visible sequence model | Organ event | −0.000160 (−0.000321 to −0.000037) | 0.000002 (−0.000011 to 0.000014) | 2.00 (−4.00 to 6.33) |
| Honest regression tree sampling versus Visible sequence model | Organ event | −0.000087 (−0.000183 to −0.000021) | 0.000001 (−0.000006 to 0.000007) | 2.67 (−1.51 to 7.67) |
| Distributional tree sampling versus Visible sequence model | Organ event | −0.000131 (−0.000290 to −0.000010) | 0.000005 (−0.000008 to 0.000018) | 2.00 (−3.67 to 7.67) |
| Independent conditional sampling versus Visible sequence model | Organ event | −0.000230 (−0.000383 to −0.000113) | −0.000006 (−0.000019 to 0.000007) | 1.00 (−4.00 to 6.00) |
| Organ history teacher versus Current-information teacher | Organ event | −0.000067 (−0.000216 to 0.000080) | −0.000016 (−0.000050 to 0.000014) | 1.00 (−5.00 to 11.00) |
| Organ history teacher versus No teacher | Organ event | −0.000249 (−0.000507 to 0.000021) | −0.000081 (−0.000133 to −0.000028) | 13.33 (2.00 to 24.01) |
| Organ history teacher versus Visible sequence model | Organ event | 0.000111 (−0.000253 to 0.000491) | −0.000037 (−0.000087 to 0.000021) | 9.33 (−2.51 to 21.67) |
| Pairwise history teacher versus Current-information teacher | Organ event | −0.000135 (−0.000261 to −0.000007) | 0.000000 (−0.000010 to 0.000010) | 3.67 (−5.00 to 9.67) |
| Pairwise history teacher versus No teacher | Organ event | −0.000918 (−0.001477 to −0.000346) | −0.000131 (−0.000164 to −0.000099) | 24.00 (9.67 to 36.01) |
| Pairwise history teacher versus Visible sequence model | Organ event | 0.002505 (0.001365 to 0.003781) | −0.000209 (−0.000387 to 0.000021) | −34.33 (−54.00 to −16.33) |
| Patient event time history teacher versus Current-information teacher | Patient event time | −0.002719 (−0.003362 to −0.002075) | −0.000600 (−0.000726 to −0.000466) | −9.67 (−21.00 to −0.67) |
| Patient event time history teacher versus Outcome labels alone | Patient event time | −0.001986 (−0.002622 to −0.001347) | −0.000536 (−0.000649 to −0.000413) | −7.33 (−17.33 to 2.00) |
| Patient event time history teacher versus Visible sequence model | Patient event time | 0.000208 (−0.002029 to 0.002608) | −0.001471 (−0.001906 to −0.001025) | −72.00 (−91.17 to −51.83) |
| Trajectory expert combination versus Visible sequence model | Organ event | −0.000540 (−0.000719 to −0.000369) | −0.000067 (−0.000091 to −0.000043) | 1.67 (−4.67 to 7.00) |
| Contrastive objective for current inputs versus Same model without contrastive objective | Organ event | −0.000113 (−0.000931 to 0.000631) | 0.000115 (0.000061 to 0.000175) | −7.67 (−21.51 to 9.35) |
| Contrastive objective for trajectory experts versus Same model without contrastive objective | Organ event | 0.001237 (0.000792 to 0.001705) | 0.000163 (0.000127 to 0.000202) | 4.67 (−7.84 to 18.84) |
| Joint event process combination versus Visible sequence model | Organ event | −0.000307 (−0.000667 to 0.000039) | −0.000081 (−0.000120 to −0.000042) | 6.33 (−4.17 to 14.33) |
| Mean probability combination versus Visible sequence model | Organ event | −0.000959 (−0.001242 to −0.000683) | −0.000221 (−0.000272 to −0.000171) | 6.33 (−4.17 to 14.33) |
| Logistic combination versus Mean probability combination | Organ event | 0.000316 (0.000152 to 0.000496) | 0.000086 (0.000054 to 0.000122) | −3.33 (−10.51 to 3.33) |
| Convex combination versus Mean probability combination | Organ event | 0.000894 (0.000512 to 0.001253) | −0.000091 (−0.000156 to −0.000016) | −16.33 (−27.51 to −3.83) |
| Schrödinger bridge imputation versus Matched direct model | Organ event | 0.011638 (0.010574 to 0.012662) | 0.000461 (0.000309 to 0.000655) | −157.33 (−179.68 to −129.82) |
| History generator with recurrent reader versus Direct recurrent model | Organ event | 0.000104 (−0.000005 to 0.000210) | 0.000032 (0.000016 to 0.000049) | −5.67 (−10.67 to 0.67) |
| History generator with recurrent reader versus Visible sequence model | Organ event | 0.001926 (0.001018 to 0.002956) | 0.000050 (−0.000100 to 0.000263) | −42.33 (−59.51 to −26.32) |

Nine scheduled assessments with six-hour patient suppression; six-hour outcome unless otherwise indicated. Each row retains its actual matched comparator and information condition. Probability improvement and capture improvement are distinct outcomes. Source-stopped methods are not claimed to have completed four-database testing. Comparisons use eICU with half of record batches withheld and 2,492 alerts. Each row retains its own reference and prediction outcome. Patient intervals are computed within the specified comparison, averaging the three record-withholding assignments within each resample.

#### Supplementary Table 35 Historical representations in a separate norepinephrine decision task

| **Information** | **Capacity** | **Patients** | **Assigned patients** | **AUTOC** | **NE reduction (µg/kg)** | **Pressure-risk difference** |
| --- | --- | --- | --- | --- | --- | --- |
| Benefit ranking: current assessment and treatment context | 0.20 | 813 | 162 | 23.33 | 18.44 | -0.00352 |
| Benefit ranking: brief history | 0.20 | 813 | 162 | 17.61 | 14.02 | -0.06673 |
| Benefit ranking: compact history | 0.20 | 813 | 162 | 18.40 | 16.86 | -0.02414 |
| Benefit ranking: full history | 0.20 | 813 | 162 | 14.84 | 11.27 | -0.01514 |
| Risk ranking: full history | 0.20 | 813 | 162 | 24.62 | 17.30 | -0.02773 |
| Risk ranking: current assessment and treatment context | 0.20 | 813 | 162 | 20.02 | 13.98 | -0.03092 |
| Risk ranking: compact history | 0.20 | 813 | 162 | 22.75 | 13.72 | 0.00289 |

All rows use 813 first opportunities, the same treatment definition and matched NE background inputs. Benefit rules rank estimated treatment benefit; Prognosis rules rank predicted hypotension-or-death risk. Recorded-exposure reduction is in micrograms/kg; the pressure-risk difference is a proportion relative to deferral. Twenty per cent capacity assigns 162 patients. This is a separate endpoint and population from organ reassessment.

#### Supplementary Table 36 Resolution and permutation of generated histories

| **Comparison and target** | **Difference** | **Hospital 95% CI** |
| --- | --- | --- |
| Conditional history versus source prior: coarse-type log loss | −0.159960 | −0.178265 to −0.139830 |
| Conditional history versus source prior: coarse-type Brier | −0.074564 | −0.083530 to −0.064298 |
| Conditional history versus source prior: four-block log loss | 0.101385 | 0.055839 to 0.149912 |
| Conditional history versus source prior: four-block Brier | −0.045288 | −0.060083 to −0.028876 |
| Intact versus permuted history: generation-branch log loss | −0.002872 | −0.004408 to −0.001252 |
| Intact versus permuted history: final-mixture log loss | −0.000030 | −0.000369 to 0.000301 |
| Final mixture versus current information: log loss | −0.000513 | −0.000941 to −0.000154 |

This supporting reconstruction analysis uses a separate eICU cohort of 4,982 patients. Permutation preserved source role, current active organ and visible-duration strata, with the same predictor, 128 history samples and mixture weight 0.3; all three permutation seeds were included. Coarse-type 90% sets covered 89.11% with mean size 1.143, versus 92.03% and 1.674 for the source prior. Four-block sets covered 81.83% with mean size 4.098, versus 91.90% and 6.611. Generation-branch and final-mixture effects are distinct contrasts; these are not hourly-workflow performance estimates.

#### Supplementary Table 37 Observed lead times across prediction horizons

| **Database** | **Information** | **Horizon (h)** | **Events captured** | **Onset Q1 (h)** | **Onset median (h)** | **Onset Q3 (h)** | **Confirm. median (h)** |
| --- | --- | --- | --- | --- | --- | --- | --- |
| MIMIC-IV | Current information | 6 | 363 | 1 | 2 | 3 | 4 |
| MIMIC-IV | Compact history | 6 | 382 | 1 | 2 | 3 | 4 |
| MIMIC-IV | Full history | 6 | 381 | 1 | 2 | 3 | 4 |
| MIMIC-IV | Current information | 12 | 504 | 2 | 3 | 6 | 5 |
| MIMIC-IV | Compact history | 12 | 526 | 1 | 3 | 6 | 5 |
| MIMIC-IV | Full history | 12 | 516 | 1 | 3 | 6 | 5 |
| MIMIC-IV | Current information | 24 | 571 | 2 | 4 | 9 | 6 |
| MIMIC-IV | Compact history | 24 | 576 | 2 | 5 | 9 | 7 |
| MIMIC-IV | Full history | 24 | 568 | 2 | 4.50 | 9 | 6.50 |
| eICU | Current information | 6 | 219 | 1 | 2 | 3 | 4 |
| eICU | Compact history | 6 | 272 | 1 | 2 | 3 | 4 |
| eICU | Full history | 6 | 282 | 1 | 2 | 3 | 4 |
| eICU | Current information | 12 | 332 | 3 | 5.50 | 8 | 7.50 |
| eICU | Compact history | 12 | 391 | 2 | 5 | 7 | 7 |
| eICU | Full history | 12 | 418 | 2 | 5 | 7 | 7 |
| eICU | Current information | 24 | 400 | 4 | 9 | 17 | 11 |
| eICU | Compact history | 24 | 483 | 3 | 7 | 13.50 | 9 |
| eICU | Full history | 24 | 495 | 3 | 7 | 15 | 9 |
| CareVue | Current information | 6 | 108 | 1 | 2 | 3 | 4 |
| CareVue | Compact history | 6 | 126 | 1 | 2 | 3 | 4 |
| CareVue | Full history | 6 | 117 | 1 | 2 | 3 | 4 |
| CareVue | Current information | 12 | 170 | 3 | 5 | 7 | 7 |
| CareVue | Compact history | 12 | 193 | 2 | 4 | 6 | 6 |
| CareVue | Full history | 12 | 197 | 2 | 3 | 6 | 5 |
| CareVue | Current information | 24 | 226 | 4 | 7 | 13.75 | 9 |
| CareVue | Compact history | 24 | 243 | 2 | 6 | 11.50 | 8 |
| CareVue | Full history | 24 | 238 | 2 | 6 | 11 | 8 |
| SICdb | Current information | 6 | 26 | 1 | 2 | 3 | 4 |
| SICdb | Compact history | 6 | 46 | 1 | 1 | 2.75 | 3 |
| SICdb | Full history | 6 | 40 | 1 | 1 | 2 | 3 |
| SICdb | Current information | 12 | 57 | 3 | 5 | 7 | 7 |
| SICdb | Compact history | 12 | 68 | 2 | 5 | 7 | 7 |
| SICdb | Full history | 12 | 71 | 2 | 5 | 7 | 7 |
| SICdb | Current information | 24 | 48 | 5.75 | 10 | 17 | 12 |
| SICdb | Compact history | 24 | 88 | 2 | 7 | 13 | 9 |
| SICdb | Full history | 24 | 93 | 2 | 5 | 13 | 7 |

Lead time is calculated among distinct captured events under the nine-origin, six-hour-suppression protocol. Onset and three-hour confirmation are separate clocks. These medians do not describe all events or the hourly main workflow.

#### Supplementary Table 38 Paired comparisons of norepinephrine prioritisation

| **Information** | **Comparator** | **Capacity** | **Measure** | **Difference (95% CI)** |
| --- | --- | --- | --- | --- |
| Benefit ranking: brief history | Benefit ranking: current assessment and treatment context | Not applicable | AUTOC | -5.72 (-13.42 to 1.92) |
| Benefit ranking: brief history | Benefit ranking: current assessment and treatment context | 0.20 | Recorded NE reduction | -4.42 (-10.24 to 2.70) |
| Benefit ranking: brief history | Benefit ranking: current assessment and treatment context | 0.20 | Pressure risk | -0.06321 (-0.14413 to 0.02816) |
| Benefit ranking: compact history | Benefit ranking: current assessment and treatment context | Not applicable | AUTOC | -4.93 (-13.94 to 3.02) |
| Benefit ranking: compact history | Benefit ranking: current assessment and treatment context | 0.20 | Recorded NE reduction | -1.58 (-9.41 to 1.98) |
| Benefit ranking: compact history | Benefit ranking: current assessment and treatment context | 0.20 | Pressure risk | -0.02062 (-0.07759 to 0.08161) |
| Benefit ranking: full history | Benefit ranking: current assessment and treatment context | Not applicable | AUTOC | -8.49 (-19.53 to 0.87929) |
| Benefit ranking: full history | Benefit ranking: current assessment and treatment context | 0.20 | Recorded NE reduction | -7.18 (-14.00 to 0.45724) |
| Benefit ranking: full history | Benefit ranking: current assessment and treatment context | 0.20 | Pressure risk | -0.01162 (-0.10019 to 0.07509) |
| Benefit ranking: compact history | Benefit ranking: full history | Not applicable | AUTOC | 3.56 (-3.27 to 10.99) |
| Benefit ranking: compact history | Benefit ranking: full history | 0.20 | Recorded NE reduction | 5.59 (-2.68 to 11.33) |
| Benefit ranking: compact history | Benefit ranking: full history | 0.20 | Pressure risk | -0.00900 (-0.05909 to 0.08686) |
| Benefit ranking: brief history | Benefit ranking: full history | Not applicable | AUTOC | 2.77 (-3.00 to 10.26) |
| Benefit ranking: brief history | Benefit ranking: full history | 0.20 | Recorded NE reduction | 2.75 (-4.36 to 10.85) |
| Benefit ranking: brief history | Benefit ranking: full history | 0.20 | Pressure risk | -0.05159 (-0.13222 to 0.04339) |
| Benefit ranking: compact history | Risk ranking: compact history | Not applicable | AUTOC | -4.35 (-33.36 to 23.60) |
| Benefit ranking: compact history | Risk ranking: compact history | 0.20 | Recorded NE reduction | 3.14 (-9.13 to 11.42) |
| Benefit ranking: compact history | Risk ranking: compact history | 0.20 | Pressure risk | -0.02703 (-0.12505 to 0.08835) |
| Benefit ranking: full history | Risk ranking: full history | Not applicable | AUTOC | -9.78 (-39.94 to 17.50) |
| Benefit ranking: full history | Risk ranking: full history | 0.20 | Recorded NE reduction | -6.03 (-17.05 to 7.28) |
| Benefit ranking: full history | Risk ranking: full history | 0.20 | Pressure risk | 0.01260 (-0.14691 to 0.10782) |

AUTOC contrasts summarise the ranking curve; capacity is not applicable to those rows. Other rows compare policies assigning 20% of 813 patients. Intervals use 500 paired patient draws conditional on the fitted adjustment and decision models. Recorded-exposure units are micrograms/kg; pressure-risk differences are proportions.

#### Supplementary Table 39 Paired effects of compact history at six hours

| **Database** | **Current inputs** | **Comparison** | **Sampling unit** | **Measure** | **Difference (95% CI)** |
| --- | --- | --- | --- | --- | --- |
| MIMIC-IV | 67 | Compact − Current | Patient | Log loss | -0.00292 (-0.00409 to -0.00180) |
| MIMIC-IV | 67 | Compact − Current | Patient | Brier score | -0.00036 (-0.00058 to -0.00013) |
| MIMIC-IV | 67 | Compact − Current | Patient | AUROC | 0.01349 (0.00825 to 0.01917) |
| MIMIC-IV | 67 | Compact − Current | Patient | Events captured | 19 (-2 to 37) |
| MIMIC-IV | 67 | Full − Current | Patient | Log loss | -0.00352 (-0.00463 to -0.00248) |
| MIMIC-IV | 67 | Full − Current | Patient | Brier score | -0.00043 (-0.00062 to -0.00022) |
| MIMIC-IV | 67 | Full − Current | Patient | AUROC | 0.01651 (0.01139 to 0.02182) |
| MIMIC-IV | 67 | Full − Current | Patient | Events captured | 18 (-5 to 35) |
| MIMIC-IV | 67 | Compact − Full | Patient | Log loss | 0.00060 (0.00008 to 0.00112) |
| MIMIC-IV | 67 | Compact − Full | Patient | Brier score | 0.00006 (-0.00008 to 0.00023) |
| MIMIC-IV | 67 | Compact − Full | Patient | AUROC | -0.00303 (-0.00519 to -0.00105) |
| MIMIC-IV | 67 | Compact − Full | Patient | Events captured | 1 (-13 to 17.02) |
| MIMIC-IV | 91 | Compact − Current | Patient | Log loss | -0.00332 (-0.00435 to -0.00224) |
| MIMIC-IV | 91 | Compact − Current | Patient | Brier score | -0.00040 (-0.00061 to -0.00019) |
| MIMIC-IV | 91 | Compact − Current | Patient | AUROC | 0.01594 (0.01060 to 0.02173) |
| MIMIC-IV | 91 | Compact − Current | Patient | Events captured | 35 (7 to 57) |
| MIMIC-IV | 91 | Full − Current | Patient | Log loss | -0.00317 (-0.00431 to -0.00200) |
| MIMIC-IV | 91 | Full − Current | Patient | Brier score | -0.00037 (-0.00059 to -0.00014) |
| MIMIC-IV | 91 | Full − Current | Patient | AUROC | 0.01584 (0.01001 to 0.02219) |
| MIMIC-IV | 91 | Full − Current | Patient | Events captured | 13 (-10 to 37) |
| MIMIC-IV | 91 | Compact − Full | Patient | Log loss | -0.00016 (-0.00059 to 0.00029) |
| MIMIC-IV | 91 | Compact − Full | Patient | Brier score | -0.00003 (-0.00013 to 0.00006) |
| MIMIC-IV | 91 | Compact − Full | Patient | AUROC | 0.00010 (-0.00168 to 0.00190) |
| MIMIC-IV | 91 | Compact − Full | Patient | Events captured | 22 (4 to 34) |
| eICU | 67 | Compact − Current | Patient | Log loss | -0.00232 (-0.00327 to -0.00143) |
| eICU | 67 | Compact − Current | Patient | Brier score | -0.00013 (-0.00029 to -3.70e-06) |
| eICU | 67 | Compact − Current | Patient | AUROC | 0.02765 (0.01883 to 0.03626) |
| eICU | 67 | Compact − Current | Patient | Events captured | 53 (23 to 81.02) |
| eICU | 67 | Full − Current | Patient | Log loss | -0.00202 (-0.00305 to -0.00105) |
| eICU | 67 | Full − Current | Patient | Brier score | 0.00002 (-0.00015 to 0.00016) |
| eICU | 67 | Full − Current | Patient | AUROC | 0.02880 (0.01920 to 0.03819) |
| eICU | 67 | Full − Current | Patient | Events captured | 63 (34 to 91) |
| eICU | 67 | Compact − Full | Patient | Log loss | -0.00030 (-0.00062 to 0.00005) |
| eICU | 67 | Compact − Full | Patient | Brier score | -0.00016 (-0.00024 to -0.00007) |
| eICU | 67 | Compact − Full | Patient | AUROC | -0.00115 (-0.00423 to 0.00167) |
| eICU | 67 | Compact − Full | Patient | Events captured | -10 (-28 to 11) |
| eICU | 67 | Compact − Current | Hospital | Log loss | -0.00232 (-0.00347 to -0.00119) |
| eICU | 67 | Compact − Current | Hospital | Brier score | -0.00013 (-0.00030 to 0.00001) |
| eICU | 67 | Compact − Current | Hospital | AUROC | 0.02765 (0.01696 to 0.03690) |
| eICU | 67 | Compact − Current | Hospital | Events captured | 53 (18.27 to 89.81) |
| eICU | 67 | Full − Current | Hospital | Log loss | -0.00202 (-0.00317 to -0.00088) |
| eICU | 67 | Full − Current | Hospital | Brier score | 0.00002 (-0.00017 to 0.00018) |
| eICU | 67 | Full − Current | Hospital | AUROC | 0.02880 (0.01747 to 0.03830) |
| eICU | 67 | Full − Current | Hospital | Events captured | 63 (25.64 to 99.50) |
| eICU | 67 | Compact − Full | Hospital | Log loss | -0.00030 (-0.00063 to 0.00001) |
| eICU | 67 | Compact − Full | Hospital | Brier score | -0.00016 (-0.00023 to -0.00009) |
| eICU | 67 | Compact − Full | Hospital | AUROC | -0.00115 (-0.00408 to 0.00141) |
| eICU | 67 | Compact − Full | Hospital | Events captured | -10 (-27.25 to 7.34) |
| eICU | 91 | Compact − Current | Patient | Log loss | -0.00162 (-0.00252 to -0.00077) |
| eICU | 91 | Compact − Current | Patient | Brier score | -4.77e-06 (-0.00016 to 0.00015) |
| eICU | 91 | Compact − Current | Patient | AUROC | 0.02418 (0.01680 to 0.03230) |
| eICU | 91 | Compact − Current | Patient | Events captured | 65 (33 to 92.02) |
| eICU | 91 | Full − Current | Patient | Log loss | -0.00186 (-0.00278 to -0.00102) |
| eICU | 91 | Full − Current | Patient | Brier score | -0.00005 (-0.00021 to 0.00008) |
| eICU | 91 | Full − Current | Patient | AUROC | 0.02615 (0.01776 to 0.03526) |
| eICU | 91 | Full − Current | Patient | Events captured | 69 (41 to 96) |
| eICU | 91 | Compact − Full | Patient | Log loss | 0.00024 (-0.00008 to 0.00057) |
| eICU | 91 | Compact − Full | Patient | Brier score | 0.00005 (-0.00002 to 0.00012) |
| eICU | 91 | Compact − Full | Patient | AUROC | -0.00197 (-0.00514 to 0.00096) |
| eICU | 91 | Compact − Full | Patient | Events captured | -4 (-24 to 11) |
| eICU | 91 | Compact − Current | Hospital | Log loss | -0.00162 (-0.00276 to -0.00043) |
| eICU | 91 | Compact − Current | Hospital | Brier score | -4.77e-06 (-0.00018 to 0.00017) |
| eICU | 91 | Compact − Current | Hospital | AUROC | 0.02418 (0.01454 to 0.03446) |
| eICU | 91 | Compact − Current | Hospital | Events captured | 65 (29.03 to 85.82) |
| eICU | 91 | Full − Current | Hospital | Log loss | -0.00186 (-0.00292 to -0.00072) |
| eICU | 91 | Full − Current | Hospital | Brier score | -0.00005 (-0.00023 to 0.00011) |
| eICU | 91 | Full − Current | Hospital | AUROC | 0.02615 (0.01624 to 0.03748) |
| eICU | 91 | Full − Current | Hospital | Events captured | 69 (39.09 to 93.11) |
| eICU | 91 | Compact − Full | Hospital | Log loss | 0.00024 (-0.00010 to 0.00065) |
| eICU | 91 | Compact − Full | Hospital | Brier score | 0.00005 (-0.00002 to 0.00012) |
| eICU | 91 | Compact − Full | Hospital | AUROC | -0.00197 (-0.00582 to 0.00103) |
| eICU | 91 | Compact − Full | Hospital | Events captured | -4 (-24.91 to 9.76) |

Nine scheduled assessments with six-hour patient suppression; six-hour outcome unless otherwise indicated. Candidate IDs 0, 16063 and 16383 are current, compact and full history. The 67- and 91-variable current assessments are separate comparisons. Intervals use 1,200 paired draws; the fitted representation and calibration remain fixed.

#### Supplementary Table 40 Paired effects of models applied without retraining in pancreatitis-related cohorts

| **Database** | **Information** | **Comparator** | **Sampling unit** | **Measure** | **Difference (95% CI)** |
| --- | --- | --- | --- | --- | --- |
| MIMIC-IV | Full history | Current information | Patient | Log loss | -0.00051 (-0.00184 to 0.00099) |
| MIMIC-IV | Full history | Current information | Patient | Brier score | -0.00019 (-0.00045 to 0.00005) |
| MIMIC-IV | Full history | Current information | Patient | AUROC | 0.00299 (-0.00352 to 0.00895) |
| MIMIC-IV | Full history | Current information | Patient | Events captured | 1 (-16 to 20.52) |
| MIMIC-IV | Full history | Compact history | Patient | Log loss | -0.00010 (-0.00075 to 0.00059) |
| MIMIC-IV | Full history | Compact history | Patient | Brier score | -0.00001 (-0.00019 to 0.00014) |
| MIMIC-IV | Full history | Compact history | Patient | AUROC | 0.00094 (-0.00173 to 0.00390) |
| MIMIC-IV | Full history | Compact history | Patient | Events captured | 1 (-16 to 10) |
| MIMIC-IV | Compact history | Current information | Patient | Log loss | -0.00041 (-0.00178 to 0.00107) |
| MIMIC-IV | Compact history | Current information | Patient | Brier score | -0.00018 (-0.00040 to 0.00008) |
| MIMIC-IV | Compact history | Current information | Patient | AUROC | 0.00205 (-0.00549 to 0.00832) |
| MIMIC-IV | Compact history | Current information | Patient | Events captured | 0 (-12 to 23) |
| eICU | Full history | Current information | Patient | Log loss | -0.00178 (-0.00325 to -0.00040) |
| eICU | Full history | Current information | Patient | Brier score | -0.00003 (-0.00018 to 0.00014) |
| eICU | Full history | Current information | Patient | AUROC | 0.03298 (0.01012 to 0.05800) |
| eICU | Full history | Current information | Patient | Events captured | 7 (-2 to 17) |
| eICU | Full history | Compact history | Patient | Log loss | 0.00004 (-0.00065 to 0.00070) |
| eICU | Full history | Compact history | Patient | Brier score | 0.00011 (-2.81e-06 to 0.00024) |
| eICU | Full history | Compact history | Patient | AUROC | -0.00055 (-0.00921 to 0.01044) |
| eICU | Full history | Compact history | Patient | Events captured | -2 (-6.52 to 6) |
| eICU | Full history | Current information | Hospital | Log loss | -0.00178 (-0.00311 to -0.00041) |
| eICU | Full history | Current information | Hospital | Brier score | -0.00003 (-0.00018 to 0.00014) |
| eICU | Full history | Current information | Hospital | AUROC | 0.03298 (0.00748 to 0.05410) |
| eICU | Full history | Current information | Hospital | Events captured | 7 (-0.52079 to 15.02) |
| eICU | Full history | Compact history | Hospital | Log loss | 0.00004 (-0.00057 to 0.00060) |
| eICU | Full history | Compact history | Hospital | Brier score | 0.00011 (0.00001 to 0.00022) |
| eICU | Full history | Compact history | Hospital | AUROC | -0.00055 (-0.00898 to 0.00903) |
| eICU | Full history | Compact history | Hospital | Events captured | -2 (-5.83 to 5.75) |
| eICU | Compact history | Current information | Patient | Log loss | -0.00181 (-0.00329 to -0.00039) |
| eICU | Compact history | Current information | Patient | Brier score | -0.00014 (-0.00028 to -6.88e-06) |
| eICU | Compact history | Current information | Patient | AUROC | 0.03353 (0.01307 to 0.05658) |
| eICU | Compact history | Current information | Patient | Events captured | 9 (0 to 17) |
| eICU | Compact history | Current information | Hospital | Log loss | -0.00181 (-0.00319 to -0.00028) |
| eICU | Compact history | Current information | Hospital | Brier score | -0.00014 (-0.00026 to -0.00001) |
| eICU | Compact history | Current information | Hospital | AUROC | 0.03353 (0.01169 to 0.05568) |
| eICU | Compact history | Current information | Hospital | Events captured | 9 (-0.49085 to 14.81) |

Hourly reassessment with four-hour patient suppression and a six-hour outcome. All methods receive the same alert budget. Six-hour results use 500 paired patient draws and additional hospital draws in eICU. Models are unchanged from sepsis. Diagnostic support is described separately from enzyme-only eligibility.

#### Supplementary Table 41 Threshold selection for lower-priority reassessment

| **Testing setting** | **Risk target (%)** | **Information** | **Patients tested** | **Threshold selected** | **Patients selected** |
| --- | --- | --- | --- | --- | --- |
| Source patients | 1 | Current information | 978 | No | 0 |
| Source patients | 1 | Brief history | 978 | No | 0 |
| Source patients | 1 | Compact history | 978 | No | 0 |
| Source patients | 1 | Full history | 978 | No | 0 |
| Source patients | 2 | Current information | 978 | No | 0 |
| Source patients | 2 | Brief history | 978 | No | 0 |
| Source patients | 2 | Compact history | 978 | No | 0 |
| Source patients | 2 | Full history | 978 | No | 0 |
| Source patients | 5 | Current information | 978 | No | 0 |
| Source patients | 5 | Brief history | 978 | No | 0 |
| Source patients | 5 | Compact history | 978 | No | 0 |
| Source patients | 5 | Full history | 978 | No | 0 |
| Target patients | 1 | Current information | 787 | No | 0 |
| Target patients | 1 | Brief history | 787 | No | 0 |
| Target patients | 1 | Compact history | 787 | No | 0 |
| Target patients | 1 | Full history | 787 | No | 0 |
| Target patients | 2 | Current information | 787 | No | 0 |
| Target patients | 2 | Brief history | 787 | No | 0 |
| Target patients | 2 | Compact history | 787 | No | 0 |
| Target patients | 2 | Full history | 787 | No | 0 |
| Target patients | 5 | Current information | 787 | No | 0 |
| Target patients | 5 | Brief history | 787 | No | 0 |
| Target patients | 5 | Compact history | 787 | No | 0 |
| Target patients | 5 | Full history | 787 | No | 0 |
| Target hospitals | 1 | Current information | 787 | No | 0 |
| Target hospitals | 1 | Brief history | 787 | No | 0 |
| Target hospitals | 1 | Compact history | 787 | No | 0 |
| Target hospitals | 1 | Full history | 787 | No | 0 |
| Target hospitals | 2 | Current information | 787 | No | 0 |
| Target hospitals | 2 | Brief history | 787 | No | 0 |
| Target hospitals | 2 | Compact history | 787 | No | 0 |
| Target hospitals | 2 | Full history | 787 | No | 0 |
| Target hospitals | 5 | Current information | 787 | No | 0 |
| Target hospitals | 5 | Brief history | 787 | No | 0 |
| Target hospitals | 5 | Compact history | 787 | No | 0 |
| Target hospitals | 5 | Full history | 787 | No | 0 |

The first available assessment contributes one decision per patient. Each model has 19 candidate thresholds. At each risk target and testing setting, multiplicity correction covers 76 model–threshold combinations. Patient analyses use binomial tests; the target-hospital analysis uses 23 hospital units. Failure includes an organ event, early death or unresolved six-hour follow-up. No threshold passed. These 36 rows are model–risk-target–setting evaluations, not 36 individual candidate thresholds.

#### Supplementary Table 42 Clinical interpretation of changed organ priorities

| **Prompt group** | **Episodes or alerts** | **Patients** | **Median paired risk change, pp** | **Organ rank improved / worsened** |
| --- | --- | --- | --- | --- |
| Newly captured with history | 122 | 117 | 10.29 | 43 / 0 |
| Captured only without history | 59 | 58 | −3.51 | 0 / 7 |
| Captured by both | 160 | 152 | 2.87 | 0 / 0 |
| History alerts without an event | 2,210 | 1,512 | 6.16 | 366 / 0 |

Scheduled own-organ-history comparison: 67 current columns versus target-organ history, nine assessment origins and six-hour patient suppression. Risk changes are percentage points. Time since abnormality belongs to clinical history; measurement age belongs to recording process. Signed net contributions are distinct from summed absolute contributions.

#### Supplementary Table 43 Urine recording around catheter removal

| **Quantity** | **Reported result** |
| --- | --- |
| Removal episodes / patients | 26 / 25 |
| Mean collections in preceding six hours | 3.23 |
| Mean collections in subsequent six hours | 1.58 |
| Paired change in collection count (95% CI) | −1.65 (−2.30 to −1.00) |
| Change per six observed ICU hours (95% CI) | −1.61 (−2.25 to −0.96) |

Distinct urine collections in the six hours before and after documented catheter removal, requiring preceding catheter evidence and excluding unusable times and irrigation. Subsequent collections were not required. Paired intervals used 2,000 patient resamples; standardisation used observed ICU time until discharge or death.

#### Supplementary Table 44 Local calibration in the hourly reassessment workflow

| **Database** | **Patients / alerts** | **Captures, source → local** | **Capture change (95% CI)** | **Log loss, source → local** |
| --- | --- | --- | --- | --- |
| MIMIC-IV | 1960 / 980 | 448 → 448 | 0.00 (0.00 to 0.00) | 0.07989 → 0.07989 |
| eICU | 4196 / 2098 | 263 → 269 | 6.00 (-7.53 to 21.00) | 0.05286 → 0.05193 |
| CareVue | 1288 / 644 | 144 → 140 | -4.00 (-18.53 to 15.00) | 0.09965 → 0.09344 |
| SICdb | 514 / 257 | 49 → 72 | 23.00 (8.00 to 38.00) | 0.09263 → 0.08304 |

Ten-source history; six-hour outcome, hourly refresh and four-hour patient cooldown. eICU uses 23 adaptation hospitals and 91 evaluation hospitals (4,196 evaluation patients); the main unadapted four-database workflow instead evaluates all 4,983 eICU patients. CareVue and SICdb use patient-disjoint calibration. MIMIC retains the source identity map. Intervals use 500 paired patient resamples with fixed predictions and calibration; eICU hospital interval for capture change is −6.44 to 24.23.

#### Supplementary Table 45 Training with incomplete records under matched calibration procedures

| **Training** | **Source calibration** | **Matched local-calibration family** |
| --- | --- | --- |
| Standard | 144.33 | 156.67 |
| Missingness-adapted | 168.00 | 171.33 |
| Adapted − standard, patient CI | — | 14.67 (1.83 to 26.84) |
| Adapted − standard, hospital CI | — | 14.67 (3.20 to 23.95) |

Mean captured events over three persistent 50% record-batch masks; 4,196 eICU patients and 2,098 alerts. Nine registered origins and four-hour cooldown. Both models use the same selected calibration family and adaptation roles, but their coefficients are fitted separately on fully visible data from 23 hospitals, then transferred unchanged to masked views in the other 91 hospitals. Intervals average corresponding mask draws before paired subtraction (500 draws); they do not treat masks as independent patient cohorts.

#### Supplementary Table 46 Calibration of the organ actually nominated at index

| **Database** | **Calibration** | **Selected patients** | **Predicted risk (%)** | **Observed events (%)** |
| --- | --- | --- | --- | --- |
| MIMIC-IV | Source | 820 | 20.85 | 21.34 |
| MIMIC-IV | Organ-specific | 864 | 21.29 | 20.37 |
| eICU | Source | 1341 | 13.04 | 9.32 |
| eICU | Organ-specific | 1124 | 10.38 | 9.61 |
| CareVue | Source | 486 | 14.55 | 12.35 |
| CareVue | Organ-specific | 653 | 13.33 | 12.86 |
| SICdb | Source | 347 | 22.53 | 14.41 |
| SICdb | Organ-specific | 301 | 14.65 | 17.61 |

At index, select the highest-risk eligible organ and retain patients above 5% predicted risk. Predicted and observed values refer to that nominated organ, not to the mean of all six organ risks. Patient-disjoint five-fold calibration with three inner folds is evaluated over each complete database cohort, including all 4,983 eICU patients. Recalibration can change both the nominated organ and selected patients; these are selected-group diagnostics, not a paired change in a fixed group.

#### Supplementary Table 47 Patient-time selection and organ nomination

| **Database** | **Patient and time selection** | **Organ nomination** | **Alerts** | **Captured events** |
| --- | --- | --- | --- | --- |
| MIMIC-IV | Current | Current | 980 | 363 |
| MIMIC-IV | Current | History | 980 | 364 |
| MIMIC-IV | History | Current | 980 | 379 |
| MIMIC-IV | History | History | 980 | 382 |
| eICU | Current | Current | 2492 | 219 |
| eICU | Current | History | 2492 | 223 |
| eICU | History | Current | 2492 | 237 |
| eICU | History | History | 2492 | 272 |
| CareVue | Current | Current | 644 | 108 |
| CareVue | Current | History | 644 | 110 |
| CareVue | History | Current | 644 | 124 |
| CareVue | History | History | 644 | 126 |
| SICdb | Current | Current | 257 | 26 |
| SICdb | Current | History | 257 | 28 |
| SICdb | History | Current | 257 | 42 |
| SICdb | History | History | 257 | 46 |

Current denotes the shared current-information baseline; History denotes ten-source history. Nine scheduled assessment origins and six-hour patient suppression, at 0.50 alerts per patient. Each row fixes the accepted patient-time slots from one policy and uses the other policy only to nominate an eligible organ at those slots. Distinct patient-organ-onset events are counted once. Cross-routing differences are predictive decompositions and are not additive causal contributions. These counts are not the hourly/four-hour workflow.
